# Frequent functional orthology of long noncoding RNAs and genomic loci associated with complex traits and disorders

**DOI:** 10.1101/2025.10.31.25339260

**Authors:** Mitchell J. Cummins, John S. Mattick

**Affiliations:** School of Biotechnology and Biomolecular Sciences, Faculty of Science, and UNSW RNA Institute, UNSW Sydney, NSW 2052, Australia

## Abstract

Genome wide association studies (GWAS) have identified many haplotype blocks linked to a wide range of complex human traits, including intelligence, neuropsychiatric disorders, and immunological disorders, among many others. Approximately a quarter of these haplotype blocks lack protein-coding sequences but most express long noncoding RNAs (lncRNAs). Here we show that human loci orthologous with mammalian lncRNAs involved in neurological or immunological functions are commonly associated with a human trait that is commensurate with the reported lncRNA function. We also show that for many neurological, autoimmune, and cancer complex traits, the vast majority (>90%) of associated haplotype blocks express one or more lncRNAs. We present a database of lncRNAs expressed from human haplotype blocks associated with GWAS traits, with their mouse orthologs, as a resource for functional analysis. Our analyses establish a framework for investigating the molecular etiology of complex traits and suggest a general solution to the challenge of diagnosing and treating complex disorders.

---

There have been thousands of genome-wide association studies (GWAS) conducted over the past two decades, a concerted effort to identify the genetic factors underpinning human complex traits and disorders^1^. GWAS relies on the different distribution between study and control populations of sentinel single nucleotide polymorphisms (SNPs) that are in linkage disequilibrium with other sequences within haplotype blocks (i.e., are rarely separated by meiotic recombination). The sentinel SNPs may or may not be causative for the observed differences in the trait between the studied populations^2^.

Most trait- and disease-associated SNPs are located in noncoding intronic and ‘intergenic’ sequences^2,3^ and the causative variations are assumed to be regulatory in nature^4,5^. Efforts to identify the relationship between sequence variations in haplotype blocks and the trait being studied have incorporated gene expression analyses to identify genes whose expression is correlated with the allelic configuration of the SNP, referred to as expression quantitative trait loci (eQTLs)^6^. While limited by the lack of availability of appropriate tissues and developmental stages^7^, such studies have achieved some success in identifying relevant pathways^8^, although the correlated protein-coding genes are often distal to the associated SNPs^9^, the strongest gene-trait associations are often with genes encoding long noncoding RNAs (lncRNAs)^10,11^, and the gene-trait associations are dependent on the developmental context and/or the tissue under observation^12,13^.

Consistent with these observations, many of the GWAS regions associated with complex traits, including neurological traits, map to enhancers, genetic loci that control the spatiotemporal patterns of gene expression during differentiation and development^4,11,14–18^. An early model of enhancer action speculated that transcription factors bound at enhancer loci are brought into contact with target gene promoters via a series of protein–protein interactions^19,20^. However, molecular evidence shows that the transcription factors bound by enhancers promote the transcription of lncRNAs from the enhancer locus (elncRNAs) and that the ensuing lncRNAs bind chromatin remodeling, transcription factors (TFs) and other effector proteins, guiding them to target loci through RNA:DNA:DNA triplexes into topologically-associated domains for the expression of target genes^21–29^. Recent work shows that many canonical TFs directly bind eRNAs with sub-µM affinities, and supports a complementary eRNA-TF recruitment/retention mechanism with TFs contacting enhancer-derived lncRNAs at their own chromatin sites^30^.

Direct evidence that enhancer function is mediated by lncRNAs is provided by the demonstrations that knockdown of enhancer-derived lncRNAs abrogates enhancer action^31–40^, that ectopic expression of elncRNAs increases the expression of genes targeted by the enhancer^41,42^, and that splicing and modification of elncRNAs modulate enhancer activity^43–46^.

There is abundant evidence that lncRNAs transcribed from enhancers play central roles in mammalian development^47^ and that lncRNAs are associated with complex traits^48–55^. Deep sequencing has also shown that lncRNAs overlapping trait-associated SNPs are specifically expressed in cell types relevant to the traits^56^, and that most GWAS regions associated with complex traits and disorders express lncRNAs^57,58^, which must be prime candidates for the molecular bases of these traits and disorders^59,60^.

Here we validate this contention by showing that the majority of lncRNAs with experimentally described functions in a mammalian species map to a human GWAS trait locus that is commensurate with the lncRNA function.

Based on this relationship we have generated a database of human haplotype blocks associated with each GWAS trait, the human lncRNAs located therein, and possible mouse orthologous lncRNAs from the syntenic loci (Fig. 1). This dataset allows for the design of knockout and knockdown experiments in rodents to test associations between lncRNAs emanating from human haplotype blocks associated with GWAS traits, and commensurate behaviors in rodents, an important step in understanding the molecular etiology of complex traits.

**Figure 1.**
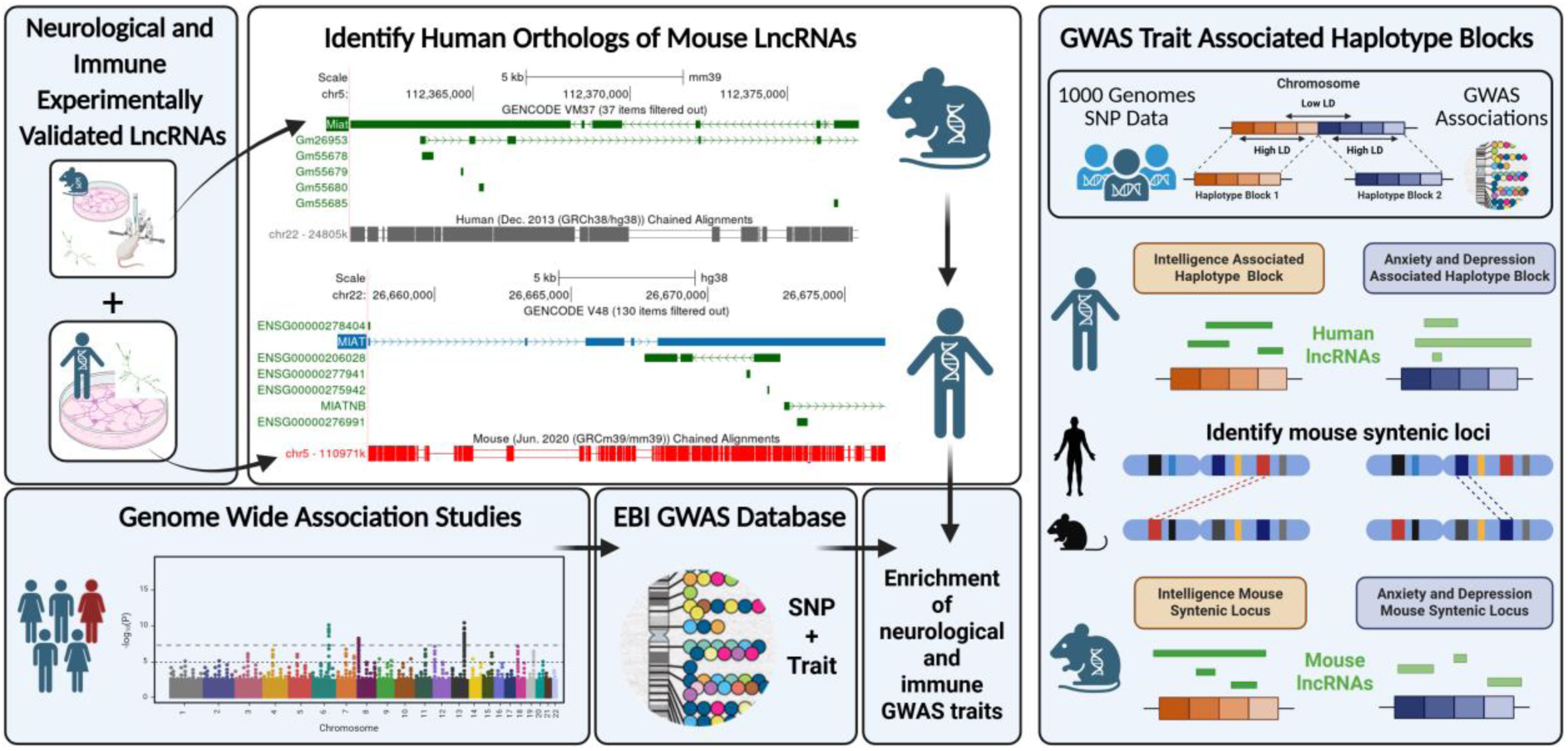
A database of GWAS trait associated human haplotype blocks containing lncRNAs. We identified mammalian neurological and immunological-related lncRNAs from the literature and identified the human syntenic loci. We searched the EBI-GWAS database for GWAS traits with associations in these loci and categorized traits as neurological/immunological or not. We found that lncRNAs with validated neurological functions were enriched for neurological GWAS traits, and that immunological lncRNAs were similarly enriched for immunological GWAS traits. Based on this relationship, we developed a database of human GWAS trait associated haplotype blocks, identified the lncRNAs produced from these blocks, and identified possible mouse orthologs for experimental examination. GWAS, Genome wide association study; LD, linkage disequilibrium; SNP single nucleotide polymorphism. Created with BioRender.com.

## Results

### LncRNAs with experimentally validated functions are frequently associated with commensurate GWAS traits

We collated 88 mammalian (mainly rodent and human) lncRNAs that have been reported to affect brain development, function and disorders (‘neurological lncRNAs’)^61^, and mapped the positions of their syntenic orthologs in the human genome. We found that ∼70% of these positions contain GWAS neurological phenotype associations, encompassing brain morphology, intelligence, educational attainment, behavior, and neurological disorders (Supplemental Table S1, Supplementary Material Online), examples of which are provided in Table 1.

**Table 1.** Examples of the correlation between functionally validated lncRNAs and commensurate GWAS traits.

| lncRNA | Reported function / phenotype | Human coordinates and ortholog | Neurological GWAS associations (controls ±SD) | Non-neurological GWAS associations (controls ±SD) | Examples of neurological GWAS associations |
| --- | --- | --- | --- | --- | --- |
| *BS-DRL1*<br>(mouse) | Modulates DNA damage response and genome stability in neurons<sup>133</sup>.<br>Knockout mice exhibit cell-type specific impairment of DNA damage response in the cortex and cerebellum and Purkinje cell degeneration<sup>133</sup>. | 15:89361579<br>-89424983<br>*MIR9-3HG* | 26<br>(0.4±0.7) | 18<br>(11.7±26.3) | • Alcohol use disorder measurement<br>• Anxiety measurement<br>• Attention deficit hyperactivity disorder<br>• Brain attribute<br>• Cannabis dependence<br>• Mathematical ability<br>• Schizophrenia<br>• Self-reported educational attainment<br>• Smoking behavior<br>• Substance abuse |
| *C130071C03*<br>*Riken*<br>(mouse) | Represses neural differentiation of mouse embryonic stem cells.<br>Knockdown of splice variants *Rik-201* and *Rik-203* inhibits neural differentiation *in vitro*.<br>Knockout of *Rik-201* and *Rik-203* *in vivo* causes retardation of embryonic brain development<sup>134</sup>. | 5:88408982-88691313<br>*MIR9-2HG* | 55<br>(8.1±5.6) | 68<br>(19.2±16.5) | • Autism spectrum disorder<br>• Bipolar disorder<br>• Cerebral cortex area attribute<br>• Cognitive function measurement<br>• Insomnia<br>• Intelligence<br>• Mood disorder<br>• Obsessive-compulsive disorder<br>• Substance abuse<br>• Tourette syndrome |
| *Emx2OS*<br>(mouse) | Regulates the expression of *EMX2*, a homeobox transcription factor implicated in several aspects of cerebral cortex development.<br>Knockdown of *Emx2OS* increases *Emx2* expression.<br>When delivered to rhombo-spinal precursors, stimulates ectopic expression of *Emx2*.<br>*Emx2* knock-out dramatically impairs *Emx2OS* transcription<sup>135</sup>. | 10:11747321<br>3-117545068<br>*EMX2OS* | 17<br>(1.9±1.8) | 20<br>(11±14.2) | • Alzheimer disease<br>• Anxiety measurement<br>• Bipolar disorder<br>• Cerebral cortex area attribute<br>• Cortical thickness<br>• Depressive symptom measurement<br>• Neuroticism measurement<br>• Obsessive-compulsive disorder<br>• PHF-tau measurement<br>• Schizophrenia |
| *Evf2* / *Dlx6os1* / *Dlx6as* (mouse) | Antisense to *Dlx6* homeobox gene.<br>Regulates interneuron subtype genes by recruiting Dlx and Mecp2 transcription factors.<br>Required for the formation GABAergic interneuronal circuitry<sup>62,63</sup>.<br>*Evf2* mouse mutants have reduced GABAergic interneurons in early postnatal hippocampus and dentate gyrus. GABAergic interneurons return to normal in *Evf2* mutant adult hippocampus, but have reduced synaptic inhibition<sup>62</sup>.<br>Mice lacking *Evf2* have increased seizure susceptibility and severity<sup>63</sup>. | 7:96955141-97014088<br>*DLX6-AS1* | 12<br>(0.9±1.2) | 12<br>(7.3±9.6) | • Attention deficit hyperactivity disorder<br>• Behcet's syndrome (includes increased seizure susceptibility and severity<sup>65</sup>)<br>• Educational attainment<br>• Intelligence<br>• PHF-tau measurement<br>• Self-reported educational attainment<br>• Smoking behavior<br>• Smoking initiation<br>• Social inhibition quality<br>• Substance abuse |
| *Linc-Brn1b*<br>(mouse) | Important for cortical lamination.<br>Knockout mice exhibit a decrease in proliferation of cortical progenitors *in vivo*, restricted to a specific subpopulation of progenitors in the subventricular zone of developing cortex.<br>Knockout also (consequently) results in reduction in intermediate progenitor cells in the cerebral cortex<sup>136</sup>. | 2:104865401-104874447<br>*LINC01159* | 7<br>(0.8±1.2) | 3<br>(1.4±1.4) | • Amygdala volume<br>• Brain attribute<br>• Cerebral cortex area attribute<br>• Cortical thickness<br>• Hippocampal volume<br>• Memory performance<br>• Neuroimaging measurement |
| *lncRNA_ES1*<br>/ *LINC01108*<br>(human) | Required for pluripotency. Knockdown resulted in downregulation of pluripotency markers and simultaneous upregulation of lineage markers including neuroectoderm, endoderm and mesoderm germ layers<sup>137</sup>. | 6:14280127-14285454 | 10<br>(0±0) | 24<br>(0.7±0.8) | • Educational attainment<br>• Intelligence<br>• Mathematical ability<br>• Memory performance<br>• Neurofibrillary tangles measurement<br>• Neuropsychological test<br>• Self-reported educational attainment<br>• Short-term memory<br>• Smoking initiation<br>• Vascular dementia |
| *NeuroLNC*<br>(mouse, rat) | Regulates neurite elongation, neuronal migration and presynaptic activity by interacting with the neurodegeneration-associated protein TDP-43<sup>138</sup>.<br>Expressed during retinal development<sup>139</sup>.<br>Overexpression drastically increases the release of synaptic vesicles from neurons<sup>138</sup>.<br>Overexpression significantly enhances Ca<sup>2+</sup> influx while down-regulation decreases the influx of Ca<sup>2+</sup><sup>138</sup>.<br>Down-regulation decreases neurite length and branching while overexpression has the opposite effect<sup>138</sup>.<br>Down-regulation decreases migration of neurons from the<br>ventricular zone to the cortical plate, while overexpression has the opposite effect<sup>138</sup>. | 8:9899744-9919233<br>*MIR124-1HG* | 23<br>(0.4±0.7) | 63<br>(2.5±1.6) | • Amygdala volume<br>• Bipolar disorder<br>• Brain connectivity attribute<br>• Cognitive function measurement<br>• Cortical thickness<br>• Epilepsy<br>• Neurotic disorder<br>• Schizophrenia<br>• Sleep duration trait<br>• White matter hyperintensity measurement |
| *Paupar*<br>(mouse) | Involved in neural differentiation and regulates neural gene expression.<br>Knockdown disrupts the cell cycle profile of neuroblastoma cells and induces neural differentiation<sup>140</sup>. | 11:31812307-32002405<br>*PAUPAR* | 17<br>(3.6±3.6) | 17<br>(18.6±20) | • Alzheimer disease<br>• Anorexia nervosa<br>• Attention deficit hyperactivity disorder<br>• Autism spectrum disorder<br>• Cognitive function measurement<br>• Major depressive disorder<br>• Obsessive-compulsive disorder<br>• PHF-tau measurement<br>• Schizophrenia<br>• Tourette syndrome |
| *Pnky / Gm30731*<br>(mouse) | Regulates neuronal differentiation of embryonic and postnatal neural stem cells<sup>141</sup>, neural stem cell migration<sup>142</sup> and production of projection neurons from neural stem cells<sup>143</sup>.<br>Regulates neurogenesis and modulates cognitive and affective behaviors<sup>144</sup>.<br>Knockdown increases neuronal differentiation in culture and *in vivo*<sup>141</sup>.<br>Silencing suppresses and overexpression promotes *in vitro* migration of neural stem cells<sup>142</sup>.<br>Conditional deletion in developing cortex regulates production of projection neurons from neural stem cells, altering postnatal cortical lamination<sup>143</sup>. | 6:98759637-98835826<br>*ENSG00000283010* and *ENSG00000309555* | 6<br>(0.8±1.5) | 6<br>(4.6±3) | • Attention deficit hyperactivity disorder<br>• Autism spectrum disorder<br>• Educational attainment<br>• Ganglion thickness<br>• Intelligence<br>• Memory performance |
|  | Knockout reduces fear context generalization in males and increases acoustic startle response in females<sup>144</sup>.<br>Transgenic *Pnky* expression rescues acoustic startle response in females<sup>144</sup>. |  |  |  |  |
| *Six3OS1* / *RNCR1*<br>(mouse) | Expressed during retinal development<sup>139</sup>.<br>Regulates retinal cell fate specification<sup>145</sup>.<br>Implicated in neuronal modulation by inhibiting glial cell differentiation<sup>146</sup>.<br>Knockdown results in decreased protein kinase C $\alpha$ (PKCα) positive rod bipolar cells and increased glutamine synthetase-positive Muller glia<sup>145</sup>.<br>Knockdown decreases the number of neurons and oligodendrocytes and increases the number of astrocytes in culture<sup>146</sup>. | 2:44921054-44939344<br>*LINC01833* | 34<br>(0.4±0.7) | 26<br>(1.2±1) | • Alcohol and nicotine co-dependence<br>• Amount of iron in brain<br>• Attention deficit hyperactivity disorder<br>• Brain volume<br>• Cognitive function measurement<br>• Cortical thickness<br>• Educational attainment<br>• Executive function measurement<br>• Opioid use disorder<br>• Self-reported educational attainment |

An illustrative case is the clinically relevant human GWAS trait associated with the conserved neurodevelopmental elncRNA *Evf2* (human ortholog *DLX6-AS1*)^62,63^. *Evf2* is critical for formation of a subset of GABAergic interneurons in mice^62^, resulting in reduced inhibition and increased seizure susceptibility^63^. As far as we are aware, there are not yet any clinical reports of *Evf2* mutations in epilepsy patients, likely because most analyses use exome sequencing. However, an intronic SNP in *Evf2* (rs731263) is associated with the inflammatory disorder Behçet’s disease^64^. A rare (∼5%) manifestation of this disease has neurological involvement (Neuro-Behçet’s disease), which may involve epileptic seizures^65^. It has yet to be determined if *Evf2* variations in Neuro-Behçet’s disease patients predicts seizure susceptibility.

A confounding factor is the fact that GWAS associated haplotype blocks cover over half of the genome and many exhibit pleiotropy, i.e., are associated with multiple traits^66^: ∼50% of the neurological lncRNAs are also located in a GWAS region associated with an immunological phenotype (Supplemental Table S1, Supplementary Material Online and Table 2). The neurologically validated lncRNAs have correlations with 509 GWAS-associated neurological traits, and 177 GWAS-associated immunological traits (∼2.9-fold enrichment).

**Table 2.**
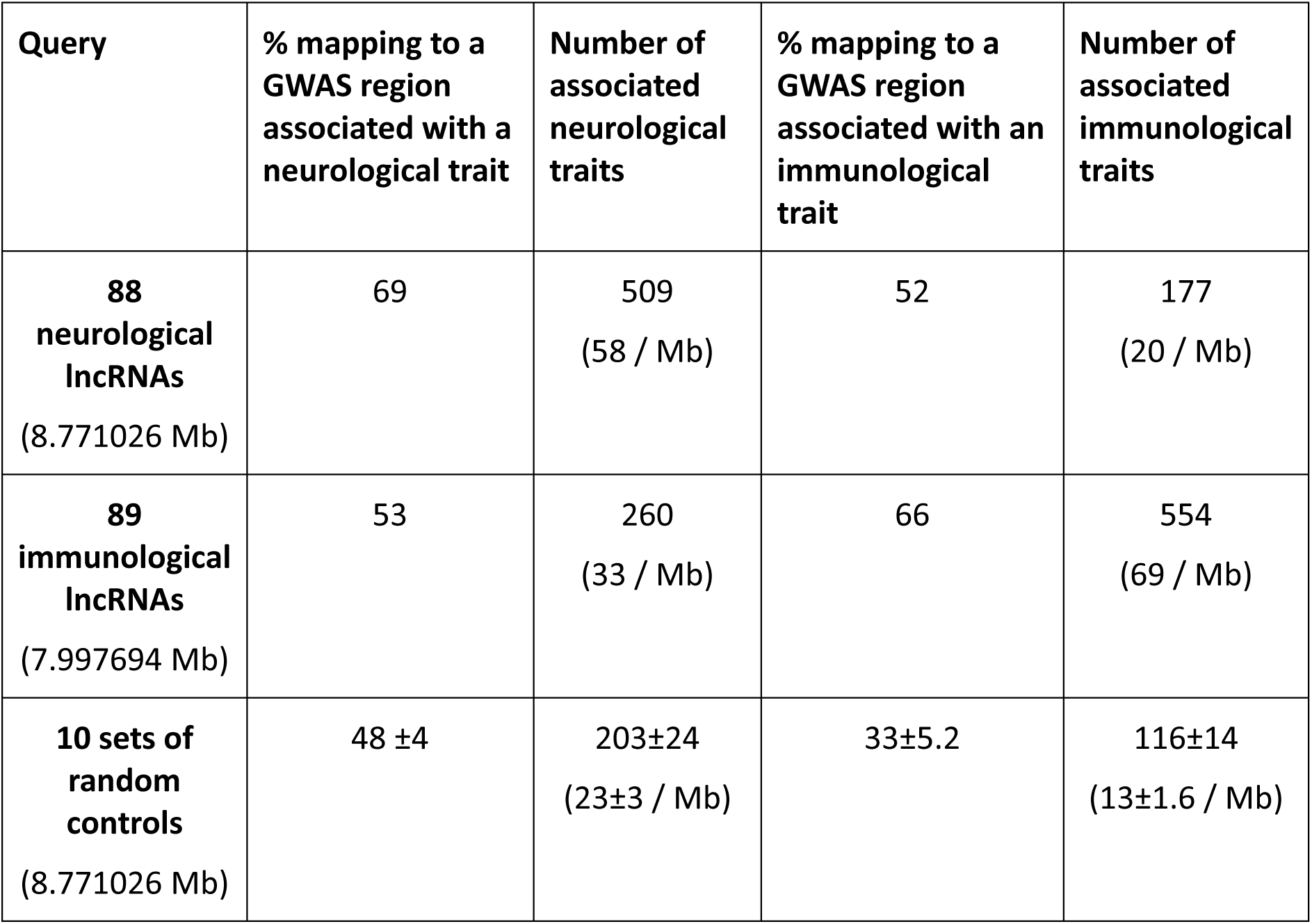
Correlation between functionally validated lncRNAs and commensurate GWAS traits. All values rounded to nearest whole number.

We therefore undertook a reciprocal analysis for 89 lncRNAs that have been functionally validated to affect immunological processes (‘immunological lncRNAs’, encompassing immunological phenomena, immunomodulators and immunological disorders). We found that ∼66% of the immunological lncRNAs map to an orthologous GWAS position associated with an immunological trait, and ∼53% to an orthologous GWAS position associated with a neurological trait (Supplemental Table S2, Supplementary Material Online), with a 2.1-fold enrichment of the number of immunological versus neurological traits (Table 2). The higher crossover is not necessarily surprising as there is considerable interaction and molecular overlap between the immunological and nervous systems.

To test the specificity of the correlations we also analyzed the GWAS associations of 10 sets of number and size-matched (to neurological lncRNAs) sequences randomly chosen from the human genome. Control sequences were taken from the same chromosome as their associated lncRNA. We found that ∼48±4% of shuffled coordinates contained a neurological GWAS trait in the locus (Table 2, Odds ratio 2.4744, 95% CI 1.3356-4.5843, z-statistic 2.880, p = 0.0040), and the sets contained on average ∼200 neurological traits, less than half of the validated neurological lncRNAs. It is not surprising that a high percentage of shuffled sequences overlap neurological GWAS SNP-trait associations, as the brain expresses a large fraction of the genome^67,68^, especially during development^69,70^. We also found that fewer shuffled sequences contained immunological traits compared to validated immunological lncRNA loci (∼33% v ∼66%, Odds ratio 4.0011, 95% CI 2.1415-7.4759, z-statistic 4.348, p < 0.0001), and that shuffled sequences contained fewer immunological traits (∼13 v ∼69 / Mb).

### A database of lncRNAs expressed from human haplotype blocks associated with GWAS traits and their mouse orthologs

Based on the findings that many human orthologs of rodent lncRNAs have GWAS associations that are commensurate with the described function, and that many human lncRNAs with described functions have rodent orthologs, we developed a database of human lncRNAs expressed from haplotype blocks associated with GWAS traits, and their possible mouse orthologs (available at https://github.com/Mitchell-Cummins/GWAS_trait_haplotype_blocks_lncRNA). To do this, we generated linkage disequilibrium (LD) blocks from the 1000 genomes project^71^ data (hg38 build) with all SNPs in high LD assumed to compromise a haplotype block. We intersected the blocks with human gene coordinates to generate 4 sets of blocks: (1) All blocks, (2) Blocks containing human lncRNAs, (3) Blocks containing human lncRNAs and not protein-coding genes, (4) Blocks without known lncRNAs or protein-coding genes. We then intersected the haplotype blocks with GWAS SNPs from the NHGRI-EBI GWAS catalog^72^ for each GWAS trait, to generate trait-associated blocks and counts of associations for each trait. The numbers of blocks for each set are summarized in Fig. 2a.

**Figure 2.**
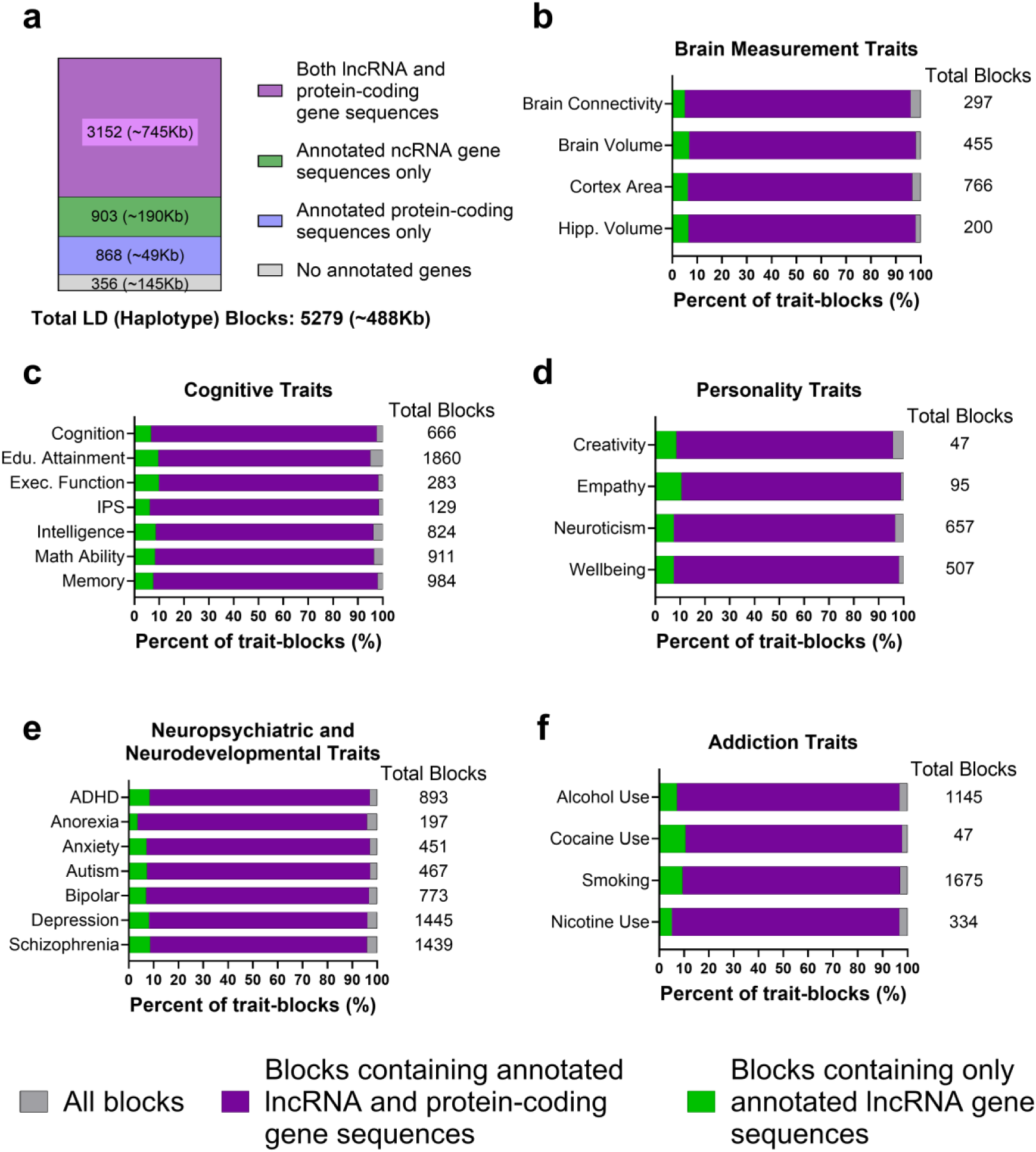
Summary of haplotype blocks containing lncRNAs. (**a**) Numbers of all GWAS trait-associated haplotype blocks containing protein coding genes, lncRNAs, both (mixed), or neither (no gene). (**b-f**) Percentages of haplotype blocks containing lncRNAs for (b) brain measurement traits, (c) cognitive traits, (d) personality traits, (e) neuropsychiatric and neurodevelopmental disorders, (f) addiction traits. Bars of block types are superimposed over one another. ADHD, attention-deficit hyperactivity disorder; Edu, educational; Exec, executive; Hipp, hippocampal; IPS, information processing speed.

We also summarized the percentages of blocks containing lncRNAs for a subset of neurological traits including those involved in brain measurements (Fig. 2b), cognition (Fig. 2c), personality (Fig. 2d), neuropsychiatric and neurodevelopmental disorders (Fig. 2e), and addiction (Fig. 2f). Percentages of blocks containing lncRNAs for some autoimmune disorders (Fig. S1a Supplementary Material Online) and cancers (Fig. S1b Supplementary Material Online) are summarized in the supplements. The results show that the vast majority (>90%) of haplotype blocks associated with complex traits by GWAS express one or more lncRNAs.

For all trait-associated blocks, we identified mouse syntenic loci using the UCSC^73^ program *liftOver*, only requiring 10% of the block to align between species as many lncRNAs contain only patches of sequence alignment between human and mouse. We intersected these syntenic loci with mouse genes to identify likely orthologs. Using this method we have identified human haplotype blocks and mouse syntenic loci containing lncRNAs (but not protein-coding genes) associated with brain structure (Fig. S2a-b Supplementary Material Online) and cognitive traits (Fig. S2c-f Supplementary Material Online), as well as sleep (Fig. 3a-b Supplementary Material Online), mood (Fig. S3c-d Supplementary Material Online), addiction (Fig. S3e-f Supplementary Material Online) and neurodevelopmental (Fig. S3g-h Supplementary Material Online) disorders. Many of these blocks contained associations with other neurological traits. For example, the intelligence block (Fig. S2d) also contained the greatest number of autism, bipolar, and attention deficit/hyperactivity disorder associations.

**Figure 3.**
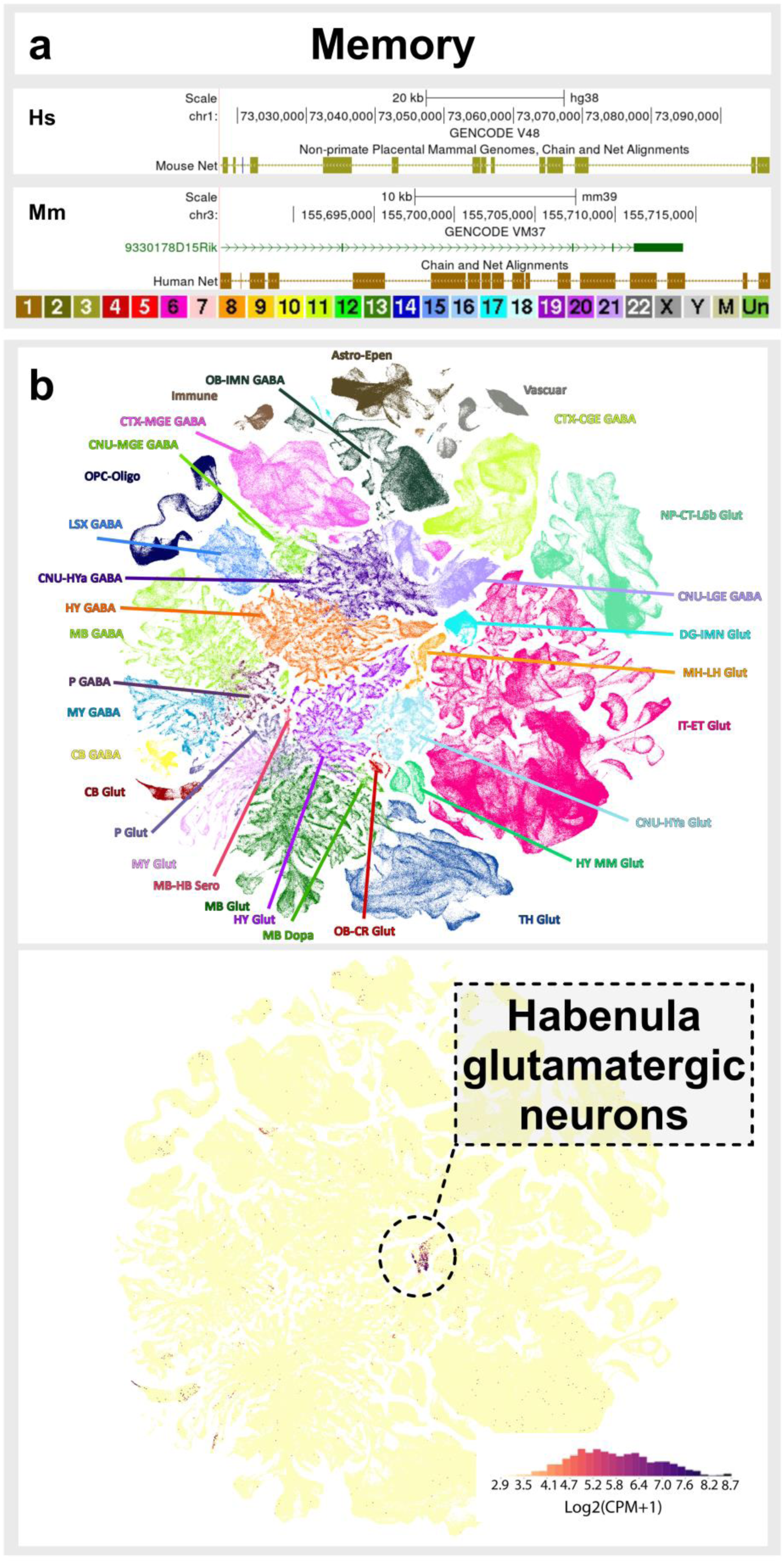
A human haplotype block associated with memory by GWAS expressing a lncRNA in the mouse syntenic locus. (**a**) UCSC genome browser view of a haplotype block containing 25 GWAS associations with memory but no annotated human gene and the mouse syntenic locus. Gene names: Green, annotated non-coding; pink, pseudogene; red, problem transcripts (retained introns, to be experimentally confirmed, or disrupted domains); blue, coding. Hs, *Homo sapiens*; Mm, *Mus musculus*. Chronograph of chromosome number applies to both browser images. (**b**) Single cell sequencing data from the ABC Atlas 10x scRNAseq whole brain data indicating (top) cell groups, and (bottom) expression of *9330178D15Rik*. CPM, counts per million. Astro, astrocyte; CB, cerebellum; CGE, caudal ganglionic eminence; CNU, cerebral nuclei; CR, Cajal–Retzius; CT, corticothalamic; CTX, cerebral cortex; DG, dentate gyrus; Epen, ependymal; ET, extratelencephalic; HB, hindbrain; HY, hypothalamus; HYa, anterior hypothalamic; IMN, immature neurons; IT, intratelencephalic; L6b, layer 6b; LGE, lateral ganglionic eminence; LH, lateral habenula; LSX, lateral septal complex; MB, midbrain; MGE, medial ganglionic eminence; MH, medial habenula; MM, medial mammillary nucleus; MY, medulla; NP, near-projecting; OB, olfactory bulb; Oligo, oligodendrocytes; OPC, oligodendrocyte precursor cells; P, pons; TH, thalamus. Neurotransmitter types shown: Dopa, dopaminergic; GABA, GABAergic; Glut, glutamatergic; Sero, serotonergic.

To demonstrate generality, we also provide examples of human haplotype blocks and the mouse equivalents associated with general physiological traits (Fig. S4a-e Supplementary Material Online), immunological traits (Fig. S5a-d Supplementary Material Online), cancers (Fig. S6a-d Supplementary Material Online), and idiopathic diseases (Fig. S7a-b Supplementary Material Online).

Interestingly, there are no identified protein-coding or lncRNA genes in some GWAS trait-associated haplotype blocks in the human annotation, whereas there are in the mouse syntenic locus. For example, we identified a human haplotype block containing 25 GWAS memory associations in humans but no annotated gene (Fig. 3a, top). However, the mouse syntenic locus contains a single lncRNA (*9330178D15Rik*) gene (Fig. 3a, bottom) that is highly expressed in habenula glutamatergic neurons in the ABCAtlas^74^ (Fig. 3b). The epithalamic habenula nuclei are highly conserved in vertebrates^75^, are involved in a range of behaviors^76,77^, including anxiety^78^ and memory^79,80^ in rodents, and are thought to be essential to adaptive behaviors in complex tasks^81^. Our data suggest *9330178D15Rik* is a prime candidate for molecular investigation into the mechanisms underlying habenula control of adaptive behaviors, and we speculate a yet to be identified human ortholog explains the GWAS memory associations.

We found similar blocks lacking protein-coding or lncRNA genes for major depressive disorder (4 associations, Fig. S8a, Supplementary Material Online), alcohol consumption (6 associations Fig. S8b, Supplementary Material Online), and smoking initiation (5 associations Fig. S8c, Supplementary Material Online), although these human blocks contained pseudogenes, which may produce lncRNAs^47^. We also discovered GWAS trait-associated blocks that lack annotated human genes, but have mouse lncRNAs, for cancer and immunological cells (3 associations each, Fig. S8d, Supplementary Material Online) and diabetes mellitus (28 associations, Fig. S8e, Supplementary Material Online).

## Discussion

While many genomic loci associated with human complex traits have been identified by GWAS, the molecules responsible for these associations have been difficult to ascertain. Since some haplotype blocks associated by GWAS with neuropsychiatric and other complex traits express lncRNAs but not protein-coding genes^57,58^, lncRNAs are the most likely molecular candidates for many GWAS SNP-trait associations. This has been demonstrated for lncRNAs *ANRIL* and *LINC00305* in atherosclerosis^54,60,82^, *KILR*^83^ in breast cancer susceptibility, and implicated in many other cases^84–86^.

There is a compelling body of literature suggesting that lncRNAs are the prime regulatory molecules driving neurodevelopmental patterning (and development generally). In particular, it is becoming apparent that lncRNAs mediate the action enhancers^29,47^, which control the spatiotemporal patterns of gene expression by acting as scaffolds and guides for effector proteins in the formation of topologically-associated chromatin domains^87^. Approximately 40,000 of these enhancers have been found to exhibit dynamic epigenomic rearrangement during early human brain development and are enriched for GWAS loci associated with brain-related traits and disorders^88^. They are also thought to have driven the evolution of the uniquely human aspects of neurodevelopment that resulted in increased neocortex size and enhanced cognitive abilities compared to other hominids and mammals generally^89–94^.

Our conclusion that lncRNA products of GWAS loci are responsible for the observed trait differences in the population is consistent with the observations that sequence variation within enhancers is linked to the risk of neurological and psychiatric disorders^95^ and that GWAS hits are enriched in ‘long enhancers’ with the strongest selective constraints distal to the TSS^96^.

Thus far, despite the substantial circumstantial evidence, the generality of lncRNA etiology of variations in complex traits and disorders has been difficult to test. We propose that variation in many complex traits will be explained by lncRNA sequence variation and consequent shifts in intermolecular interactions with DNA, RNA and protein binding partners^97^. In support of this contention, the results presented herein suggest not only a frequent functional association of lncRNAs with GWAS-identified traits, but also that many rodent lncRNAs are functionally orthologous to humans. While it is well known that lncRNAs evolve more rapidly than protein-coding genes, due to more relaxed structure-function relationships and positive selection for adaptive radiation^47^, many have orthologs that exhibit positional conservation^98^. The frequent pleiotropy of GWAS loci reflects the hierarchical and canalized sequence of gene regulatory networks in development, wherein a fraction of embryonic regulatory RNAs act at the top of the hierarchy, with broad effects, whereas others are expressed with greater cellular specificity and play roles at the periphery of development, affecting the terminal features of specialized cells”^99^.

The correlation with GWAS traits suggests conservation of (approximate) lncRNA functions between species for many lncRNAs, which has been previously demonstrated for subsets of lncRNAs involved in embryonic development^100^. Together, these findings allow for the design of experiments to test causal relationships between lncRNAs and human GWAS associated complex traits.

Although causal relationships between molecules and complex traits cannot be easily studied in humans due to ethical and experimental limitations of genetic manipulation, these relationships can be studied in rodents where genetic manipulations are routine, allowing the mechanistic study of human complex traits and behavior by proxy, provided appropriate behavioral paradigms exist or can be developed. In a recent review on the emerging roles of lncRNAs in the nervous system^61^, we advocate for the widespread adoption of modern behavioral techniques, such as the use of touchscreen computer testing to mimic human cognitive tests^101,102^, rodent virtual reality goggles^103^ and automated artificial intelligence (AI)-assisted visual tracking of behavior^104,105^ to make high throughput investigation of lncRNA-neurological trait associations more tractable.

While the relationships we have demonstrated appear to apply generally, we expect these results to be especially important to the cognitive, neuropsychiatric, and neurodevelopmental fields. There are now a number of examples demonstrating lncRNA control of cognitive traits such as fear extinction (e.g., *ADRAM*^106^, *CDR1-AS*^107^, *Gas5*^108^, and *SLAMR*^109^), short term memory (e.g., *Carip*^110^, *LoNa*^111^, *Neat1*^112^, and *Snhg11*^113^). In the neuropsychiatric field lncRNAs have been associated with addiction (e.g., *BDNF-AS*^114,115^ and *GAS5*^116^), mood disorders (e.g., *AP1AR-DT*^117^, *FEDORA*^118^ and *Miat*^119,120^), and social deficits (e.g., *Neat1*^121^ and *Synage*^122^). The associations between lncRNA loci and neuropsychiatric disorders are particularly timely given a recent review of neuropsychiatric genetics^123^ overlooked the role of lncRNAs in neuropsychiatric brain functions, their correlations with psychiatric disorders, and their roles in the brain generally^47,61,124^.

If the targeting of rodent orthologs of lncRNAs expressed from genomic regions associated with complex human traits and disorders identifies a commensurate phenotype, we suggest that the next step would be to undertake high resolution sequencing of the region (after PCR amplification) from a cohort of affected individuals to look for consistent sequence and/or structural variations in the encoded lncRNA(s). Phenotypic analysis of the neurodevelopmental effects of human lncRNAs and their variants can be undertaken using CRISPR-Cas13 or ASO-mediated knockdown in brain organoid cultures. The high visibility of most complex traits in family histories suggests that variant regulatory sequences are co-dominant and should be amenable to reduction by ASO-targeted knockdown *in vivo*, providing a general pathway to early diagnosis of, and a generic platform for therapeutic solutions to, the complex disorders that constitute the major health burden of the population.

## Methods

### Identification of human and rodent long noncoding RNAs associated with neurological and immunological functions

Human and rodent lncRNAs associated with neurological functions were identified by literature search. Many were identified as part of a recent review^61^. LncRNAs were searched for by coordinates in the UCSC Genome Browser^73^ GRCh38 v485 mm39 (mouse) or hg38 (human) build to avoid differences in lncRNA names. For mouse lncRNAs, human syntenic loci were selected from the Placental Chains/Nets UCSC Genome Browser track to identify possible human orthologs. Orthologs were required to have the same strand orientation between species. Where orthologs were identified, the genomic coordinates of the ortholog were used to identify GWAS SNP-trait associations using the NHGRI-EBI GWAS catalog (https://www.ebi.ac.uk/gwas/)^72^. Where no ortholog was found but species alignment was present, the genomic coordinates of the syntenic loci were used. Human lncRNAs associated with neurological functions were searched by genomic coordinates for GWAS associations. The trait labels, as opposed to reported traits, were used to ensure consistency between studies. All associations in the GWAS catalog with a SNP-trait p-value <1.0 x 10-5 were included (see https://www.ebi.ac.uk/gwas/docs/methods/criteria for more details on GWAS inclusion criteria).

Immunological associated lncRNAs were identified from four review articles^125–128^ supplemented by a literature search. Human orthologs of rodent lncRNAs and GWAS associations were identified following the same method as for neurological lncRNAs.

### Generation of human haplotype blocks by linkage disequilibrium

Human linkage disequilibrium (LD) blocks were generated from the 1000 genomes project^71^ hg38 variant call format (VCF) files (available from https://ftp.1000genomes.ebi.ac.uk/vol1/ftp/data_collections/1000_genomes_project/releas <u>e/20181203_biallelic_SNV/</u>). VCF files were converted to binary variant call format (BCF) files using *bcftools*^129^. Variants were pruned from BCF files using *PLINKv1.9*^130^ with the following settings --maf 0.10 --indep 50 5 10 to select variants in high LD. Browser extensible data (BED) files were generated and all variants within 10kb of another variant were merged into a single haplotype block, and blocks <1000bp were discarded. Haplotype blocks were intersected using *bedtools intersect*^131^ with BED files of protein-coding genes, and lncRNA genes to generate 4 sets of blocks; 1. All blocks. 2. Blocks with lncRNAs. 3. Blocks with lncRNAs but no protein coding genes. 4. Blocks with no annotated genes (protein coding or lncRNAs). BED files of genes were generated from the Ensembl hg38 gene transfer format (GTF) file (available at https://ftp.ensembl.org/pub/release-113/gtf/homo_sapiens/Homo_sapiens.GRCh38.113.chr.gtf.gz).

### Identification of GWAS trait-associated human haplotype blocks and mouse orthologs

A file of all published associations in the GWAS catalog (v1.0.2) was downloaded (available at (https://www.ebi.ac.uk/gwas/api/search/downloads/studies/v1.0.2.1) and a custom script used to generate a BED file of all associations with the mapped trait included as a column and a list of all mapped traits. These were used to generate BED files for all traits separately. The haplotype block BED files were then intersected with the trait BED files using *bedtools intersect*. To identify human lncRNAs in these blocks, a bed file of all human lncRNAs (lncRNAs and TEC genes) was generated from the Ensembl^132^ hg38 GTF file (available at https://ftp.ensembl.org/pub/release-113/gtf/homo_sapiens/Homo_sapiens.GRCh38.113.chr.gtf.gz). This lncRNA BED file was then intersected using *bedtools intersect* with the trait-associated block BED files.

The UCSC tool *liftOver* was then used to identify the mouse syntenic loci for all trait associated blocks from the hg38ToMm39 chain file (available at https://hgdownload.soe.ucsc.edu/goldenPath/hg38/liftOver/hg38ToMm39.over.chain.gz). The following settings were used -minMatch=0.1 -multiple -minSizeQ=10000 - minChainT=5000. To identify rodent lncRNAs in these blocks, a bed file of all mouse lncRNAs (lncRNAs and TEC genes) was generated from the Ensembl mm39 GTF file (available at https://ftp.ensembl.org/pub/release-115/gtf/mus_musculus/Mus_musculus.GRCm39.115.gtf.gz). This lncRNA BED file was then intersected using *bedtools intersect* with the mouse syntenic loci BED files for all traits.

## Acknowledgements

We would like to thank Saba Altaf who compiled a list of neurological lncRNAs for a recent review, the basis on which our neurological lncRNAs are founded. We would like to thank Kevin Blighe for making the code for generating linkage disequilibrium blocks from 1000 genomes variant data publicly available (available at https://www.biostars.org/p/335605/). We thank Martin Smith (UNSW) and Grant Montgomery (University of Queensland) for helpful comments and suggestions. Analyses were performed on the UNSW Compute Cluster Katana (https://researchdata.edu.au/katana/1733007).

## Author Contributions

J.S.M. and M.J.C. conceived the study, identified neurological and immunological lncRNAs through literature search, and generated tables. M.J.C. intersected GWAS traits with neurological and immunological lncRNAs and controls, generated the database of GWAS associated haplotype blocks with mouse syntenic loci and lncRNAs, and prepared the figures. J.S.M. and M.J.C drafted and edited the manuscript.

## Data Availability

Datasets used in this study were obtained from publicly available databases as described in the methods section. Scripts to generate the datasets and the database of GWAS haplotype blocks are available on GitHub (https://github.com/Mitchell-Cummins/GWAS_trait_haplotype_blocks_lncRNA). All data are available from the authors upon request.

Track images were taken from the UCSC Genome Browser. Graphs and figures were generated using Graphpad Prism 9.

