## Supplemental Table S1 for "Frequent functional orthology of long noncoding RNAs and genomic loci associated with complex traits and disorders"

### Supplementary Table S1. GWAS correlates of lncRNAs associated with neurological functions

LncRNAs implicated in neurological processes whose orthologous positions correlate with a GWAS neurological association are highlighted in green. Immunological associations are highlighted in yellow.

| lncRNA | Reported function / phenotype | Human coordinates and possible human ortholog | Neurological GWAS associations | Non-brain/neural GWAS associations |
|---|---|---|---|---|
| 1. *1500011B03Rik / Gm57857 / ncRNA_A* (mouse) | High expression pattern during the early stages of neuronal reprogramming, as well as in postnatal and adult mouse brain[1]. | 12:110027028-110073636<br>*C12orf76* | • Alcohol consumption quality<br>• Bipolar disorder<br>• Cognitive function measurement<br>• Intelligence<br>• JT interval<br>• Schizophrenia | • Body height<br>• Forced expiratory volume<br>• Hematocrit<br>• High density lipoprotein cholesterol measurement<br>• Hypothyroidism<br>• Platelet volume<br>• Susceptibility to childhood ear infection measurement |

| | | | | |
|---|---|---|---|---|
| 2. *1600020E01Rik / ncRNA_AA* (mouse) | High expression pattern during the early stages of neuronal reprogramming, as well as in postnatal and adult mouse and postnatal human brain[1] | 2:69962263-70103753<br>*PCBP1-AS1 / h.ncRNA_AA* | • None | • Anti-citrullinated protein antibody seropositivity<br>• Appendicular lean mass<br>• Body height<br>• Erythrocyte count<br>• High density lipoprotein cholesterol measurement<br>• Hypothyroidism<br>• IgF-1 measurement<br>• Leukocyte quantity<br>• Rheumatoid arthritis<br>• Rheumatoid factor seropositivity measurement<br>• Thyroid stimulating hormone amount<br>• Total cholesterol measurement<br>• Triglyceride measurement<br>• Vital capacity<br>• Whole body water mass |

| | | | | |
|---|---|---|---|---|
| 3. *2610316D01Rik / ncRNA_B* (mouse) | High expression pattern during the early stages of neuronal reprogramming, as well as in postnatal and adult mouse brain[1]. Also expressed in human brain[1]. | 4:133075311-133151069<br>*PCDH10-DT* | • Memory performance<br>• Memory performance<br>• Reading and spelling ability<br>• Rolandic epilepsy<br>• Short-term memory | • Abnormal vaginal discharge itching<br>• Allergen exposure measurement<br>• Amino acid measurement<br>• Breast carcinoma<br>• Breastfeeding duration<br>• C-C motif chemokine 2 level<br>• Endometrial carcinoma<br>• Gait quality<br>• Gut microbiome measurement<br>• Insulin measurement<br>• Number of siblings<br>• Prostate cancer<br>• Protein measurement<br>• Sex interaction measurement<br>• Vaginal microbiome measurement |

| | | | | |
|---|---|---|---|---|
| 4. *5031425E22Rik / ncRNA_C* (mouse) | High expression pattern during the early stages of neuronal reprogramming, as well as in postnatal and adult mouse brain[1]. Also expressed in human brain[1]. | 7:104894628-105014321<br><br>*KMT2E-AS1 / h.ncRNA_B / h.ncRNA_C* | • Alcohol drinking<br>• Cognitive function measurement<br>• Dyslexia<br>• Educational attainment<br>• Income<br>• Insomnia<br>• Intelligence<br>• Mathematical ability<br>• Post-traumatic stress disorder<br>• Risk-taking behaviour<br>• Schizophrenia<br>• Sleep duration trait<br>• Socioeconomic status | • Diastolic blood pressure<br>• Interleukin-6 measurement<br>• Low density lipoprotein cholesterol measurement<br>• Metabolic syndrome<br>• Peak expiratory flow<br>• Protein measurement<br>• Systolic blood pressure<br>• Vital capacity<br>• Vitamin D amount |

| | | | | |
|---|---|---|---|---|
| 5. *ADEPTR / Gm38257* (mouse) | Intronic lncRNA required for the activity-dependent structural plasticity of dendritic spines and synaptic transmission.<br><br>Knockdown suppresses activity-dependent changes synaptic transmission, reduces cAMP-dependent up-regulation of excitatory postsynaptic current amplitude and structural plasticity of dendritic spines by reducing mushroom spine density[2]. | 10:18661193-18663548<br><br>None | • Sudden cardiac arrest[a] | • None |
| 6. *ADRAM* (mouse) | Enhancer-derived lncRNA induced by experience epigenetically regulates *Nr4a2* and is required for the formation of fear extinction memory.<br><br>Knockdown impairs fear extinction memory without effecting anxiety-like behaviour[4]. | 2:156345249-156414988<br><br>None (opposite strand ENSG00000309 294 ; ENSG00000310 173) | • Schizophrenia<br>• Neuroticism measurement | • Body height |

[a] Aberrant nerve function ('channelopathy') is a cause of 15-30% of sudden cardiac arrests[3].

| | | | | |
|---|---|---|---|---|
| 7. *ALAE* (rat, mouse) | Required locally for axon elongation via regulation of local mRNA translation[5].<br><br>Knockdown reduces axon elongation[5].<br><br>Drives stress granule formation in tauopathies. Significantly reduced in a mouse model of tauopathy and in FTLD-tau, progressive supranuclear palsy, and Alzheimer's disease brains[6]. | 4:118278462-118285316<br><br>*SNHG8* | • Neuroticism measurement | • Childhood trauma measurement |
| 8. *AP1AR-DT* (human) | A bipolar disorder-associated upregulated long non-coding RNA[7].<br><br>Human lncRNA with no sequence homology in mouse but ectopic expression of *AP1AR-DT* in mice induces depressive and anxiety-like behaviours by reducing Negr1 (neuronal growth regulator 1)-mediated excitatory synaptic transmission, which can be overcome by Negr1 overexpression[7]. | 4:112229561-112231596 | • Neuroticism measurement<br><br>• Schizophrenia | • Cleft lip<br>• Cleft palate<br>• Disorder of pharynx<br>• Lymphocyte count |

| | | | | |
|---|---|---|---|---|
| 9. *AtLAS* (mouse) | Regulates social hierarchy by modulating postsynaptic AMPA receptor trafficking.<br><br>Modulating expression alters social hierarchy, with silencing promoting increased social dominance[8]. | 3:12178692-12234274<br><br>*ENSG00000288952* | • Alcohol drinking<br>• Alcohol consumption quality<br>• Depressive symptom measurement<br>• Neuroticism measurement<br>• Wellbeing measurement | • Apolipoprotein B measurement<br>• Basophil count<br>• Basophil measurement<br>• Bilirubin measurement<br>• BMI-adjusted hip circumference<br>• BMI-adjusted waist-hip ratio<br>• BMI-adjusted waist circumference<br>• Body height<br>• Bone density<br>• BPI fold-containing family B member 1 measurement<br>• Cholesterol:total lipids ratio<br>• Choline measurement<br>• Chronic kidney disease<br>• Diabetic nephropathy<br>• Eosinophil count<br>• Erythrocyte volume<br>• Fatty acid amount<br>• Glomerular filtration rate<br>• HbA1c measurement<br>• Heel bone mineral density<br>• Hematocrit<br>• Hematological measurement<br>• Hemoglobin A1 measurement<br>• Hemorrhagic disease<br>• High density lipoprotein cholesterol measurement |

| | | | | |
|---|---|---|---|---|
| | | | | • Hypercholesterolemia<br>• Hyperlipidemia<br>• Hypothyroidism<br>• Interleukin-27 measurement<br>• Kunitz-type protease inhibitor 1 measurement<br>• Lactadherin measurement<br>• LAG3/VCAM1 protein level ratio in blood<br>• Leukocyte quantity<br>• Level of receptor-type tyrosine-protein phosphatase beta in blood<br>• Linoleic acid measurement<br>• Low density lipoprotein cholesterol measurement<br>• Lymphocyte activation gene 3 protein level<br>• Lymphocyte amount<br>• Lymphocyte count<br>• Mean corpuscular hemoglobin concentration<br>• Mean reticulocyte volume<br>• Metabolic disease<br>• Monocyte count<br>• Myeloid leukocyte count<br>• Neutrophil count<br>• Neutrophil measurement<br>• Neutrophil percentage of leukocytes |

| | | | | |
|---|---|---|---|---|
| | | | | • Non-alcoholic fatty liver disease<br>• Omega-6 polyunsaturated fatty acid measurement<br>• Phosphoglycerides measurement<br>• Platelet count<br>• Platelet crit<br>• Platelet quantity<br>• Polypeptide N-acetylgalactosaminyltransferase 3 measurement<br>• Polyunsaturated fatty acid measurement<br>• Protein HEG homolog 1 measurement<br>• Red cell distribution width<br>• Remnant cholesterol measurement<br>• Reticulocyte amount<br>• Saturated fatty acids measurement<br>• Spleen volume<br>• Systemic lupus erythematosus<br>• Thyroid stimulating hormone amount<br>• Total blood protein measurement<br>• Total cholesterol measurement |

| | | | | |
|---|---|---|---|---|
| | | | | • Wap, kazal, immunoglobulin, kunitz and ntr domain-containing protein 1 measurement |

| | | | | |
|---|---|---|---|---|
| 10. *ATXN2-AS* (human) | Associated with Spinocerebellar ataxia type II (SCA2) and amyotrophic lateral sclerosis (ALS).<br><br>*ATXN2-AS* contains the repeat as a CUG tract. CUG repeat expansions are toxic in a SCA2 cell model and form RNA foci in SCA2 cerebellar Purkinje cells[9,10]. | 12:111598856-111650079 | • Alcohol consumption quality<br>• Alcohol drinking<br>• Alzheimer disease<br>• Family history of Alzheimer's disease<br>• Neuroimaging measurement | • 2-aminooctanoate measurement<br>• Asthma<br>• Base metabolic rate measurement<br>• Basophil count<br>• BMI-adjusted waist-hip ratio<br>• Blood protein amount<br>• Body height<br>• Body mass index<br>• Breast carcinoma<br>• Cancer<br>• Childhood onset asthma<br>• Colorectal cancer<br>• Coronary artery disease<br>• Diastolic blood pressure<br>• Eosinophil count<br>• Erythrocyte volume<br>• Estrogen-receptor negative breast cancer<br>• Fatty acid amount<br>• HbA1c measurement<br>• Hematocrit<br>• Hematological measurement<br>• Hemoglobin measurement<br>• Hemolysis<br>• High density lipoprotein cholesterol measurement<br>• Intraocular pressure measurement<br>• Large artery stroke |

| | | | | |
|---|---|---|---|---|
| | | | | • Linoleic acid measurement<br>• Low density lipoprotein cholesterol measurement<br>• Lung adenocarcinoma<br>• Lung carcinoma<br>• Lymphocyte count<br>• Mean arterial pressure<br>• Myocardial infarction<br>• Neutrophil percentage of leukocytes<br>• Omega-6 polyunsaturated fatty acid measurement<br>• Ovarian carcinoma<br>• Ovarian endometrioid carcinoma<br>• Ovarian serous carcinoma<br>• Parental longevity<br>• Platelet count<br>• Platelet glycoprotein Ib alpha chain level<br>• Polyunsaturated fatty acid measurement<br>• Primary biliary cirrhosis<br>• Prostate carcinoma<br>• Psoriasis<br>• Sex hormone-binding globulin measurement<br>• Small vessel stroke<br>• Sphingomyelin measurement<br>• Squamous cell lung carcinoma |

| | | | | |
|---|---|---|---|---|
| | | | | • Stroke<br>• Systolic blood pressure<br>• Tetralogy of fallot<br>• Total cholesterol measurement<br>• Triglycerides:total lipids ratio<br>• Uterine leiomyoma<br>• Whole body water mass |
| 11. *ATXN8OS* (human) | Antisense to the *KLHL1* protein-coding gene, which is likely involved in actin cytoskeleton organisation in neurons[11].<br>CTG expansion causes Spinocerebellar Ataxia Type 8[11,12]. | 13:70107213-70171738 | • Cognitive decline measurement<br>• Dementia<br>• Educational attainment<br>• Information processing speed<br>• Language measurement<br>• Major depressive disorder<br>• Memory performance<br>• Neurofibrillary tangles measurement<br>• Schizophrenia<br>• Word reading | • Age at menopause<br>• Coronary artery disease<br>• COVID-19<br>• Platelet reactivity efficacy<br>• Pneumonia<br>• Protein measurement<br>• Response to clopidogrel<br>• Susceptibility to strep throat |

| | | | | |
|---|---|---|---|---|
| 12. *Bace1-AS* (mouse) | Elevated in AD, regulates/stabilises *BACE1* mRNA and subsequently BACE1 protein expression *in vitro* and *in vivo*[13].<br><br>Knockdown reduces insoluble beta-amyloid and improves memory and learning in AD disease mouse models[14]. | 11:117288453-117293494<br><br>*BACE1-AS* | • None | • Apolipoprotein a 1 measurement<br>• CLMP/TGFBR2 protein level ratio in blood<br>• Fatty acid amount<br>• TGFBR2/TNFRSF1A protein level ratio in blood<br>• Triglyceride measurement |

| | | | | |
|---|---|---|---|---|
| 13. *Bdnf-AS* (mouse, rat) | Represses *BDNF* expression[15].<br>Interferes with the inhibitory learning that underpins extinction of nicotine-seeking and predisposes animals to drug relapse[16]. Implicated in epigenetic reprogramming in the amygdala of early onset alcoholics[17].<br>Knockdown induces neuronal outgrowth and differentiation[15].<br>Knockdown in the infralimbic cortex during extinction training weakens the reinstatement of nicotine-seeking without affecting behaviour during abstinence[16]. | 11:27506808-27698231<br>*BDNF-AS* | • Alcohol consumption quality<br>• Alcohol use disorder measurement<br>• Anxiety measurement<br>• Attention deficit hyperactivity disorder<br>• Bitter alcoholic beverage consumption measurement<br>• Brain attribute<br>• Brain-derived neurotrophic factor measurement<br>• Cannabis dependence<br>• Chronic pain<br>• Chronotype measurement<br>• Cigarettes per day measurement<br>• Coffee consumption measurement<br>• Feeling miserable measurement<br>• Feeling nervous measurement<br>• Mathematical ability | • Abdominal aortic aneurysm<br>• Abnormality of the skeletal system<br>• Age at menarche<br>• Age at menopause<br>• Aspartate aminotransferase measurement<br>• Base metabolic rate measurement<br>• Biological sex<br>• Blood sodium bicarbonate amount<br>• Blood VLDL cholesterol amount<br>• Body fat percentage<br>• Body height<br>• Body mass index<br>• Body surface area<br>• Body weight<br>• Bone density<br>• Chloride amount<br>• Cholesterol:total lipids ratio<br>• Cholesteryl ester measurement<br>• Cholesteryl esters:total lipids ratio<br>• Chylomicron amount<br>• Coronary artery disease<br>• COVID-19<br>• C-reactive Protein measurement |

| | | | | |
|---|---|---|---|---|
| | | | • Neuroimaging measurement<br>• Postural instability<br>• Restless legs syndrome<br>• Risk-taking behaviour<br>• Schizophrenia<br>• Self-reported educational attainment<br>• Smoking behavior<br>• Smoking behavior trait<br>• Smoking cessation<br>• Smoking initiation<br>• Smoking status measurement<br>• Social inhibition quality<br>• Substance abuse<br>• Tea consumption measurement | • Degree of unsaturation measurement<br>• Diastolic blood pressure<br>• Diet measurement<br>• Diverticular disease<br>• Docosahexaenoic acid to total fatty acids percentage<br>• Eosinophil count<br>• Eosinophil percentage of leukocytes<br>• Fat pad mass<br>• Fatty acid amount<br>• Free cholesterol measurement<br>• Free cholesterol:total lipids ratio<br>• Gout<br>• Grip strength measurement<br>• Hematocrit<br>• Hemoglobin measurement<br>• High density lipoprotein cholesterol measurement<br>• Hip circumference<br>• Inflammatory biomarker measurement<br>• Leukocyte quantity<br>• Linoleic acid measurement<br>• Lipid measurement<br>• Low density lipoprotein cholesterol measurement |

| | | | | |
|---|---|---|---|---|
| | | | | • Low density lipoprotein triglyceride measurement<br>• Metabolic syndrome<br>• Morbid obesity<br>• Obesity<br>• Omega-6 polyunsaturated fatty acid measurement<br>• Osteoarthritis<br>• Overall survival<br>• Overnutrition<br>• Pancreatic carcinoma<br>• Phospholipid amount<br>• Phospholipid amount,triglyceride measurement<br>• Phospholipids:total lipids ratio<br>• Physical activity measurement<br>• Polyunsaturated fatty acids to monounsaturated fatty acids ratio<br>• Polyunsaturated fatty acids to total fatty acids percentage<br>• Puberty onset measurement<br>• Saturated fatty acids to total fatty acids percentage<br>• Serum alanine aminotransferase amount |

| | | | | |
|---|---|---|---|---|
| | | | | • Sex hormone-binding globulin measurement<br>• Sexual dimorphism measurement<br>• Sleep apnea<br>• Snoring measurement<br>• Sodium measurement<br>• Systolic blood pressure<br>• Total cholesterol measurement<br>• Triglyceride measurement<br>• Triglycerides in IDL measurement<br>• Triglycerides:total lipids ratio<br>• Type 2 diabetes mellitus<br>• Urate measurement<br>• Uric acid measurement<br>• Urinary albumin to creatinine ratio<br>• Visceral adipose tissue quantity<br>• Waist circumference<br>• Waist-hip ratio<br>• Whole body water mass |

| | | | | |
|---|---|---|---|---|
| 14. *Brd3os / ncRNA_W* (mouse) | High expression pattern during the early stages of neuronal reprogramming, as well as in postnatal mouse and human brain[1]. Overlaps with a CNV morbidity map locus for neurodevelopmental disorders and craniofacial congenital malformations[1]. | 9:134025476-134031587<br><br>*BRD3OS / LINC00094 / h.ncRNA_D* | • None | • Aspartate aminotransferase measurement<br>• C-reactive protein measurement<br>• CD84/SEMA4D protein level ratio in blood<br>• Central corneal thickness<br>• CLIP2/LAT2 protein level ratio in blood<br>• Corneal resistance factor<br>• Eosinophil count<br>• Eosinophil measurement<br>• Free androgen index<br>• Granulocyte percentage of myeloid white cells<br>• HEXIM1/IPCEF1 protein level ratio in blood<br>• LAT2/YES1 protein level ratio in blood<br>• Leukocyte quantity<br>• Myeloid leukocyte count<br>• Neutrophil count<br>• PDGFA/VEGFC protein level ratio in blood<br>• PDGFB/VEGFC protein level ratio in blood<br>• Platelet component distribution width<br>• Platelet count<br>• Platelet crit<br>• Platelet volume |

| | | | | |
|---|---|---|---|---|
| | | | | • Red cell distribution width<br>• Reticulocyte amount<br>• S-6-hydroxywarfarin measurement<br>• Sex hormone-binding globulin measurement<br>• Type 2 diabetes mellitus |

| | | | | |
|---|---|---|---|---|
| 15. *BS-DRL1* (mouse) | Modulates DNA damage response and genome stability in neurons[18].<br><br>Knockout mice exhibit cell-type specific impairment of DNA damage response in the cortex and cerebellum and Purkinje cell degeneration[18]. | 15:89361579-89424983<br><br>*MIR9-3HG* | • Alcohol consumption<br>• Alcohol use disorder measurement<br>• Anxiety measurement<br>• Attention deficit hyperactivity disorder<br>• Bitter alcoholic beverage consumption measurement<br>• Brain attribute<br>• Cannabis dependence<br>• Chronic pain<br>• Cigarettes per day measurement<br>• Coffee consumption measurement<br>• Dopamine beta-hydroxylase measurement<br>• Feeling miserable measurement<br>• Feeling nervous measurement<br>• Mathematical ability<br>• Neuroimaging measurement<br>• Restless legs syndrome | • Abnormality of the skeletal system<br>• Acute myeloid leukemia<br>• Appendicular lean mass<br>• Base metabolic rate measurement<br>• Biological sex<br>• Body height<br>• Body mass index<br>• Body weight<br>• Comparative body size at age 10<br>• Diet measurement<br>• Fat pad mass<br>• Glioma pathogenesis-related protein 1 measurement<br>• IgF-1 measurement<br>• Lean body mass<br>• Level of phosphatidylethanolamine<br>• Pulse pressure measurement<br>• Retinal drusen<br>• Whole body water mass |

| | | | |
|---|---|---|---|
| | | | • Risk-taking behaviour<br>• Schizophrenia<br>• Self-reported educational attainment<br>• Smoking behavior<br>• Smoking cessation<br>• Smoking initiation<br>• Smoking status<br>• Social inhibition quality<br>• Substance abuse<br>• Tea consumption measurement |

| | | | | |
|---|---|---|---|---|
| 16. *C130071C03Riken* (mouse) | Represses neural differentiation of mouse embryonic stem cells.<br><br>Knockdown of splice variants *Rik-201* and *Rik-203* inhibits neural differentiation *in vitro*.<br><br>Knockout of *Rik-201* and *Rik-203 in vivo* causes retardation of embryonic brain development[19]. | 5:88408982-88691313<br><br>*MIR9-2HG* | • ADHD symptom measurement<br>• Age at initiation of smoking<br>• Aggressive behavior quality<br>• Alcohol consumption quality<br>• Alzheimer disease<br>• Anorexia nervosa<br>• Anxiety<br>• Anxiety measurement<br>• Attention deficit hyperactivity disorder<br>• Autism spectrum disorder<br>• Autosomal dominant compelling helio-ophthalmic outburst syndrome<br>• Bipolar disorder<br>• Brain volume<br>• Cannabis dependence<br>• Cannabis use<br>• Caudate nucleus volume<br>• Cerebral cortex area attribute | • Abnormality of refraction<br>• Age at diagnosis<br>• Age at first sexual intercourse measurement<br>• Age at menarche<br>• Age at onset<br>• Agents acting on the renin-angiotensin system use measurement<br>• Age-related macular degeneration<br>• Aging<br>• Biological sex<br>• Body fat percentage<br>• Body mass index<br>• Body weight<br>• Bone density<br>• Cardiovascular disease<br>• Complex trait<br>• COVID-19<br>• Creatinine clearance measurement<br>• Cup-to-disc ratio measurement<br>• Cystatin c measurement<br>• Diastolic blood pressure<br>• Diet measurement<br>• Docosahexaenoic acid measurement<br>• Drugs used in diabetes use measurement |

| | | | | |
|---|---|---|---|---|
| | | | • Chronotype measurement<br>• Cigarettes per day measurement<br>• Coffee consumption measurement<br>• Cognitive function measurement<br>• Conscientiousness measurement<br>• Depressive symptom measurement<br>• Educational attainment<br>• Feeling miserable measurement<br>• Feeling tense measurement<br>• Ganglion thickness<br>• Household income<br>• Income<br>• Insomnia<br>• Intelligence<br>• Irritability measurement<br>• Major depressive disorder<br>• Mathematical ability<br>• Mood disorder | • Erythrocyte count<br>• Essential hypertension<br>• Eye measurement<br>• Fat pad mass<br>• Fatty acid amount<br>• Glaucoma<br>• Glomerular filtration rate<br>• Glucose measurement<br>• Handedness<br>• HbA1c measurement<br>• Health study participation<br>• Hearing loss<br>• Hemoglobin A1 measurement<br>• Hypertension<br>• Lean body mass<br>• Leukocyte quantity<br>• Level of Phosphatidylethanolamine (18:1_0:0) in blood serum<br>• Macula attribute<br>• Macular telangiectasia type 2<br>• Mean arterial pressure<br>• Metabolic syndrome<br>• Morbid obesity<br>• Myopia<br>• Neutrophil count<br>• Open-angle glaucoma<br>• Optic disc size trait<br>• Photoreceptor cell layer thickness measurement |

| | | | | |
|---|---|---|---|---|
| | | | • Neuroimaging measurement<br>• Neuroticism measurement<br>• Obsessive-compulsive disorder<br>• Putamen volume<br>• Risk-taking behaviour<br>• Schizophrenia<br>• Self-reported educational attainment<br>• Sensorineural hearing impairment<br>• Smoking behavior<br>• Smoking behavior trait<br>• Smoking cessation<br>• Smoking initiation<br>• Smoking status measurement<br>• Social inhibition quality<br>• Social interaction measurement<br>• Substance abuse<br>• Substance-related disorder<br>• Tourette syndrome | • Physical activity measurement<br>• Platelet count<br>• Refractive error<br>• Response to lithium ion<br>• Retinal layer thickness<br>• Retinal vasculature measurement<br>• Retinopathy<br>• Retinoschisis<br>• Rheumatoid arthritis<br>• Sunburn<br>• Systolic blood pressure<br>• Taste liking measurement<br>• Trait in response to tenofovir (anhydrous)<br>• Type 2 diabetes mellitus<br>• Vaccination<br>• Visceral adipose tissue quantity<br>• Vitamin D amount<br>• Waist-hip ratio |

| | | | | |
|---|---|---|---|---|
| | | | • Wellbeing measurement<br>• Worry measurement | |
| 17. *C2dat1* (rat, mouse) | Promotes neuronal survival by regulating CaMKIIδ expression after cerebral ischemia[20]. | 4:113504052-113506339<br>Overlaps an exon of *CAMK2D* | • Artificial cardiac pacemaker<br>• Electrocardiography<br>• PR interval<br>• QT interval | • Humerus fracture |
| 18. *Carip / Lhx5as1* (mouse) | Functions as a scaffold, interacts with CaMKIIβ and regulates the phosphorylation of AMPA NMDA receptor subunits in the hippocampus.<br>Knockout causes dysfunction of synaptic transmission and attenuates long term potentiation in hippocampal CA3-CA1 synapses and impairs spatial learning and memory[21].<br>GWAS associated with Alzheimer's disease risk[22]. | 12:113467570-113480636<br>*LHX5-AS1* | • Cerebrospinal fluid composition attribute<br>• Cognitive function measurement<br>• Mathematical ability<br>• PR interval<br>• Self-reported educational attainment | • Acute myeloid leukemia<br>• Body mass index<br>• C-reactive protein measurement<br>• High density lipoprotein cholesterol measurement<br>• LDL peak particle diameter measurement<br>• Protein measurement<br>• Total fat intake measurement<br>• Triglyceride measurement |

| | | | | |
|---|---|---|---|---|
| 19. *CDR1as / ciRS-7* (mouse) | Binds *miR-7* in neural tissues[23].<br>Reported to control miR-7 availability to influence immediate early gene expression, synaptic vesicle release and sensorimotor gating behaviour[24].<br>Expression of the human ortholog of *CDR1as* in zebrafish impairs midbrain development[23].<br>Deficiency leads to dysfunction of excitatory synaptic transmission[24].<br>Knockdown impairs fear extinction memory[25]. | X:140709562-140793215<br>*LINC00632* | • Pack-years measurement | • Esophageal adenocarcinoma |
| 20. *CHASERR* (human) | A highly conserved lncRNA adjacent to *CHD2*–a coding gene in which loss-of-function variants cause developmental and epileptic encephalopathy.<br>*Chaserr* deletion causes severe encephalopathy, facial dysmorphisms, cortical atrophy, and cerebral hypomyelination[26]. | 15:92819305-92899701 | • Educational attainment<br>• Wellbeing measurement | • Body height<br>• Body mass index<br>• Glioma pathogenesis-related protein 1 measurement<br>• Serum creatinine amount<br>• Uric acid measurement |

| | | | | |
|---|---|---|---|---|
| 21. *Cyrano* (mouse, zebrafish) | Required for proper embryonic development of the nervous system in zebrafish[34].<br>Targets *mir7* for degradation and forms a regulatory network with *CDRas*[35].<br>Loss of function in zebrafish embryos using morpholino antisense oligos causes small heads and eyes, and short, curly tails, defects in neural tube opening, loss of NeuroD-positive neurons in the retina and tectum, and enlarged nasal placodes[34].<br>Rescued by human/mouse orthologs.<br>Deletion in mouse does not cause overt developmental defects but alters regulatory circuitry in neurons[35]. | 15:41283958-41322392<br>*OIP5-AS1* | • Cognitive function measurement<br>• Self-reported educational attainment | • COVID-19<br>• Lymphocyte amount<br>• Neutrophil measurement<br>• Systolic blood pressure<br>• Tyrosine-protein kinase receptor TYRO3 measurement |

| | | | | |
|---|---|---|---|---|
| 22. *Dali* (mouse) | Transcribed from an enhancer downstream of the *Pou3f3* transcription factor gene.<br>Epigenetically regulates transcription of the *Pou3f3* locus.<br>Interacts with the DNMT1 DNA methyltransferase and regulates DNA methylation of promoters of nearby and distal genes involved in neural differentiation.<br>Depletion of *Dali* disrupts the differentiation of neuroblastoma cells[36]. | 2:104853775-104927577<br>*ENSG00000269707* | • Amygdala volume<br>• Brain attribute<br>• Cerebral cortex area attribute<br>• Cortical thickness<br>• Hippocampal volume<br>• Memory performance<br>• Neuroimaging measurement | • Abnormality of the skeletal system<br>• BMI-adjusted waist circumference<br>• Body height<br>• Body mass index<br>• Vital capacity |
| 23. *Dlx1as* (mouse) | Antisense to and negative regulator of the *Dlx1* homeobox gene[37,38].<br>Ablation results in mild skeletal and neurological phenotypes. Dysregulates interneuron development[37]. | 2:172081990-172093647<br>*ENSG00000288958* | • Schizophrenia | • None |

| | | | | |
|---|---|---|---|---|
| 24. *Durga* / *Kalnc2* (mouse) | Regulates *Kalrn* expression and modulates dendrite density[39].<br>*Kalnc2* is the mammalian ortholog[40].<br>Ectopic expression in mouse neuronal cells leads to downregulation of the major *Kalrn* mRNA isoforms[40].<br>Neurons cultured from *durga* injected zebrafish embryos have significantly fewer and shorter dendrites[39]. | 3:124094661-124164643<br>*hsKALNC2* *(KALRN* isoforms *ENST000004888 25.5* and *ENST000006844 41.1*) | • Cognitive function measurement<br>• Educational attainment<br>• Neurofibrillary tangles measurement<br>• PR interval<br>• Self-reported educational attainment | • Body height<br>• Breastfeeding duration<br>• Gut microbiome measurement<br>• Spontaneous preterm birth |

| | | | | |
|---|---|---|---|---|
| 25. *Emx2OS* (mouse) | Regulates the expression of *EMX2*, a homeobox transcription factor implicated in several aspects of cerebral cortex development.<br><br>Knockdown of *Emx2OS* increases *Emx2* expression.<br><br>When delivered to rhombo-spinal precursors, stimulates ectopic expression of *Emx2*.<br><br>*Emx2* knock-out dramatically impairs *Emx2OS* transcription[41]. | 10:117473213-117545068<br><br>*EMX2OS* | • Alzheimer disease<br>• Anxiety measurement<br>• Autosomal dominant compelling helio-ophthalmic outburst syndrome<br>• Bipolar disorder<br>• Brain attribute<br>• Cerebral cortex area attribute<br>• Chronotype measurement<br>• Cortical thickness<br>• Depressive symptom measurement<br>• Family history of Alzheimer's disease<br>• Neuroimaging measurement<br>• Neuroticism measurement<br>• Obsessive-compulsive disorder<br>• PHF-tau measurement<br>• Schizophrenia<br>• Schizophrenia symptom severity measurement | • Allergen exposure measurement<br>• Balding measurement<br>• BMI-adjusted waist-hip ratio<br>• BMI-adjusted waist circumference<br>• Body height<br>• Bone density breast carcinoma<br>• Cartilage disease<br>• Early cardiac repolarization measurement<br>• Eye morphology trait<br>• Gut microbiome measurement<br>• Hip geometry<br>• Lobe attachment<br>• Peripheral arterial disease<br>• Pit and fissure surface dental caries<br>• Pulmonary function measurement<br>• Traffic air pollution measurement<br>• Trait in response to paliperidone<br>• Urate measurement<br>• Waist-hip ratio<br>• X-24334 measurement |

| | | | | |
|---|---|---|---|---|
| | | | • Wellbeing measurement | |
| 26. *Evf2 / Dlx6os1 / Dlx6as* (mouse) | Antisense to *Dlx6* homeobox gene.<br>Regulates interneuron subtype genes by recruiting Dlx and Mecp2 transcription factors.<br>Required for the formation GABAergic interneuronal circuitry[42,43].<br>*Evf2* mouse mutants have reduced GABAergic interneurons in early postnatal hippocampus and dentate gyrus. GABAergic interneurons return to normal in *Evf2* mutant adult hippocampus, but have reduced synaptic inhibition[42].<br>Mice lacking *Evf2* have increased seizure susceptibility and severity[43]. | 7:96955141-97014088<br>*DLX6-AS1* | • Attention deficit hyperactivity disorder<br>• Behcet's syndrome<br>• Educational attainment<br>• Intelligence<br>• PHF-tau measurement<br>• Self-reported educational attainment<br>• Smoking behavior<br>• Smoking behavior trait<br>• Smoking initiation<br>• Smoking status measurement<br>• Social inhibition quality<br>• Substance abuse | • Alkaline phosphatase measurement<br>• Anthropometric measurement<br>• Body height<br>• Cardioembolic stroke<br>• Gastroesophageal reflux disease<br>• Heel bone mineral density<br>• Hemoglobin measurement<br>• Ossification of the posterior longitudinal ligament of the spine<br>• Osteoarthritis<br>• Response to radiation<br>• Sodium measurement<br>• Total hip arthroplasty |

| | | | | |
|---|---|---|---|---|
| 27. *Fedora / CEDORA / RP11-298D21.1* (human) | Sex-specific regulation of depression.<br>Increased in the prefrontal cortex of depressed females only.<br>In female mice, neuronal ectopic expression induced anxiety and depression-like behaviours, while oligodendrocyte expression impaired reward learning and performance[44]. | 16:83706000-83772974 | • Cognitive function measurement<br>• Hippocampal volume<br>• Neuropsychological test<br>• PHF-tau measurement | • Allergen exposure measurement<br>• Breastfeeding duration<br>• Chemotherapy-induced cytotoxicity measurement<br>• Diastolic blood pressure<br>• Gut microbiome measurement<br>• Level of heat shock factor-binding protein 1 in blood<br>• Level of protein BRICK1 in blood<br>• Level of WASH complex subunit 3 in blood<br>• Neutropenia<br>• Periodontitis<br>• Response to platinum-based neoadjuvant chemotherapy<br>• Serum IgG glycosylation measurement<br>• Systolic blood pressure |

| | | | | |
|---|---|---|---|---|
| 28. *FILNC1* (FOXO induced long non-coding RNA 1) (human) | | 6:139677639-139860476 | • Alzheimer disease<br>• Brain attribute<br>• Caudate nucleus volume<br>• Cerebral cortex area attribute<br>• Cognitive function measurement<br>• Dementia<br>• PHF-tau measurement<br>• Response to trauma exposure<br>• Schizophrenia<br>• Smoking initiation | • 2-naphthol sulfate measurement<br>• 5-acetylamino-6-amino-3-methyluracil measurement<br>• Age at onset<br>• Appendicular lean mass<br>• Atrial fibrillation<br>• Birth weight<br>• Blood copper level<br>• Blood immunoglobulin amount<br>• Blood VLDL cholesterol amount<br>• BMI-adjusted waist circumference<br>• BMI-adjusted waist-hip ratio<br>• Body height<br>• Bone density<br>• Chronic obstructive pulmonary disease<br>• Chylomicron amount<br>• Diet measurement<br>• Docosahexaenoic acid measurement<br>• Erythrocyte volume<br>• FEV/FVC ratio<br>• Forced expiratory volume<br>• Free cholesterol measurement<br>• Glomerular filtration rate |

| | | | | |
|---|---|---|---|---|
| | | | | • Gut microbiome measurement<br>• Health trait<br>• Heel bone mineral density<br>• Lean body mass<br>• Lipid measurement<br>• Mean corpuscular hemoglobin<br>• Mean reticulocyte volume<br>• Metabolic syndrome<br>• Monocyte count<br>• Monocyte percentage of leukocytes<br>• Omega-3 polyunsaturated fatty acid measurement<br>• PCSK9 protein measurement<br>• Peak expiratory flow<br>• Phospholipid amount<br>• Protein measurement<br>• Red cell distribution width<br>• Response to simvastatin<br>• Reticulocyte count<br>• Serum IgA amount<br>• Serum IgG glycosylation measurement<br>• Size<br>• Thyroid stimulating hormone amount<br>• Triglyceride measurement<br>• Type 2 diabetes mellitus<br>• Urate measurement |

| | | | | |
|---|---|---|---|---|
| | | | | • Visceral:abdominal adipose tissue ratio measurement<br>• Vital capacity |
| 29. *Firre* (mouse) | Implicated in Poly microgia[45,46]. | X:131688584-131830928<br><br>*FIRRE / LINC01200* | • None | • None |
| 30. *FosDT* (rat) | Exacerbates ischemic brain injury[47,48].<br><br>*FosDT* knockdown ameliorates postischemic motor deficits and reduces the infarct volume after middle cerebral artery occlusion[47].<br><br>*FosDT* knockout rats show enhanced sensorimotor recovery and reduced brain damage after middle cerebral artery occlusion[48]. | 14:75210494-75279526<br><br>*ENSG00000258740* | • Smoking initiation | • Ankylosing spondylitis<br>• Body fat percentage<br>• Body height<br>• C-reactive protein measurement<br>• Crohn's disease<br>• Inflammatory bowel disease<br>• Psoriasis<br>• Sclerosing cholangitis<br>• Trait in response to apixaban<br>• Ulcerative colitis<br>• Vaginal microbiome measurement |

| | | | | |
|---|---|---|---|---|
| 31. *Gas5* (mouse, rat) | Promotes hippocampal neural stem cell differentiation into neurons[49].<br><br>Regulates the activity-dependent trafficking and clustering of synaptic RNA condensates[49,50].<br><br>Essential for fear extinction[50], motivated behaviour[51] and spatial memory recovery[49].<br><br>Overexpression restores learning in rats with cholinergic injury[49], while overexpression in nucleus accumbens reduces cocaine intake and facilitates extinction[51].<br><br>Synapse-specific knockdown impairs fear extinction[50].<br><br>Knockdown increases the trafficking of RNA granules and alters the pattern of clustering and assembly, regulating intrinsic excitability[50]. | 1:173851424-173868940<br><br>*GAS5* | • Intelligence | • Body height<br>• Erythrocyte count<br>• Hematocrit<br>• Hemoglobin measurement<br>• Level of tenascin-N in blood<br>• Red blood cell density |

| | | | | |
|---|---|---|---|---|
| 32. *Gm12371* (mouse) | Regulates synaptic transmission, synapse density, dendritic arborization and expression of several genes involved in neuronal growth and development.<br><br>Knockdown results in a decrease in synaptic transmission, total synapse density, number of mushroom spines, and dendritic arborization[52]. | 9:29213251-29229299<br><br>*ENSG00000287038* | • None | • None |
| 33. *Gm16277 / ENSMUSG00000144135 / ncRNA_V* (mouse) | High expression pattern during the early stages of neuronal reprogramming, as well as in postnatal and adult mouse brain[1]. | 10:116024972-116062503<br><br>None | • None | • Joint damage measurement<br>• Response to TNF antagonist |

| | | | | |
|---|---|---|---|---|
| 34. *Gm21284 / LOC120102321* (rat) | Promotes neural stem cell differentiation toward cholinergic neurons by competitive inhibition of *miR-30e-3P* and subsequent regulation of *Lhx8* expression.<br>Proliferation of neural stem cells is repressed after overexpression[53]. | 2:73823135-73856957<br>*ENSG00000287250* and *ENSG00000293671* | • None | • 2-aminooctanoate measurement<br>• ABHD14B/STAMBP protein level ratio in blood<br>• Amino acid measurement<br>• Breastfeeding duration<br>• CC2D1A/STAMBP protein level ratio in blood<br>• CRADD/STAMBP protein level ratio in blood<br>• DARS1/STAMBP protein level ratio in blood<br>• Gut microbiome measurement<br>• IgF-1 measurement<br>• Level of STAM-binding protein in blood<br>• N-acetylarginine measurement<br>• N-delta-acetylornithine measurement<br>• Pregnancy disorder<br>• RWDD1/STAMBP protein level ratio in blood<br>• Serum metabolite level<br>• SNX9/STAMBP protein level ratio in blood<br>• STAMBP/WWP2 protein level ratio in blood |

| | | | | |
|---|---|---|---|---|
| 35. *HTT-AS* / *ENSMUSG00000140667* (mouse) | Antisense to repeat in huntington (HTT) protein and reduced in HD brains. Negatively regulates *HTT* expression.<br><br>Overexpression reduces endogenous *HTT*, while knockdown increases *HTT*[1]. | 4:3046139-3074624<br><br>*HTT-AS* | • None | • Aspartate aminotransferase measurement<br>• Body fat percentage<br>• Gastric cancer<br>• Glomerular filtration rate<br>• Hepatocyte growth factor activator amount<br>• High density lipoprotein cholesterol measurement<br>• Low density lipoprotein triglyceride measurement<br>• Monocyte count<br>• Myocardial infarction<br>• Neutrophil count<br>• Neutrophil percentage of leukocytes<br>• Serum alanine aminotransferase amount<br>• Sex hormone-binding globulin measurement<br>• Triglyceride measurement |

| | | | | |
|---|---|---|---|---|
| 36. *LINC00473* (human) | Sex-specific regulation of depression.<br><br>Ectopic expression in mouse results in stress resilience and reduces both depression- and anxiety-like behaviours in female mice only[54]. | 6:165327287-165988301 | • Alcohol drinking<br>• Alzheimer disease<br>• Behavioural inhibitory control measurement<br>• Bipolar disorder<br>• Chronotype measurement<br>• Conduct disorder<br>• Economic and social preference<br>• Educational attainment<br>• Family history of Alzheimer's disease<br>• Opioid dependence<br>• PHF-tau measurement<br>• Self-reported educational attainment | • Adolescent idiopathic scoliosis<br>• Agents acting on the renin-angiotensin system use measurement<br>• Age-related nuclear cataract<br>• Antihypertensive use measurement<br>• Appendicular lean mass<br>• Aspartate aminotransferase measurement<br>• Autoimmune thyroid disease<br>• Beta blocking agent use measurement<br>• Birth weight<br>• Blood immunoglobulin amount<br>• Body height<br>• Body mass index<br>• Bone density<br>• Breastfeeding duration<br>• Cardiovascular disease<br>• Chloride amount<br>• Cortisol measurement<br>• COVID-19<br>• C-reactive protein measurement<br>• Cytokine measurement<br>• Diastolic blood pressure<br>• Diastolic blood pressure change measurement |

| | | | | |
|---|---|---|---|---|
| | | | | • Drug use measurement<br>• Environmental exposure measurement<br>• Erythrocyte count<br>• Fetal genotype effect measurement<br>• Fever<br>• Glioblastoma multiforme<br>• Glioma pathogenesis-related protein 1 measurement<br>• Glutamine measurement<br>• Gut microbiome measurement<br>• Heel bone mineral density<br>• Hematocrit<br>• Hemoglobin measurement<br>• High density lipoprotein cholesterol measurement<br>• Hormone measurement<br>• Hypertension<br>• Hyperthyroidism<br>• Hypothyroidism<br>• IgF-1 measurement<br>• Lean body mass<br>• Level of diglyceride<br>• Level of thyrotropin subunit beta in blood<br>• Lipid measurement<br>• Mean arterial pressure<br>• Metabolic syndrome<br>• Oligodendroglioma |

| | | | | |
|---|---|---|---|---|
| | | | | • Parental longevity<br>• Physical activity measurement<br>• Placenta mass<br>• Platelet count<br>• Platelet crit<br>• Platelet reactivity efficacy<br>• Radiation-induced disorder<br>• Red blood cell density<br>• Refractive error<br>• Response to clopidogrel<br>• Response to fenofibrate<br>• Serum metabolite level<br>• Size<br>• Sodium measurement<br>• Stromelysin-2 measurement<br>• Systolic blood pressure<br>• Thyroid disease<br>• Thyroid preparation use measurement<br>• Thyroid stimulating hormone amount<br>• Thyroiditis<br>• Thyrotoxicosis<br>• Trait in response to sulfasalazine<br>• Type 2 diabetes mellitus<br>• Urate measurement<br>• Wheezing<br>• Whole body water mass |

| | | | | |
|---|---|---|---|---|
| 37. *Linc-Brn1b* (mouse) | Important for cortical lamination.<br>Knockout mice exhibit a decrease in proliferation of cortical progenitors *in vivo*, restricted to a specific subpopulation of progenitors in the subventricular zone of developing cortex.<br>Knockout also (consequently) results in reduction in intermediate progenitor cells in the cerebral cortex[55]. | 2:104865401-104874447<br>*LINC01159* | • Amygdala volume<br>• Brain attribute<br>• Cerebral cortex area attribute<br>• Cortical thickness<br>• Hippocampal volume<br>• Memory performance<br>• Neuroimaging measurement | • Body height<br>• Body mass index<br>• Vital capacity |
| 38. *LncND / TCONS_00003533 / TCONS_00003534 / TCONS_00004657 / TCONS_00003535* (human) | *LncND* binding and release of *miR-143-3p* controls expression of Notch receptors. Maintains neural progenitor pool and supports cortical development.<br>Downregulation in neuroblastoma cells reduced proliferation and induced neuronal differentiation. Ectopic expression in developing mouse cortex led to an expansion of PAX6+ radial glial cells[56]. | 2:663814-667014 | • None | • Allergen exposure measurement<br>• Body mass index<br>• Gut microbiome measurement |

| | | | | |
|---|---|---|---|---|
| 39. *lnc-NR2F1 / A830082K12Rik* (mouse) | Regulates neuronal gene transcription and enhances neuronal maturation. Induced during mouse embryonic fibroblast-to-induced neuron cell conversion.<br>Co-expressed with *Ascl1* in mouse embryonic fibroblast increase in the number of TauEGFP positive cells with neurites.<br>Overexpression results in significant increase in neurite length.<br>Knockout down regulates neuronal pathfinding and axon guidance genes.<br>A t(5:12) chromosomal translocation disrupting *Lnc-NR2F1* occurs in a family manifesting neurodevelopmental symptoms[1]. | 5:93360779-93585649<br>*NR2F1-AS1* | • Brain attribute<br>• Brain volume<br>• Cerebral cortex area attribute<br>• Cognitive function measurement<br>• Cortical thickness<br>• Educational attainment<br>• Income<br>• Intelligence<br>• Mathematical ability<br>• Neuroimaging measurement<br>• Self-reported educational attainment<br>• Total cortical area measurement | • Balding measurement<br>• Biological sex<br>• Body height<br>• Body mass index<br>• Bone remodeling disease<br>• C-reactive protein measurement<br>• COVID-19<br>• Diet measurement<br>• Facial morphology trait<br>• Glioma pathogenesis-related protein 1 measurement<br>• Hip circumference<br>• Type 2 diabetes mellitus |

| | | | | |
|---|---|---|---|---|
| 40. *lnc-OPC / Gm41031 / 5330416C01Rik* (mouse) | Regulates oligodendrocyte precursor cell differentiation.<br>Knockdown significant decreases expression of OPC markers during. Neural stem cell differentiation to oligodendrocyte progenitor cells[57]. | 5:73947266-74054445<br>*ENSG00000304504* | • Diffuse plaque measurement<br>• Smoking initiation | • 1,3,7-trimethylurate measurement<br>• Amino acid measurement<br>• C-reactive protein measurement<br>• Chronic graft versus host disease<br>• Cleft lip<br>• Diabetic retinopathy<br>• Donor genotype effect measurement<br>• Vaginal microbiome measurement |

| | | | | |
|---|---|---|---|---|
| 41. *lncRNA_ES1 / LINC01108* (human) | Required for pluripotency.<br>Knockdown resulted in downregulation of pluripotency markers and simultaneous upregulation of lineage markers including neuroectoderm, endoderm and mesoderm germ layers[58]. | 6:14280127-14285454 | • Educational attainment<br>• Intelligence<br>• Mathematical ability<br>• Memory performance<br>• Neurofibrillary tangles measurement<br>• Neuropsychological test<br>• Self-reported educational attainment<br>• Short-term memory<br>• Smoking initiation<br>• Vascular dementia | • BMI-adjusted waist-hip ratio<br>• BMI-adjusted waist circumference<br>• Body height<br>• Caffeic acid measurement<br>• Caffeine metabolite measurement<br>• Cataract<br>• CD83 antigen measurement<br>• Ceramide amount<br>• Cytokine measurement<br>• Dipeptidase 1 measurement<br>• Fatty acid amount<br>• Glioma pathogenesis-related protein 1 measurement<br>• Heart rate<br>• Herpes Zoster<br>• HVA measurement<br>• Lymphoma<br>• MHPG measurement<br>• Ovarian reserve<br>• Parental longevity<br>• Serum metabolite level<br>• Trait in response to tofacitinib<br>• Uric acid measurement<br>• Vaginal microbiome measurement<br>• Waist-hip ratio |

| | | | | |
|---|---|---|---|---|
| 42. *lncRNA_ES2 / ENSG00000282849* (human) | Required for pluripotency.<br>Knockdown resulted in downregulation of pluripotency markers and simultaneous upregulation of lineage markers including neuroectoderm, endoderm and mesoderm germ layers[58]. | 1:200439757-200483604 | • Schizophrenia | • Calcium measurement<br>• Type 2 diabetes mellitus |
| 43. *lncRNA-ES3 / LINC00458* (human) | Required for pluripotency.<br>Knockdown resulted in downregulation of pluripotency markers and simultaneous upregulation of lineage markers including neuroectoderm, endoderm and mesoderm germ layers[58]. | 13:53950255-54270397 | • Insomnia<br>• Insomnia measurement<br>• Memory performance<br>• Smoking initiation | • 3-hydroxy-1-methylpropylmercapturic acid measurement<br>• Body height<br>• Body mass index<br>• Breastfeeding duration<br>• Cardiotoxicity<br>• Creatine amount<br>• DNA methylation<br>• Fat pad mass<br>• Gut microbiome measurement<br>• Heart disease<br>• Metabolic syndrome<br>• Olfactomedin-4 measurement<br>• Protein measurement<br>• Response to trastuzumab<br>• Stroke<br>• Tryptophan measurement<br>• Waist-hip ratio |

| | | | | |
|---|---|---|---|---|
| 44. *lncRNA_N1 / LINC01109 / LINC01111 / ENSG00000270866* (human) | Required for neuronal differentiation from stem cells.<br>Knockdown results in fewer early post-mitotic neurons in culture[58]. | 8:76403559-76524356 | • Cortical thickness<br>• Educational attainment<br>• Mathematical ability<br>• Self-reported educational attainment<br>• Smoking cessation<br>• Smoking initiation | • Anthropometric measurement<br>• Beta blocking agent use measurement<br>• Body height<br>• Body mass index<br>• Diet measurement<br>• Metabolic syndrome<br>• Ovarian serous carcinoma<br>• Susceptibility to chronic sinus infection measurement<br>• Systolic blood pressure |

| | | | | |
|---|---|---|---|---|
| 45. *lncRNA_N2 / MIR100HG* (human) | Required for neuronal differentiation from stem cells.<br><br>Knockdown resulted in fewer early post-mitotic neurons in culture[58]. | 11:122028325-122556721 | • Alcohol consumption quality<br>• Attempted suicide<br>• Brain connectivity attribute<br>• Brain physiology trait<br>• Chronotype measurement<br>• Cognitive function measurement<br>• Depressive symptom measurement<br>• Early-onset Alzheimers disease<br>• Educational attainment<br>• Intelligence<br>• Language measurement<br>• Late-onset Alzheimer's disease<br>• Mathematical ability<br>• Memory performance<br>• Nicotine dependence symptom count<br>• PHF-tau measurement<br>• Schizophrenia | • 1-linoleoyl-2-linolenoyl-GPC (18:2/18:3) measurement<br>• 1-oleoyl-GPC (18:1) measurement<br>• Able to hear with hearing aids<br>• Age at assessment, cardiovascular disease<br>• Astrocytoma<br>• Balding measurement<br>• Body height<br>• Body mass index<br>• Body shape measurement<br>• Bone density<br>• Childhood trauma measurement<br>• Colorectal cancer<br>• Corneal endothelial cell attribute<br>• Corneal resistance factor<br>• Diet measurement<br>• Erythrocyte attribute<br>• Eye morphology trait<br>• Facial morphology trait<br>• FEV/FVC ratio<br>• Gene expression attribute<br>• Glioma<br>• Glioma pathogenesis-related protein 1 measurement<br>• Gut microbiome measurement |

| | | | | |
|---|---|---|---|---|
| | | | • Schizophrenia symptom severity measurement<br>• Self-reported educational attainment<br>• Smoking initiation<br>• Smoking status measurement<br>• Trait in response to paliperidone<br>• Visual masking measurement | • Heel bone mineral density<br>• High density lipoprotein cholesterol measurement<br>• Low density lipoprotein cholesterol measurement<br>• Metabolic syndrome<br>• Oligodendroglioma<br>• Pathological myopia<br>• Platelet aggregation<br>• Platelet crit<br>• Response to bronchodilator<br>• Stroke<br>• Taxonomic microbiome measurement<br>• Triglyceride measurement<br>• Vaginal microbiome measurement<br>• Waist-hip ratio<br>• Waist circumference |
| 46. *lncRNA_N3 / AK055040* (human) | Required for neuronal differentiation from stem cells.<br>Knockdown resulted in fewer early post-mitotic neurons in culture[58]. | 7:81946444-81948479 | • None | • None |

| | | | | |
|---|---|---|---|---|
| 47. *lnc-ZFP238 / LINC02774 / LOC339529* (human) | High expression pattern during the early stages of neuronal reprogramming, as well as in postnatal mouse and human brain.<br><br>Locus is disrupted by two focal CNVs in two distinct ASD/ID patients[1]. | 1:243917316-244047551 | • Attention deficit hyperactivity disorder<br>• Cognitive inhibition measurement<br>• Mathematical ability<br>• PHF-tau measurement<br>• Post-traumatic stress disorder<br>• Risk-taking behaviour<br>• Social inhibition quality<br>• Substance abuse | • Body height<br>• Breast cancer<br>• Breastfeeding duration<br>• Color vision disorder<br>• Colorectal cancer<br>• Diabetic retinopathy<br>• Ghrelin measurement<br>• Gut microbiome measurement<br>• Lean body mass<br>• Monocyte count<br>• Mortality<br>• Myocardial infarction<br>• Response to allogeneic hematopoietic stem cell transplant<br>• Size<br>• Systolic blood pressure<br>• Trait in response to hydrochlorothiazide<br>• Trauma exposure measurement<br>• Triglyceride measurement<br>• Urate measurement |

| | | | | |
|---|---|---|---|---|
| 48. *LoNa / Gm17382* (mouse) | Modulates learning by controlling protein translation.<br>Knockdown enhances learning and long-term memory and restores impaired memory function in APP/PS1 transgenic mice[59]. | 5:173249862-173284273<br>*ENSG00000298841*[b], *ENSG00000289493*[b], *ENSG00000309052*[b] | • None | • Health study participation |
| 49. *Maalin* (mouse) | Regulates the expression of the monoamine oxidase A gene in the brain, is elevated in the dentate gyrus of individuals with impulsive-aggressive suicides.<br>Overexpression in hippocampus reduces monoamine oxidase A levels and promotes impulsive-aggressive behaviour in mice[60]. | X:43746818-43766609<br>*MAALIN* | • Smoking behavior | • None |

[b] Correct strand but downstream of alignment.

| | | | | |
|---|---|---|---|---|
| 50. *Malat1* (mouse) | Regulates hippocampal neurite outgrowth via a *MALAT1/microRNA-30* axis controlling *Spastin* expression[61-63].<br><br>Malat1 ($m^6A$-modified) localises to synapses during fear-extinction learning, where it interacts with the m6A reader DPYSL2 to regulate dendritic spine formation and support memory consolidation[64].<br><br>Knockdown results in defects in neurite outgrowth[61,62] as well as enhanced cell death[61].<br><br>Knockdown decreases while overexpression increases synaptic density in cultured hippocampal neurons[63].<br><br>Synapse-specific and state-dependent reduction of $m^6A$ on *Malat1* impairs fear-extinction memory[64].<br><br>Upregulated in ischemic stroke and plays a protective role by inhibiting apoptosis and inflammation[65]. | 11:65497606-65508073<br><br>*MALAT1* | • Schizophrenia symptom severity measurement<br>• Trait in response to paliperidone | • BMI-adjusted hip circumference<br>• Bone density<br>• Fat pad mass<br>• Femoral neck bone mineral density<br>• Glaucoma<br>• Heart rate<br>• High density lipoprotein cholesterol measurement<br>• Level of latent-transforming growth factor beta-binding protein 3 in blood<br>• Lymphocyte count<br>• Metabolic syndrome<br>• Osteoarthritis<br>• Periodontitis<br>• Sex hormone-binding globulin measurement<br>• Total joint arthroplasty<br>• Triglyceride measurement<br>• Uric acid measurement<br>• Visceral adipose tissue quantity<br>• Waist-hip ratio |

| | | | | |
|---|---|---|---|---|
| 51. *Meg3* (mouse) | Activity induced; modulates AMPA receptor surface expression[66].<br><br>Knockdown enhances surface AMPAR expression and dysregulates the PTEN/PI3K/AKT signalling pathway in neurons[66].<br><br>Upregulated in AD patient temporal gyrus and induces necroptosis in neurons[67].<br><br>Increased in HD cell and animal models. Transient knockdown reduces aggregates formed by mutant huntingtin[68]. | 14:100779206-100861031<br><br>*MEG3* | • None | • 2,6-dihydroxybenzoic acid measurement<br>• Birth weight<br>• Blood cobalt amount<br>• Blood protein amount<br>• Body height<br>• Breast carcinoma<br>• Breastfeeding duration<br>• Chymotrypsinogen B measurement<br>• Delta-like protein 1 measurement<br>• Diastolic BP change measurement<br>• Emphysema imaging measurement<br>• Environmental exposure measurement<br>• Glucose measurement<br>• Gut microbiome measurement<br>• HbA1c measurement<br>• HETE measurement<br>• IgF-1 measurement<br>• Inactive pancreatic lipase-related protein 1 measurement<br>• Lean body mass<br>• Level of phospholipase A2 in blood<br>• Optic disc size trait |

| | | | | |
|---|---|---|---|---|
| | | | | • Peripheral arterial disease<br>• Protein delta homolog 1 measurement<br>• Response to diuretic<br>• Size<br>• Traffic air pollution measurement<br>• Trypsin-2 measurement<br>• Type 1 diabetes mellitus<br>• Type 2 diabetes mellitus |

| | | | | |
|---|---|---|---|---|
| 52. *Megamind / Tuna / TUNAR* (mouse, zebrafish) | Regulates brain morphogenesis and eye development[34] and is required for neural lineage commitment, differentiation and function. Interacts with the RNA-binding proteins PTBP1, hnRNP-K, and nucleolin[69].<br><br>Also encodes a micro-peptide that is conserved in vertebrates[70].<br><br>Morpholino antisense oligos produce defects in brain-ventricle morphology such as an unusual expansion of the midbrain ventricle, loss of the midbrain hingepoint, contraction of the forebrain ventricle. Morphants also have smaller heads and eyes, enlarged brain ventricles, and loss of NeuroD-positive neurons in the retina and tectum[34].<br><br>Depletion causes impaired cell proliferation; overexpression is associated with elevated levels of proliferation[69]. | 14:95866587-95925663<br><br>*TUNAR* | • Short-term memory | • Atypical femoral fracture<br>• Basophil measurement<br>• Bone density<br>• Chromosome telomeric region length<br>• Clonal hematopoiesis<br>• Drug use measurement<br>• Emphysema imaging measurement<br>• Endometriosis<br>• Gut microbiome measurement<br>• Level of T-cell leukemia/lymphoma protein 1A in blood<br>• Lymphocyte count<br>• Mosaic loss of chromosome Y measurement<br>• Myeloproliferative disorder<br>• Neutrophil count<br>• Platelet count<br>• Platelet crit<br>• Protein measurement<br>• Response to bisphosphonate<br>• Retinal vasculature measurement |

| | |
|---|---|
| | Knockdown blocks neural differentiation in H9 human embryonic stem cells[69].<br><br>Knockdown inhibits neural differentiation of mouse embryonic stem cells[70].<br><br>*TUNAR* micropeptide (pTUNAR) deficiency in mouse embryonic stem cells improves their differentiation potential towards the neural lineage.<br><br>pTUNAR overexpression impairs neuronal differentiation by reduced neurite formation[70]. |

| | | | | |
|---|---|---|---|---|
| 53. *Meteor* (mouse) | Transcription elongation through the *Meteor* locus is required for *Eomes* activation in mouse embryonic stem cells. *Meteor* is suppressed in neuronal differentiation of embryonic stem cells[71]. | 3:27796694-27931579<br><br>*LINC01980* | • Amyloid-beta measurement<br>• Depressive symptom measurement<br>• Educational attainment<br>• Lifestyle measurement<br>• Maximum cigarettes per day measurement<br>• Multiple sclerosis<br>• Neuritic plaque measurement<br>• Response to antipsychotic drug<br>• Response to selective serotonin reuptake inhibitor | • Ankylosing spondylitis<br>• Anti-citrullinated protein antibody seropositivity<br>• Basophil count<br>• Cancer<br>• Chronic lymphocytic leukemia<br>• Common variable immunodeficiency<br>• Eosinophil count<br>• Eosinophil percentage of leukocytes<br>• Hemoglobin A1 measurement<br>• Hodgkins lymphoma<br>• Leukocyte quantity<br>• Lymphocyte count<br>• Multiple myeloma<br>• Non-Hodgkins lymphoma<br>• Platelet count<br>• Platelet crit<br>• PR interval<br>• Response to stimulus<br>• Rheumatoid arthritis<br>• Rheumatoid factor seropositivity measurement<br>• RS-warfarin measurement<br>• Tumor necrosis factor alpha amount<br>• Triglyceride change measurement |

| | | | | |
|---|---|---|---|---|
| | | | | • Vaginal microbiome measurement |

| | | | | |
|---|---|---|---|---|
| 54. *Miat / Gomafu / RNCR2* (mouse) | Knockdown increases both amacrine cells and Müller glia in retina. Overexpression does not yield a detectable phenotype but forced mis-localisation phenocopies knockdown[72].<br><br>Knockdown in the medial prefrontal cortex increases anxiety-like behaviour without affecting long-term memory anxiety[73].<br><br>Knockout mice exhibit mild hyperactivity and increased sensitivity to methamphetamine, without major developmental or behavioural abnormalities[74].<br><br>Genetic association of rs1894720 and rs4274 SNPs with risk of paranoid schizophrenia[75].<br><br>Suppresses neuronal interferon response pathways affected in neuropsychiatric diseases[76].<br><br>Implicated in schizophrenia-associated alternative splicing[75-77]. | 22:26646411-26676478<br><br>*MIAT* | • Dementia<br>• Electroencephalogram measurement<br>• PHF-tau measurement<br>• Theta wave measurement | • Energy intake |

| | | | | |
|---|---|---|---|---|
| 55. *MIR600HG* (human) | Downregulated in autism.<br>Synapse-associated mRNAs are downregulated after knockdown [78]. | 9:123109494-123115477 | • Insomnia | • Body weight<br>• Sex hormone-binding globulin measurement |
| 56. *MSNP1AS* (human) | Antisense to a *moesin* pseudogene; moesin regulates neuronal architecture[79].<br>Highly overexpressed in postmortem cerebral cortex of individuals with autism spectrum disorder and decreases expression of moesin protein in human cell lines[79].<br>Influences the number and length of neurites in culture and overexpression inhibits neuron viability and migration and promotes apoptosis[80].<br>Genetic association of rs4307059 with autism[79]. | 5:25909503-25911234 | • Autism<br>• Hippocampal atrophy<br>• Neurofibrillary tangles measurement<br>• Pain<br>• Retinal nerve fibre layer thickness | • 3-hydroxydodecanedioate measurement<br>• Acute myeloid leukemia<br>• Body height |

| | | | | |
|---|---|---|---|---|
| 57. *N1LR / AK051854* (mouse, rat) | Enhances neuroprotection in ischemic stroke.<br><br>Overexpression is associated with reduced infarct volume and neurological deficit while knockdown results in increased infarct volume and greater neurological deficit[81]. | 3:136862233-136865932<br><br>None annotated | • Pain measurement | • Apolipoprotein a 1 measurement<br>• Apolipoprotein B measurement<br>• Alkaline phosphatase measurement<br>• Aspartate aminotransferase to alanine aminotransferase ratio<br>• Bilirubin measurement<br>• Blood protein amount<br>• Blood VLDL cholesterol amount<br>• Body height<br>• Fatty acid amount<br>• Free cholesterol measurement<br>• High density lipoprotein cholesterol measurement<br>• IgF-1 measurement<br>• Lipid measurement<br>• Low density lipoprotein cholesterol measurement<br>• Monocyte count<br>• Phospholipid amount<br>• Serum alanine aminotransferase amount<br>• Serum gamma-glutamyl transferase measurement<br>• Sex hormone-binding globulin measurement |

| | | | | |
|---|---|---|---|---|
| | | | | • Total cholesterol measurement<br>• Triglyceride measurement |

| | | | | |
|---|---|---|---|---|
| 58. *Neat1* (mouse, rat) | Suppresses hippocampus-dependent, long-term memory formation[82].<br><br>Regulates adaptive behavioural response to stress in mouse[83].<br><br>Knockout leads to hyperlocomotion, panic escape, social deficits, and disrupted circadian activity; neurons are hyperexcitable with altered calcium homeostasis, but not neurodegeneration nor inflammation[83].<br><br>Knockdown improves memory, alters gene expression and splicing, and reduces repressive H3K9me2 at key genes like *c-Fos*[82].<br><br>Overexpression impairs memory in young mice and mimics age-related cognitive decline via increased H3K9me2 and reduced *c-Fos* expression[82].<br><br>Increased in HD cell and animal models; transient knockdown reduces | 11:65422774-65445540<br><br>*NEAT1* | • Schizophrenia (family history) | • Abnormality of the skeletal system<br>• Acute myeloid leukemia<br>• BMI-adjusted hip circumference<br>• Diastolic blood pressure<br>• Hemoglobin A1 measurement<br>• Level of sorting nexin-15 in blood<br>• N6,N6,N6-trimethyllysine measurement<br>• N6,N6-dimethyllysine measurement<br>• Serum alanine aminotransferase amount<br>• Sex hormone-binding globulin measurement<br>• Systolic blood pressure |

| | | | | |
|---|---|---|---|---|
| | aggregates formed by mutant huntingtin[68]. | | | |
| 59. *NeuID* / *Gm10419* (mouse) | Downregulated in the brains of Alzheimer's disease (AD) patients. *NeuID* maintains neuronal identity by repressing developmental and glial genes via interaction with the PRC2 subunit EZH2 and regulation of H3K27me3. Knockdown of *NeuID* disrupts this repression, leading to impaired neuronal activity and memory formation. *NeuID* overexpression restores neuronal function in Aβ42-treated neurons[84]. | 4:576404-623311<br>*NEUID* / *LOC105374338* | • None | • 3-hydroxy-1-methylpropylmercapturic acid measurement<br>• Alpha-L-iduronidase measurement<br>• Level of spondin-2 in blood serum<br>• Type 2 diabetes mellitus |

| | | | | |
|---|---|---|---|---|
| 60. *NeuroLNC* (mouse, rat) | Regulates neurite elongation, neuronal migration and presynaptic activity by interacting with the neurodegeneration-associated protein TDP-43[85].<br><br>Expressed during retinal development[86].<br><br>Overexpression drastically increases the release of synaptic vesicles from neurons[85]. Overexpression significantly enhances Ca2+ influx while down-regulation decreases the influx of Ca2+[85].<br><br>Down-regulation decreases neurite length and branching while overexpression has the opposite effect[85].<br><br>Down-regulation decreases migration of neurons from the ventricular zone to the cortical plate, while overexpression has the opposite effect[85]. | 8:9899744-9919233<br><br>*MIR124-1HG* | • Alcohol consumption quality<br>• Alcohol drinking<br>• Amygdala volume<br>• Bipolar disorder<br>• Brain connectivity attribute<br>• Cerebral cortex area attribute<br>• Cognitive function measurement<br>• Cortical thickness<br>• Educational attainment<br>• Epilepsy<br>• Mathematical ability<br>• Neuroimaging measurement<br>• Neurotic disorder<br>• Neuroticism measurement<br>• Risk-taking behaviour<br>• Schizophrenia<br>• Self-reported educational attainment<br>• Sleep duration trait | • Adolescent idiopathic scoliosis<br>• Alkaline phosphatase measurement<br>• Allergen exposure measurement<br>• Antithrombotic agent use measurement<br>• Aortic measurement<br>• Appendicular lean mass<br>• Ascending aorta diameter<br>• Blood VLDL cholesterol amount<br>• BMI-adjusted hip circumference<br>• BMI-adjusted waist circumference<br>• BMI-adjusted waist-hip ratio<br>• Body fat percentage<br>• Body mass index<br>• Breastfeeding duration<br>• CCL11 measurement<br>• Cervical carcinoma<br>• Cholesterol:total lipids ratio<br>• Cholesteryl esters:total lipids ratio<br>• Complex trait<br>• Corneal endothelial cell attribute<br>• Coronary artery disease |

| | | | | |
|---|---|---|---|---|
| | | | • Smoking initiation<br>• Smoking status measurement<br>• Wellbeing measurement<br>• White matter hyperintensity measurement<br>• White matter microstructure measurement | • C-reactive protein measurement<br>• Cumulative dose response to bevacizumab<br>• Degree of unsaturation measurement<br>• Diastolic blood pressure<br>• Diet measurement<br>• Docosahexaenoic acid to total fatty acids percentage<br>• Facial morphology trait<br>• Fatty acid amount<br>• Glioma pathogenesis-related protein 1 measurement<br>• Glomerular filtration rate<br>• Gut microbiome measurement<br>• Hearing loss<br>• Heel bone mineral density<br>• High density lipoprotein cholesterol measurement<br>• Lean body mass<br>• Linoleic acid measurement<br>• Low density lipoprotein cholesterol measurement<br>• Mean arterial pressure<br>• Metabolic syndrome<br>• Multiple myeloma<br>• Obesity<br>• Omega-6 polyunsaturated fatty acid measurement |

| | | | | |
|---|---|---|---|---|
| | | | | • Pathological myopia<br>• Peak expiratory flow<br>• Peptic ulcer disease<br>• Polyunsaturated fatty acid measurement<br>• Polyunsaturated fatty acids to monounsaturated fatty acids ratio<br>• Polyunsaturated fatty acids to total fatty acids percentage<br>• Protein measurement<br>• Pulse pressure measurement<br>• Response to antineoplastic agent<br>• Response to radiation<br>• Saturated fatty acids measurement<br>• Sirtuin-2 measurement<br>• Systemic lupus erythematosus<br>• Systolic blood pressure<br>• Tinnitus<br>• Total cholesterol measurement<br>• Trauma exposure measurement<br>• Triglyceride measurement<br>• Triglycerides in IDL measurement<br>• Triglycerides:total lipids ratio |

| | | | | |
|---|---|---|---|---|
| 61. *Nkx2.2AS* (mouse) | Involved in oligodendrocyte differentiation. Antisense to homeobox protein NKX-2.2.<br><br>Forced expression of *Nkx2.2AS* in neural stem cells enhances oligodendrocyte differentiation.<br><br>Differentiation of MBP-positive and PLP-DM20-positive oligodendrocytes is retarded in *Nkx2.2*-null mutants along the rostrocaudal axis[87]. | 20:21511447-21513711<br><br>*NKX2-2-AS1* | • None | • None |
| 62. *Norad* / *ncRNA_BB* / *ncRNA_O* (mouse) | High expression pattern during the early stages of neuronal reprogramming, as well as in postnatal mouse and human brain[1]. Overlaps with a CNV morbidity map locus for neurodevelopmental disorders and craniofacial congenital malformations[1]. | 20:36042280-36051018<br><br>*NORAD* / *LOC467979* / *ncRNA_BB* / *ncRNA_O* | • None | • BMI-adjusted hip circumference<br>• Body mass index<br>• Cancer<br>• Docosahexaenoic acid measurement<br>• Melanoma<br>• Sexual dimorphism measurement<br>• Suntan<br>• Waist-hip ratio |

| | | | | |
|---|---|---|---|---|
| 63. *OLMALINC* (human) | Associated with regulation of oligodendrocyte maturation.<br><br>Knockdown affects genes regulating cell activation and membrane signalling in oligodendrocytes.<br><br>Knockdown in neurons affects cell proliferation pathway genes[88]. | 10:100372914-100454043 | • Cerebral cortex area attribute | • Alkaline phosphatase measurement<br>• Diastolic blood pressure<br>• Fatty acid amount<br>• Lysophosphatidylcholine 16:1 measurement<br>• Polyunsaturated fatty acids to monounsaturated fatty acids ratio<br>• Saturated fatty acids to total fatty acids percentage<br>• Triacylglycerol 50:3 measurement<br>• Triacylglycerol 52:3 measurement |

| | | | | |
|---|---|---|---|---|
| 64. *Particl / ncRNA_Z* (mouse) | High expression pattern during the early stages of neuronal reprogramming, as well as in postnatal mouse and human brain[1]. Overlaps with 2 CNV morbidity map loci for neurodevelopmental disorders and craniofacial congenital malformations[1]. | 2:85534705-85539110<br>*PARTICL / LOC100630918 / ncRNA_Z* | • None | • Albuminuria<br>• Basophil count<br>• Blood osmolality<br>• Body height<br>• Body weight<br>• Circulating fibrinogen levels<br>• Coronary artery disease<br>• COVID-19<br>• Eosinophil count<br>• Factor VII measurement<br>• Factor VIII measurement<br>• Factor XI measurement<br>• Granulysin measurement<br>• Heart disease<br>• Heart failure<br>• Level of trans-Golgi network integral membrane protein 2 in blood<br>• Level of tumor necrosis factor receptor superfamily member 10C in blood<br>• Macrophage-capping protein measurement<br>• Malunion fracture<br>• Mean corpuscular hemoglobin concentration<br>• Moderate albuminuria<br>• Myocardial infarction<br>• Neutrophil count |

| | | | | |
|---|---|---|---|---|
| | | | | • Neutrophil gelatinase-associated lipocalin measurement<br>• Platelet count<br>• Platelet-to-lymphocyte ratio<br>• Red cell distribution width<br>• Serum gamma-glutamyl transferase measurement<br>• Urinary albumin to creatinine ratio<br>• Vesicle-associated membrane protein 5 measurement<br>• Vesicle-associated membrane protein 8 measurement<br>• Von Willebrand factor quality |

| | | | | |
|---|---|---|---|---|
| 65. *Paupar* (mouse) | Involved in neural differentiation and regulates neural gene expression.<br><br>Knockdown disrupts the cell cycle profile of neuroblastoma cells and induces neural differentiation[89]. | 11:31812307-32002405<br><br>*PAUPAR* | • Alzheimer disease<br>• Anorexia nervosa<br>• Attention deficit hyperactivity disorder<br>• Autism spectrum disorder<br>• Bipolar disorder<br>• Cognitive function measurement<br>• Depressive symptom measurement<br>• Family history of Alzheimer's disease<br>• Insomnia<br>• Major depressive disorder<br>• Neuroticism measurement<br>• Obsessive-compulsive disorder<br>• PHF-tau measurement<br>• Schizophrenia<br>• Smoking initiation<br>• Tourette syndrome<br>• Wellbeing measurement | • 1-linoleoyl-2-linolenoyl-GPC (18:2/18:3) measurement<br>• 5-HIAA measurement<br>• Acute myeloid leukemia<br>• Age at onset<br>• Body height<br>• Body mass index<br>• Breast density<br>• Eye measurement<br>• HVA measurement<br>• Level of extracellular glycoprotein lacritin in blood<br>• Lipid measurement<br>• Omega-3 polyunsaturated fatty acid measurement<br>• Otosclerosis<br>• Pulse pressure measurement<br>• Systolic blood pressure<br>• Taste liking measurement<br>• Velopharyngeal dysfunction |

| | | | | |
|---|---|---|---|---|
| 66. *Prdm16-DT / Prdm16os* (mouse) | Regulates astrocyte function.<br><br>Knockdown results in loss of astrocyte homeostasis dysregulating genes essential for glutamate uptake, lactate release, and neuronal spine density through interactions with the RE1-Silencing Transcription factor and Polycomb Repressive Complex 2.<br><br>Overexpression mitigated stimuli-induced astrocyte functional deficits[90]. | 1:3059564-3070251<br><br>*PRDM16-DT* | • None | • BMI-adjusted waist-hip ratio<br>• BMI-adjusted waist circumference<br>• Body composition measurement<br>• Body height<br>• Coronary artery disease<br>• Growth/differentiation factor 8 measurement<br>• High density lipoprotein cholesterol measurement<br>• Monocyte count<br>• Myocardial infarction<br>• Platelet crit<br>• Pyridoxate measurement<br>• Waist-hip ratio |

| | | | | |
|---|---|---|---|---|
| 67. *Pnky / Gm30731* (mouse) | Regulates neuronal differentiation of embryonic and postnatal neural stem cells[91], neural stem cell migration[92] and production of projection neurons from neural stem cells[93].<br><br>Regulates neurogenesis and modulates cognitive and affective behaviors[94].<br><br>Knockdown increases neuronal differentiation in culture and *in vivo*[91].<br><br>Silencing suppressed but overexpression promoted *in vitro* migration of both murine neural stem cells[92].<br><br>Conditional deletion in developing cortex regulates production of projection neurons from neural stem cells, altering postnatal cortical lamination[93].<br><br>Knockout reduces fear context generalization in males and increases acoustic startle response in females[94]. | 6:98759637-98835826<br><br>*ENSG00000283010* and *ENSG00000309555* | • Attention deficit hyperactivity disorder<br>• Autism spectrum disorder<br>• Educational attainment<br>• Ganglion thickness<br>• Intelligence<br>• Memory performance | • Body height<br>• Drug allergy<br>• Glucagon measurement<br>• Sex interaction measurement<br>• Susceptibility to chickenpox measurement<br>• Trait in response to platinum |

| | |
|---|---|
| | Transgenic *Pnky* expression rescues acoustic startle response in females[94]. |

| | | | | |
|---|---|---|---|---|
| 68. *RMST* (human) | Required for neuronal differentiation from stem cells via binding SOX2 to promoter regions of neurogenic transcription factors[58,95,96].<br><br>Knockdown blocks neurogenesis[58,95].<br><br>Overexpression results in increased neuronal marker expression and a larger percentage of TUJ1-expressing neurons[95].<br><br>Implicated as a cause of KS, a rare Mendelian disorder manifested by abnormal puberty, hypogonadotropism, infertility and loss of smell. A rare translocation in a KS patient t(7;12)(q22;q24) affected *RMST*[97]. | 12:97386565-97598415 | • Alcohol consumption quality<br>• Cognitive function measurement<br>• Diffuse plaque measurement<br>• Educational attainment<br>• Empathy measurement<br>• Exploratory eye movement measurement<br>• Intelligence<br>• Migraine disorder<br>• Self-reported educational attainment<br>• Smoking initiation<br>• Social communication impairment<br>• Socioeconomic status<br>• Sudden cardiac arrest<br>• Vascular dementia | • Age at menarche<br>• Allergen exposure measurement<br>• Appendicular lean mass<br>• Base metabolic rate measurement<br>• Blood glucose amount<br>• Body height<br>• Body surface area<br>• Body weight<br>• Body mass index<br>• Cardioembolic stroke<br>• C-C motif chemokine 3 level<br>• Colorectal health<br>• COVID-19<br>• Gamma-aminoisobutyric acid measurement<br>• Gestational diabetes<br>• Glucose measurement<br>• Gut microbiome measurement<br>• HbA1c measurement<br>• Hemoglobin A1 measurement<br>• HLA allele carrier status<br>• Joint damage measurement<br>• Lean body mass<br>• Lipid measurement<br>• Mucocutaneous lymph node syndrome<br>• Obesity |

| | | | | |
|---|---|---|---|---|
| | | | | • Obstructive sleep apnea<br>• Prostate carcinoma<br>• Respiratory symptom measurement<br>• Revision of total knee arthroplasty<br>• RS-warfarin measurement<br>• Squamous cell carcinoma<br>• Type 2 diabetes mellitus<br>• Vaginal microbiome measurement<br>• Vitamin D amount<br>• Whole body water mass |
| 69. *RNCR4* (mouse) | Timed expression during retinal development, regulates processing of pri-miR-183/96/182 and retina architecture[86,98]. | 7:129779001-129781249<br>*ENSG00000304993*<br>*ENSG00000290319* | • None | • None |

| | | | | |
|---|---|---|---|---|
| 70. *RUS / LINC01322* (mouse) | Required for neuronal differentiation.<br>Knockdown locks neuronal precursors in an intermediate state towards neuronal differentiation resulting in arrested cell cycle and increased apoptosis[99]. | 3:165206440-165846519<br>*LINC01322* | • Circadian rhythm<br>• Diffuse plaque measurement<br>• Educational attainment<br>• Intelligence<br>• Lifestyle measurement<br>• Mathematical ability<br>• PHF-tau measurement<br>• Self-reported educational attainment<br>• Smoking behavior trait<br>• Socioeconomic status<br>• Tobacco smoke exposure measurement | • A disintegrin and metalloproteinase with thrombospondin motifs 3 measurement<br>• Acidic leucine-rich nuclear phosphoprotein 32 family member A measurement<br>• Adenylyltransferase and sulfurtransferase MOCS3 measurement<br>• Adolescent idiopathic scoliosis<br>• ADP-ribosylation factor-like protein 11 measurement<br>• Adrenomedullin measurement<br>• Adseverin measurement<br>• Aldehyde dehydrogenase, dimeric NADP-preferring measurement<br>• Alpha-N-acetylgalactosaminide alpha-2,6-sialyltransferase 2 measurement<br>• Alpha-N-acetylgalactosaminide alpha-2,6-sialyltransferase 3 measurement<br>• Angiopoietin-related protein 4 measurement<br>• Ankle injury |

| | | | | |
|---|---|---|---|---|
| | | | | • Anosmin-1 measurement<br>• Apolipoprotein B measurement<br>• Apolipoprotein E measurement<br>• Apolipoprotein F measurement<br>• Apolipoprotein M measurement<br>• Arginine/serine-rich protein 1 measurement<br>• Aspirin hydrolysis measurement<br>• Astrocytoma<br>• ATP synthase subunit beta, mitochondrial measurement<br>• ATPase ASNA1 measurement<br>• ATP-dependent RNA helicase A measurement<br>• Augurin measurement<br>• Aurora kinase A measurement<br>• Aurora kinase B measurement<br>• Axin-2 measurement<br>• Baculoviral IAP repeat-containing protein 5 measurement<br>• BAG family molecular chaperone regulator 4 measurement |

| | | | | |
|---|---|---|---|---|
| | | | | • BCL-2-like protein 1 measurement<br>• BCL-2-related protein A1 measurement<br>• Beta-defensin 107 measurement<br>• Beta-defensin 108B measurement<br>• Beta-defensin 110 measurement<br>• Biglycan measurement<br>• Blood protein amount<br>• Bone density<br>• BPI fold-containing family A member 1 measurement<br>• Breastfeeding duration<br>• Butyrylcholinesterase measurement<br>• Cadherin-related family member 5 measurement<br>• Carbohydrate sulfotransferase 14 measurement<br>• Carbonic anhydrase 2 measurement<br>• Carboxypeptidase e measurement<br>• Cardioembolic stroke<br>• Casein kinase II subunit alpha measurement<br>• Caveolin-2 measurement |

| | | | | |
|---|---|---|---|---|
| | | | | • C-C motif chemokine 1 measurement<br>• C-C motif chemokine 3-like 1 measurement<br>• CCAAT/enhancer-binding protein beta measurement<br>• CCL20 measurement<br>• Cellular retinoic acid-binding protein 1 measurement<br>• Cellular tumor antigen p53 measurement<br>• Chitinase-3-like protein 2 measurement<br>• Chromobox protein homolog 5 measurement<br>• Chronic obstructive pulmonary disease<br>• Chymotrypsin-like protease CTRL-1 measurement<br>• Citrate measurement<br>• Coiled-coil domain-containing protein 126 measurement<br>• Cold shock domain-containing protein C2 measurement<br>• Collectin-10 measurement<br>• Complement component 1 q subcomponent-binding protein, mitochondrial measurement |

| | | | | |
|---|---|---|---|---|
| | | | | • Complement component C8 measurement<br>• Complement factor D measurement<br>• Coronary artery calcification<br>• CREB-binding protein measurement<br>• C-type lectin domain family 4 member D measurement<br>• C-type lectin domain family 4 member M amount<br>• Cyclin-dependent kinase 2-associated protein 1 measurement<br>• Cyclin-dependent kinase 5:cyclin-dependent kinase 5 activator 1 complex measurement<br>• Cyclin-dependent kinase 8:cyclin-c complex measurement<br>• Cysteine-rich motor neuron 1 protein measurement<br>• Cytochrome b-c1 complex subunit 7 measurement<br>• Cytochrome c oxidase assembly factor 3 homolog, mitochondrial measurement<br>• Cytochrome c oxidase subunit 4 isoform 2, mitochondrial measurement |

| | | | | |
|---|---|---|---|---|
| | | | | • Cytochrome p450 3a4 measurement<br>• Cytohesin-2 measurement<br>• Cytokine measurement<br>• Cytoskeleton-associated protein 2 measurement<br>• Cytosolic non-specific dipeptidase measurement<br>• DCC-interacting protein 13-alpha measurement<br>• Deoxynucleoside triphosphate triphosphohydrolase SAMHD1 measurement<br>• Dermokine measurement<br>• Desmoglein-1 measurement<br>• Dickkopf-like protein 1 measurement<br>• Dipeptidyl peptidase 1 measurement<br>• DNA repair protein RAD51 homolog 1 amount<br>• DNA-binding protein SATB2 measurement<br>• DNA-directed RNA polymerases I and III subunit RPAC1 measurement<br>• Dolichyl-diphosphooligosaccharide--protein glycosyltransferase subunit 1 measurement |

| | | | | |
|---|---|---|---|---|
| | | | | • Dual specificity mitogen-activated protein kinase kinase 1 measurement<br>• Dual specificity protein kinase CLK2 measurement<br>• Dual specificity protein phosphatase 16 measurement<br>• Dual specificity tyrosine-phosphorylation-regulated kinase 3 measurement<br>• Dynactin subunit 2 measurement<br>• E3 ubiquitin-protein ligase NEURL1 measurement<br>• E3 ubiquitin-protein ligase RNF128 measurement<br>• E3 ubiquitin-protein ligase RNF149 measurement<br>• E3 ubiquitin-protein ligase RNF43 measurement<br>• Early endosome antigen 1 measurement<br>• Ectodysplasin-a, secreted form measurement<br>• Ectonucleoside triphosphate diphosphohydrolase 1 measurement<br>• Ectonucleoside triphosphate diphosphohydrolase 3 measurement |

| | | | | |
|---|---|---|---|---|
| | | | | • Endothelial differentiation-related factor 1 measurement<br>• Endothelin-converting enzyme 1 measurement<br>• Enhancer of mRNA-decapping protein 4 measurement<br>• Epididymal-specific lipocalin-10 measurement<br>• ES1 protein homolog, mitochondrial measurement<br>• Estrogen sulfotransferase measurement<br>• Eukaryotic translation initiation factor 5 amount<br>• Fatty acid-binding protein, epidermal measurement<br>• Fatty acid-binding protein, liver measurement<br>• F-box/LRR-repeat protein 4 measurement<br>• FEV/FVC ratio<br>• Focal adhesion kinase 1 measurement<br>• Forced expiratory volume<br>• Forkhead box protein C2 measurement<br>• Fractalkine measurement<br>• Furin measurement<br>• General transcription factor II-I measurement |

| | | | | |
|---|---|---|---|---|
| | | | | • Glutathione S-transferase A4 measurement<br>• Glycerol-3-phosphate dehydrogenase 1-like protein measurement<br>• Glycoprotein hormone alpha-2 measurement<br>• Glypican-2 measurement<br>• Glypican-6 measurement<br>• Golgi SNAP receptor complex member 1 measurement<br>• G-protein coupled receptor 26 measurement<br>• Granulocyte colony-stimulating factor level<br>• Growth factor receptor-bound protein 14 measurement<br>• Growth factor receptor-bound protein 7 measurement<br>• Guanylate-binding protein 6 measurement<br>• Gut microbiome measurement<br>• Heart failure<br>• Heat shock 70 kDa protein 1-like measurement<br>• Heat shock 70 kDa protein 6 measurement<br>• Hephaestin-like protein 1 measurement |

| | | | | |
|---|---|---|---|---|
| | | | | • Heterogeneous nuclear ribonucleoproteins C1/C2 measurement<br>• High mobility group protein B2 measurement<br>• High mobility group protein B3 measurement<br>• Histone acetyltransferase type b catalytic subunit measurement<br>• Histone-lysine n-methyltransferase EHMT2 measurement<br>• Histone-lysine N-methyltransferase SETD2 measurement<br>• Inactive dipeptidyl peptidase 10 measurement<br>• Inactive gamma-glutamyltranspeptidase 2 measurement<br>• Inactive peptidyl-prolyl cis-trans isomerase FKBP6 measurement<br>• Insulin growth factor-like family member 3 measurement<br>• Insulin-like 3 measurement<br>• Integral membrane protein 2B measurement<br>• Integrin alpha-5 measurement |

| | | | | |
|---|---|---|---|---|
| | | | | • Interferon alpha-5 measurement<br>• Interferon gamma measurement<br>• Interleukin-10 receptor subunit alpha measurement<br>• Interleukin-17A measurement<br>• Interleukin-34 measurement<br>• Interleukin-37 measurement<br>• Kallikrein-12 measurement<br>• Kallikrein-6 measurement<br>• Kelch-like protein 13 measurement<br>• Keratin, type II cytoskeletal 7 measurement<br>• Keratinocyte differentiation-associated protein measurement<br>• Killer cell immunoglobulin-like receptor 2DL2 measurement<br>• Kin of IRRE-like protein 3 measurement<br>• Kinesin-like protein KIF16B measurement<br>• Kinetochore protein NDC80 homolog measurement<br>• Kv channel-interacting protein 1 measurement<br>• Ladinin-1 measurement |

| | | | | |
|---|---|---|---|---|
| | | | | • Laminin subunit alpha-4 measurement<br>• Layilin measurement<br>• Leucine-rich repeat and fibronectin type III domain-containing protein 1 measurement<br>• Leucine-rich repeat and transmembrane domain-containing protein 2 measurement<br>• Leucine-rich repeat neuronal protein 1 measurement<br>• Leucine-rich repeat serine/threonine-protein kinase 2 measurement<br>• Leucine-rich repeat-containing protein 74A measurement<br>• Level of cholinesterase in blood<br>• Level of triacylglycerol (56:6) in blood serum<br>• Low-density lipoprotein receptor measurement<br>• Lymphotoxin alpha2:beta1 measurement<br>• Lysophosphatidylcholine 20:3 measurement<br>• Lysosome membrane protein 2 amount |

| | | | | |
|---|---|---|---|---|
| | | | | • Macrophage-stimulating protein receptor measurement<br>• Magnesium transporter NIPA4 measurement<br>• Malignant T-cell-amplified sequence 1 measurement<br>• MAP kinase-activated protein kinase 5 measurement<br>• Mast cell-expressed membrane protein 1 measurement<br>• MICAL-like protein 2 measurement<br>• Mitochondrial DNA measurement<br>• Mitochondrial import inner membrane translocase subunit TIM14 amount<br>• M-phase inducer phosphatase 2 measurement<br>• Mucin-1 measurement<br>• Multimerin-2 measurement<br>• Myelin regulatory factor measurement<br>• Myocardial zonula adherens protein measurement<br>• Myotubularin-related protein 1 measurement<br>• Nectin-2 measurement |

| | | | | |
|---|---|---|---|---|
| | | | | • NKG2-D type II integral membrane protein amount<br>• Nuclear pore complex-interacting protein family member B3 measurement<br>• O-acetyl-ADP-ribose deacetylase MACROD1 measurement<br>• OCIA domain-containing protein 1 measurement<br>• Oncostatin-M measurement<br>• Orexigenic neuropeptide QRFP measurement<br>• OX-2 membrane glycoprotein amount<br>• Oxysterols receptor LXR-beta measurement<br>• Palmitoyl-protein thioesterase 1 measurement<br>• PAX-interacting protein 1 measurement<br>• Peak expiratory flow<br>• Pentraxin-related protein PTX3 measurement<br>• Peptidyl-prolyl cis-trans isomerase-like 2 measurement<br>• PH and SEC7 domain-containing protein 1 measurement |

| | | | | |
|---|---|---|---|---|
| | | | | • PH and SEC7 domain-containing protein 2 measurement<br>• Phosphatidylinositol 4,5-bisphosphate 3-kinase catalytic subunit gamma isoform measurement<br>• Phosphatidylinositol 4-phosphate 3-kinase C2 domain-containing subunit alpha measurement<br>• Phosphatidylinositol transfer protein alpha isoform measurement<br>• Phosphatidylinositol transfer protein beta isoform measurement<br>• Physical activity measurement<br>• Platelet endothelial cell adhesion molecule measurement<br>• Poly(A) RNA polymerase, mitochondrial measurement<br>• Poly(U)-specific endoribonuclease measurement<br>• Potassium voltage-gated channel subfamily A member 10 measurement<br>• Potassium voltage-gated channel subfamily E |

| | | | | |
|---|---|---|---|---|
| | | | | regulatory beta subunit 5 measurement<br>• PR domain zinc finger protein 1 measurement<br>• Pregnancy-specific beta-1-glycoprotein 5 measurement<br>• Pre-mRNA-processing factor 6 measurement<br>• Pre-mRNA-splicing factor ATP-dependent RNA helicase PRP16 measurement<br>• Probable E3 ubiquitin-protein ligase HERC4 measurement<br>• Probable E3 ubiquitin-protein ligase MID2 measurement<br>• Prokineticin-1 measurement<br>• Prostate and testis expressed protein 4 measurement<br>• Proteasome subunit alpha type-1 amount<br>• Proteasome subunit beta type-5 measurement<br>• Protein CEI measurement<br>• Protein measurement<br>• Protein NDRG4 measurement<br>• Protein RIC-3 measurement<br>• Protein S100-A5 measurement |

| | | | | |
|---|---|---|---|---|
| | | | | • Protein sel-1 homolog 2 measurement<br>• Protein SERAC1 measurement<br>• Protein set measurement<br>• Protein TMEPAI measurement<br>• Protein transport protein Sec61 subunit gamma measurement<br>• Protocadherin beta-1 measurement<br>• Protocadherin beta-2 measurement<br>• Pyridoxal kinase measurement<br>• RAC-beta serine/threonine-protein kinase measurement<br>• Radiation-inducible immediate-early gene IEX-1 measurement<br>• Ras-related protein Rab-27B measurement<br>• Ras-related protein Rab-35 measurement<br>• Receptor tyrosine-protein kinase erbb-4 measurement<br>• Receptor-interacting serine/threonine-protein kinase 2 measurement<br>• Recoverin measurement |

| | | | | |
|---|---|---|---|---|
| | | | | • Redox-regulatory protein FAM213A measurement<br>• Regulator of G-protein signaling 19 measurement<br>• Regulator of G-protein signaling 3 measurement<br>• Response to xenobiotic stimulus<br>• Retinoblastoma-associated protein measurement<br>• Rho GTPase-activating protein 1 measurement<br>• Rho GTPase-activating protein 30 measurement<br>• Rho GTPase-activating protein 36 measurement<br>• Rho guanine nucleotide exchange factor 2 measurement<br>• Rho guanine nucleotide exchange factor 7 measurement<br>• Rhophilin-2 measurement<br>• Rho-related GTP-binding protein Rho6 measurement<br>• Ribosome biogenesis protein TSR3 homolog measurement<br>• RING finger protein 122 measurement<br>• RNA polymerase II elongation factor ELL measurement |

| | | | | |
|---|---|---|---|---|
| | | | | • RNA-binding protein 24 measurement<br>• Roundabout homolog 3 measurement<br>• RS-warfarin measurement<br>• Secreted frizzled-related protein 2 measurement<br>• Serine palmitoyltransferase 1 measurement<br>• Serine/threonine-protein kinase 16 measurement<br>• Serine/threonine-protein kinase Chk1 measurement<br>• Serine/threonine-protein kinase PAK 3 measurement<br>• Serine/threonine-protein kinase ULK3 measurement<br>• Serine/threonine-protein phosphatase 2A 65 kDa regulatory subunit A alpha isoform measurement<br>• Serpin I2 measurement<br>• Serum albumin amount<br>• Serum amyloid A-1 protein amount<br>• Serum copper measurement<br>• Signal-regulatory protein gamma measurement<br>• Small integral membrane protein 9 measurement<br>• Small nuclear ribonucleoprotein F amount |

| | | | | |
|---|---|---|---|---|
| | | | | • Sodium-coupled monocarboxylate transporter 1 measurement<br>• Sodium-independent sulfate anion transporter measurement<br>• Sorting nexin-7 measurement<br>• Stabilin-2 measurement<br>• Stromal interaction molecule 1 measurement<br>• SUN domain-containing protein 5 measurement<br>• Tachykinin-4 measurement<br>• T-cell surface glycoprotein CD3 epsilon chain measurement<br>• T-cell surface glycoprotein CD8 beta chain measurement<br>• Teneurin-3 measurement<br>• Teneurin-4 measurement<br>• Tetratricopeptide repeat protein 17 measurement<br>• Thioredoxin domain-containing protein 11 measurement<br>• Thymidine kinase, cytosolic measurement<br>• Tight junction protein ZO-1 measurement |

| | | | | |
|---|---|---|---|---|
| | | | | • TNF-related activation-induced cytokine measurement<br>• Toll-like receptor 1 measurement<br>• Transcription factor AP-1 measurement<br>• Transcription factor IIIb 90 kda subunit measurement<br>• Transcription factor RelB measurement<br>• Transcription regulator protein BACH1 measurement<br>• Transcriptional regulator Kaiso measurement<br>• Transmembrane protein 59-like measurement<br>• tRNA (guanine-N(7)-)-methyltransferase measurement<br>• Tropomyosin beta chain measurement<br>• Tuftelin-interacting protein 11 measurement<br>• Tumor necrosis factor ligand superfamily member 14 measurement<br>• Tumor necrosis factor ligand superfamily member 15 amount |

| | | | | |
|---|---|---|---|---|
| | | | | • Tumor necrosis factor ligand superfamily member 18 amount<br>• Tumor necrosis factor receptor superfamily member 19L amount<br>• Tumor necrosis factor receptor superfamily member 3 amount<br>• Tumor necrosis factor receptor superfamily member 4 amount<br>• Tumor protein p53-inducible protein 11 measurement<br>• Tyrosine-protein kinase ZAP-70 measurement<br>• U6 snRNA phosphodiesterase measurement<br>• UDP-GlcNAc:betaGal beta-1,3-N-acetylglucosaminyltransferase 6 measurement<br>• UDP-glucuronosyltransferase 1-6 measurement<br>• Uncharacterized protein C19orf18 measurement<br>• Uncharacterized protein C3orf18 measurement<br>• Urokinase-type plasminogen activator measurement |

| | | | | |
|---|---|---|---|---|
| | | | | • Uroplakin-3b-like protein measurement<br>• Valine--tRNA ligase measurement<br>• Versican core protein measurement<br>• Vesicle-associated membrane protein 4 measurement<br>• V-set and transmembrane domain-containing protein 4 measurement<br>• YTH domain-containing protein 1 measurement<br>• Zinc finger protein 18 measurement<br>• Zinc finger protein 180 measurement<br>• Zinc transporter 3 measurement<br>• Zona pellucida sperm-binding protein 4 measurement |
| 71. *SCAANT1* (human) | Antisense to *ataxin-7*, which causes the neurodegenerative disorder SCA7.<br>Loss of SCAANT1 derepresses *ataxin*-7 transcription[100]. | 3:63911512-63912383 | • None | • Type 2 diabetes mellitus |

| | | | | |
|---|---|---|---|---|
| 72. *SCN1ANAT* (monkey) | Antisense to the the voltage-gated sodium channel gene *SCN1A.*<br><br>Knockdown leads to upregulation of *SCN1A*, improves seizure phenotype, and increases excitability of hippocampal interneurons[101]. | 2:165957188-166390771<br><br>*SCN1A-AS1* | • Aggressive behavior quality<br>• Cannabis dependence measurement<br>• Educational attainment<br>• Epilepsy<br>• Febrile seizure (within the age range of 3 months to 6 years)<br>• Generalised epilepsy<br>• Grey matter density measurement<br>• Mathematical ability<br>• Mesial temporal lobe epilepsy with hippocampal sclerosis<br>• MMR-related febrile seizures<br>• Partial epilepsy<br>• PHF-tau measurement<br>• Schizophrenia<br>• Self-reported educational attainment<br>• Sleep duration trait | • Acute myeloid leukemia<br>• Biliary atresia<br>• Body height<br>• Body mass index<br>• Bone density<br>• Dental caries<br>• Diverticular disease<br>• IgA glomerulonephritis<br>• Lipid measurement<br>• Oxalate measurement<br>• Polypeptide N-acetylgalactosaminyltransferase 3 measurement<br>• Response to xenobiotic stimulus<br>• Soluble triggering receptor expressed on myeloid cells 2 measurement<br>• Threonate measurement<br>• Triacylglycerol 58:10 measurement |

| | | | | |
|---|---|---|---|---|
| 73. *Silc1* (mouse) | Important for neuronal regeneration in the peripheral nervous system[56].<br>Required for efficient spatial learning and memory formation[56,102].<br>Knockdown leads to reduction in total axonal outgrowth and reduces SOX11[56].<br>Knockout reduces SOX11 in neurons and mice exhibit delayed regeneration following injury[56].<br>Knockdown reduces activity-induced *Sox11* expression in mature hippocampal neurons and delays spatial learning without affecting long-term memory. Overexpression does not alter *Sox11* expression[102].<br>*Silc1* levels decline in a mouse model of AD[102]. | 2:5932543-6060570<br>*SILC1* | • Brain attribute<br>• Chronic pain<br>• Cortical thickness<br>• Neuroimaging measurement<br>• Nicotine dependence symptom count<br>• PHF-tau measurement<br>• Smoking behavior<br>• Stuttering | • Acute myeloid leukemia<br>• Age at first sexual intercourse measurement<br>• AL amyloidosis<br>• Amino acid measurement<br>• Body fat percentage<br>• Body height<br>• Body mass index<br>• Body weight<br>• Bone density<br>• Breastfeeding duration<br>• Calvaria morphology trait<br>• Cervical Intraepithelial Neoplasia Grade 2/3<br>• Comparative body size at age 10, self-reported<br>• Conotruncal heart malformations<br>• Coronary artery calcification<br>• Coronary artery disease<br>• COVID-19<br>• Cytotoxicity measurement<br>• Diet measurement<br>• Drug-induced agranulocytosis<br>• Environmental exposure measurement<br>• Eotaxin measurement<br>• Fat pad mass<br>• Glioma pathogenesis-related protein 1 measurement |

| | | | | |
|---|---|---|---|---|
| | | | | • Glutamine measurement<br>• Gut microbiome measurement<br>• Heart amyloid deposition<br>• Heart failure<br>• Hip circumference<br>• HIV-1 infection<br>• IgF-1 measurement<br>• Kidney amyloid amount<br>• Level of transcription initiation factor TFIID subunit 12 in blood serum<br>• Lower body strength measurement<br>• Metabolic syndrome<br>• Osteosarcoma<br>• Response to clozapine<br>• Response to metformin<br>• Severe acute respiratory syndrome<br>• Type 2 diabetes mellitus<br>• Urate measurement<br>• Vaginal microbiome measurement |

| | | | | |
|---|---|---|---|---|
| 74. *Six3OS1* / *RNCR1* (mouse) | Expressed during retinal development[86].<br><br>Regulates retinal cell fate specification[103].<br><br>Implicated in neuronal modulation by inhibiting glial cell differentiation[104].<br><br>Overexpression results in no significant change in any major retinal cell type[103].<br><br>Knockdown results in decreased protein kinase C α (PKCα) positive rod bipolar cells and increased glutamine synthetase-positive Muller glia[103].<br><br>Knockdown decreases the number of neurons and oligodendrocytes and increases the number of astrocytes in culture[104]. | 2:44921054-44939344<br><br>*LINC01833* | • Age at initiation of smoking<br>• Alcohol and nicotine codependence<br>• Alcohol consumption quality<br>• Alcohol dependence<br>• Alcohol use disorder measurement<br>• Amount of iron in brain<br>• Attention deficit hyperactivity disorder<br>• Bitter alcoholic beverage consumption measurement<br>• Brain attribute<br>• Brain volume<br>• Cannabis dependence<br>• Cannabis use<br>• Cerebral cortex area attribute<br>• Cognitive function measurement<br>• Cortical thickness<br>• Educational attainment | • 3-hydroxypropylmercapturic acid measurement<br>• Abnormality of refraction<br>• Body height<br>• Body mass index<br>• Calcium measurement<br>• Cholelithiasis<br>• Chronic obstructive pulmonary disease<br>• Comparative body size at age 10, self-reported<br>• Diet measurement<br>• Facial height measurement<br>• Intraocular pressure measurement<br>• Life span trait<br>• Mosquito bite reaction size measurement<br>• Myopia<br>• Obstructive sleep apnea<br>• Open-angle glaucoma<br>• Optic disc size trait<br>• Ovarian serous carcinoma<br>• Physical activity measurement<br>• Progression free survival<br>• Protein measurement<br>• Refractive error<br>• Response to carboplatin<br>• Serum gamma-glutamyl transferase measurement |

| | | | | |
|---|---|---|---|---|
| | | | • Executive function measurement<br>• Longitudinal alcohol consumption measurement<br>• Neuroimaging measurement<br>• Nicotine dependence<br>• Opioid use disorder<br>• Risk-taking behaviour<br>• Schizophrenia<br>• Self-reported educational attainment<br>• Smoking behavior<br>• Smoking behavior trait<br>• Smoking cessation<br>• Smoking initiation<br>• Smoking status measurement<br>• Social inhibition quality<br>• Substance abuse<br>• Substance-related disorder<br>• Tobacco smoke exposure measurement | • Taste liking measurement<br>• Trait in response to dolutegravir<br>• Trait in response to paclitaxel |

| | | | | |
|---|---|---|---|---|
| 75. *SLAMR* / *2610035D17Rik* (mouse) | *SLAMR* is recruited to the synapse upon stimulation; regulates activity-dependent synaptic plasticity and consolidation of long-term fear memory.<br><br>Knockdown induces a significant decrease in dendritic arborization.<br><br>Knockdown in dorsal hippocampus CA1 impairs consolidation of contextual fear memory, but not acquisition, extinction, recall, or spatial memory consolidation[105]. | 17:72221072-72640472<br><br>*LINC00511* | • Hippocampal volume<br>• Parkinson disease | • Abnormality of chromosome segregation<br>• Appetite-regulating hormone measurement<br>• Base metabolic rate measurement<br>• Bladder calculus<br>• Blood glucose amount<br>• BMI-adjusted waist circumference<br>• BMI-adjusted waist-hip ratio<br>• Body height<br>• Breast carcinoma<br>• Breastfeeding duration<br>• Cleft lip<br>• Colorectal adenoma<br>• Colorectal cancer<br>• COVID-19<br>• Crohn's disease<br>• Cytomegalovirus infection<br>• Cytotoxicity measurement<br>• Dental caries<br>• Dentures<br>• Diet measurement<br>• FEV/FVC ratio<br>• Glomerular filtration rate<br>• Gut microbiome measurement<br>• Handedness<br>• High density lipoprotein cholesterol measurement |

| | | | | |
|---|---|---|---|---|
| | | | | • Hip geometry<br>• Hyperthyroidism<br>• Inflammatory bowel disease<br>• Level of gastrin in blood<br>• Level of thyrotropin subunit beta in blood<br>• Lung adenocarcinoma<br>• Lung carcinoma<br>• Neck of femur size<br>• Nephrolithiasis<br>• Osteoarthritis<br>• Pancreatic carcinoma<br>• Prostate specific antigen amount<br>• Response to allogeneic hematopoietic stem cell transplant<br>• S-7-hydroxywarfarin measurement<br>• Serum creatinine amount<br>• Severe acute respiratory syndrome<br>• Thyroid stimulating hormone amount<br>• Total hip arthroplasty<br>• Trait in response to hydrochlorothiazide<br>• Trait in response to Triptolide<br>• Triglyceride measurement<br>• Type 2 diabetes mellitus |

| | | | | |
|---|---|---|---|---|
| | | | | • Urolithiasis<br>• Ulcerative colitis<br>• Velopharyngeal dysfunction<br>• Waist-hip ratio |
| 76. *SMN-AS1* (human) | Antisense to the Survival Motor Neuron protein (SMN), the lack of which results in Spinal Muscular Atrophy.<br>Represses the expression of *SMN*[106]. | 5:70931243-70932840 | • None | • None |
| 77. *Snhg11* (mouse) | Required for proper neurogenesis.<br>Essential for hippocampal-dependent memory formation.<br>Knockdown in dentate gyrus impairs adult neurogenesis.<br>Impaired synaptic plasticity, short-term and long-term memory, and pattern separation memory. No effect on fear conditioning[107]. | 20:38446343-38450943<br>*SNHG11* | • None | • Lipopolysaccharide-binding protein measurement |

| | | | | |
|---|---|---|---|---|
| 78. *Snhg15 / ncRNA_L* (mouse) | High expression pattern during the early stages of neuronal reprogramming, as well as in postnatal and adult mouse brain[1]. | 7:44983019-44986961<br>*SNHG15* | • Educational attainment | • Body height<br>• Cerebral cavernous malformations 2 protein measurement<br>• Diastolic blood pressure<br>• Leukocyte quantity<br>• Lymphocyte count<br>• Platelet crit |

| | | | | |
|---|---|---|---|---|
| 79. *Sox2ot* (mouse) | Expressed in several regions of the mouse and human brain. Also dynamically regulated during chicken and zebrafish embryogenesis, associated with central nervous system structures [108].<br><br>Regulates the differentiation of cortical neural progenitors[109,110]. | 3:180989510-181836880<br><br>*SOX2-OT* | • Anorexia nervosa<br>• Attention deficit hyperactivity disorder<br>• Bipolar disorder<br>• Eating disorder<br>• Educational attainment<br>• Income<br>• Insomnia<br>• Insomnia measurement<br>• Intelligence<br>• Lifestyle measurement<br>• Major depressive disorder<br>• Memory performance<br>• Pain measurement<br>• Risk-taking behaviour<br>• Schizophrenia<br>• Self-reported educational attainment<br>• Sleep duration trait<br>• Smoking initiation<br>• Smoking status measurement<br>• Social inhibition quality | • Age at onset<br>• Age-related nuclear cataract<br>• Aging rate<br>• Ankle injury<br>• Anti-Toxoplasma gondii IgG measurement<br>• Astrocytoma<br>• Balding measurement<br>• Blood insulin amount<br>• Body height<br>• Body mass index<br>• Bone density<br>• Cataract<br>• Cleft lip<br>• Color vision disorder<br>• Corneal astigmatism<br>• Corneal topography<br>• COVID-19<br>• Cystatin-M measurement<br>• Dental caries<br>• Diastolic blood pressure<br>• Eye measurement<br>• Facial hair thickness<br>• Glioma pathogenesis-related protein 1 measurement<br>• Hallux valgus<br>• Hirsutism<br>• Interferon gamma measurement<br>• Lean body mass |

| | | | | |
|---|---|---|---|---|
| | | | • Socioeconomic status<br>• Substance abuse | • Level of desmoglein-3 in blood serum<br>• Level of desmoglein-4 in blood serum<br>• Level of fatty acid-binding protein 9 in blood<br>• Level of phospholipase A2 inhibitor and Ly6/PLAUR domain-containing protein in blood<br>• Level of phospholipase B1, membrane-associated in blood<br>• Metabolic syndrome<br>• Metabolite measurement<br>• Oligodendroglioma<br>• Protein measurement<br>• Puberty onset measurement<br>• Reaction time measurement<br>• S-6-hydroxywarfarin measurement<br>• Strand of hair color<br>• Synophrys measurement<br>• Type 2 diabetes mellitus |

| | | | | |
|---|---|---|---|---|
| 80. *Synage / Gm19872 / Gm2694 / linc1582* (mouse) | Controls pluripotent stem cell state and regulates neuroectoderm differentiation[111].<br><br>Regulates synaptic stability and function during cerebellar development[112].<br><br>Regulates endoplasmic reticulum homeostasis, surface expression of AMPA receptors, and regulates excitatory synaptic transmission in depression[113].<br><br>Knockdown reduced levels of pluripotency markers such as *Nanog* and *Oct4*[111].<br><br>Knockout results in cerebellar atrophy and neuronal loss during cerebellar development[112].<br><br>Knockdown enhances excitatory synaptic transmission and alleviates depressive-like behaviours induced by chronic social defeat stress, while overexpression increases vulnerability to stress and social avoidance[113]. | 16:49282037-49350504<br><br>*ENSG00000279249* | • Cerebellin-1 measurement | • Blood protein amount<br>• Bmi-adjusted waist circumference<br>• Body height<br>• Body mass index<br>• Body weight<br>• Colorectal cancer<br>• Delivery measurement<br>• Gut microbiome measurement |

| | | | | |
|---|---|---|---|---|
| 81. *Tsx* (mouse) | Modulates hippocampal short-term memory and fear responses.<br>Male mice lacking *Tsx* exhibit reduced fear responses and enhanced hippocampal-dependent short-term memory[114]. | X:73729372-73782373<br>None | • None | • None |
| 82. *Tug1* (mouse) | Required for differentiation of the retina and is involved in cell survival.<br>Knockdown results in malformed or non-existent outer segments of photoreceptors.<br>Overexpression results in no obvious phenotype[115]. | 22:30969245-30979395<br>*TUG1* | • Alzheimer disease<br>• Bulimia nervosa<br>• Cognitive function measurement<br>• Educational attainment<br>• Intelligence<br>• Mathematical ability<br>• Self-reported educational attainment | • Body height<br>• Erythrocyte volume<br>• Protein measurement<br>• Transcobalamin-2 measurement |

| | | | | |
|---|---|---|---|---|
| 83. *Ube3a1* (rat) | A splice variant of the *Ube3a* gene, whose function is coding independent but requires 3'UTR sequences.<br>Negative regulator of dendritogenesis in mature hippocampal neurons.<br>Knockdown increased dendritic complexity *in vitro* and *in vivo* and activity of the plasticity-regulating *miR-134*[116]. | 15:25333728-25439051<br>*UBE3A1* | • None | • Bone density<br>• Prostate carcinoma |
| 84. *UBE3A-ATS* (mouse) | Antisense to *UBE3A*, which encodes an E3 ubiquitin ligase[117]. | 15:24978583-25420336<br>*SNHG14* | • Alzheimer disease<br>• Family history of Alzheimer's disease<br>• Generalized anxiety disorder<br>• Neuroticism measurement<br>• Panic disorder<br>• PHF-tau measurement | • Asthma<br>• Atrophic macular degeneration<br>• Blood osmolality<br>• Bone density<br>• Glioblastoma multiforme<br>• Inflammatory biomarker measurement<br>• Myopia<br>• Prostate carcinoma<br>• RS-10-hydroxywarfarin measurement<br>• S-methylcysteine sulfoxide measurement<br>• X-24812 measurement<br>• YKL40 measurement |

| | | | | |
|---|---|---|---|---|
| 85. *Uchl1-AS* (mouse) | Stress-responsive regulator of dopaminergic neuronal survival[118].<br>Increases UCHL1 protein synthesis at the post-transcriptional level[119].<br>Down-regulated with stimulated dopamine cell death *in vitro* and *in vivo*[118]. | 4:41220074-41256727<br>*UCHL1-DT* | • None | • Bone density<br>• Blood glucose amount<br>• High density lipoprotein cholesterol measurement |
| 86. *utNgn1* (mouse) | Enhancer-derived lncRNA that regulates the expression of neurogenin and neuronal differentiation.<br>Knockdown reduces expression of *Neurog1* and partially inhibited the induction of Tbr2 and NeuroD1, early markers of neocortical neuronal fate commitment[120]. | 5:135536347-135544103<br>*ENSG00000294972*[c] | • None | • Level of thioredoxin domain-containing protein 15 in blood |

[c] Possible ortholog based on positions of CpG islands at locus.

| | | | | |
|---|---|---|---|---|
| 87. *Zeb2os* (mouse) | Involved in astrocyte reactive astrogliosis.<br>Knockdown decreases expression of *Zeb2os*, *Zeb2*, and *Gfap*, and decreased astrocyte proliferation.<br>Knockdown also reduced percentages of GFAP-immunoreactive areas in a spinal cord injury model[121]. | 2:144517978-144521491<br>*ZEB2-AS1* | • None | • None |
| 88. *Zfh5-as* / *Zfhx2os* (mouse) | Regulates the expression of the zinc finger / homeobox-containing transcription factor ZFH-5 in the developing brain.<br>Knockout leads to ectopic expression of the sense RNA in the pontine nuclei and the developing cerebellar cortex, with generally stronger and more uniform expression of the sense RNA[122]. | 14:23511760-23560778<br>*ZFHX2-AS1* | • None | • Blood protein amount<br>• Level of ap-1 complex subunit gamma-like 2 in blood<br>• Level of thiamine-triphosphatase in blood<br>• Liver fat measurement<br>• Protein measurement |

**TOTAL: 61/88 = 69% of the human ortholog of functionally validated lncRNAs hit a GWAS region associated with a neural function**
Green (neural hits) = 509
Yellow (immune hits) = 177
**Ratio neural / immune hits = 2.9**
