## Supplemental Table S2 for "Frequent functional orthology of long noncoding RNAs and genomic loci associated with complex traits and disorders"

### Supplementary Table S2. GWAS correlates of lncRNAs associated with immunological functions

LncRNAs implicated in immunological processes whose orthologous positions correlate with a GWAS immunological association are highlighted in yellow. Neurological associations are highlighted in green.

| lncRNA | Reported function / phenotype | Human coordinates and possible human ortholog | Immune GWAS associations | Non-immune GWAS associations |
|---|---|---|---|---|
| 1. *AMANZI* (human) | Negatively regulates IL-1β expression and trained immunity through the induction of IL37 transcription[1]. | 2:112840328-112844195 | • Acne<br>• C-reactive protein measurement<br>• Interleukin-1 beta measurement<br>• Neutrophil count | • Glycerate measurement<br>• Lean body mass |

| | | | | |
|---|---|---|---|---|
| 2. *ANRIL / CDKN2B-AS1* (human) | Regulates cell proliferation and is regulated by TNF-α and NF-κB[2].<br><br>Cardiovascular disease associated[3]. | 9:21994139-22212895 | • Allergen exposure measurement<br>• Asthma<br>• Atopic eczema<br>• B-cell acute lymphoblastic leukemia<br>• Basophil count<br>• COVID19<br>• Crohn's disease<br>• Cytokine measurement<br>• Eosinophil count<br>• Granulocyte percentage of myeloid white cells<br>• Hemorrhoid<br>• Human papilloma virus infection<br>• IgF-1 measurement<br>• Interleukin-6 measurement<br>• Leukocyte quantity<br>• Lymphocyte count | • Abdominal aortic aneurysm<br>• Acute insulin response measurement<br>• Acute lymphoblastic leukemia<br>• Adolescent idiopathic scoliosis<br>• Age at onset<br>• Agents acting on the renin-angiotensin system use measurement<br>• Aging<br>• Alanine measurement<br>• Amount of iron in brain<br>• Aneurysm<br>• Angina pectoris<br>• Ankle brachial index<br>• Antiglaucoma preparations and miotics use measurement<br>• Antihyperlipidemic drug use measurement<br>• Antithrombotic agent use measurement<br>• Aortic aneurysm<br>• Apolipoprotein B measurement<br>• Aspartate aminotransferase measurement |

| | | | | |
|---|---|---|---|---|
| | | | • Lymphocyte percentage of leukocytes<br>• Lymphoid leukemia<br>• Monocyte count<br>• Monocyte percentage of leukocytes<br>• Multiple sclerosis<br>• Myeloid leukocyte count<br>• Neutrophil count<br>• Neutrophil percentage of leukocytes<br>• Psoriasis<br>• Rheumatoid arthritis<br>• Tumor necrosis factor ligand superfamily member 14 measurement<br>• Type 1 diabetes mellitus<br>• Wap, kazal, immunoglobulin kunitz and ntr domain-containing protein 1 measurement | • Aspartate aminotransferase to alanine aminotransferase ratio<br>• Aspirin use measurement<br>• Astrocytoma<br>• Atherosclerosis<br>• Atrial fibrillation<br>• Barrett's esophagus<br>• Basal cell carcinoma<br>• Benign neoplasm of skin<br>• Beta blocking agent use measurement<br>• Blood glucose amount<br>• Body height<br>• Body mass index<br>• Body weight<br>• Bone density<br>• Brain aneurysm<br>• Brain attribute<br>• Brain neoplasm<br>• Brain volume<br>• BRCAX breast cancer<br>• Breast cancer<br>• Breast carcinoma<br>• Breastfeeding duration<br>• Calcium channel blocker use measurement<br>• Cancer<br>• Cardiovascular disease |

| | | | | |
|---|---|---|---|---|
| | | | | • Cardiovascular disease biomarker measurement<br>• Cardioverter defibrillator<br>• Carotid atherosclerosis<br>• Carotid plaque build<br>• Cataract<br>• Central nervous system cancer<br>• Cerebral cortex area attribute<br>• Cerebrovascular disorder<br>• Chest pain<br>• Chloride amount<br>• Chronic lymphocytic leukemia<br>• Circulating fibrinogen levels<br>• Cleft lip<br>• Colorectal adenoma<br>• Colorectal cancer<br>• Congestive heart failure<br>• Coronary artery calcification<br>• Coronary artery disease<br>• Coronary atherosclerosis<br>• Cortical thickness<br>• Cup-to-disc ratio measurement<br>• Cutaneous melanoma<br>• Dental caries |

| | | | | |
|---|---|---|---|---|
| | | | | • Device complication<br>• Diabetes mellitus<br>• Diabetic eye disease<br>• Diabetic neuropathy<br>• Diabetic polyneuropathy<br>• Diabetic retinopathy<br>• Diastolic blood pressure<br>• Diastolic blood pressure change measurement<br>• Drug use measurement<br>• Drugs used in diabetes use measurement<br>• Endometrial carcinoma<br>• Endometrial neoplasm<br>• Endometriosis<br>• Erythrocyte count<br>• Erythrocyte volume<br>• Esophageal squamous cell carcinoma<br>• Estrogen-receptor negative breast cancer<br>• Exploratory eye movement measurement<br>• Facial pigmentation<br>• Factor VII measurement<br>• Factor VIII measurement<br>• Factor XI measurement<br>• Family history<br>• Female infertility |

| | | | | |
|---|---|---|---|---|
| | | | | • Fibromuscular dysplasia<br>• Gestational diabetes<br>• Glaucoma<br>• Glioblastoma multiforme<br>• Glioma<br>• Glucose homeostasis trait<br>• Glucose measurement<br>• Glutamine measurement<br>• Gut microbiome measurement<br>• Hair color<br>• Hamartoma<br>• HbA1c measurement<br>• Healthspan<br>• Heart disease<br>• Heart failure<br>• Heart rate<br>• Hematocrit<br>• Hemoglobin A1 measurement<br>• Hemoglobin measurement<br>• High density lipoprotein cholesterol measurement<br>• HMG CoA reductase inhibitor use measurement<br>• Hypertension<br>• Hypertensive heart disease<br>• Insulin measurement<br>• Insulin resistance |

| | | | | |
|---|---|---|---|---|
| | | | | • Intermediate coronary syndrome<br>• Ischemic stroke<br>• Keratinocyte carcinoma<br>• Large artery stroke<br>• Lean body mass<br>• Level of Kazal-type serine protease inhibitor domain-containing protein 1 in blood<br>• Level of serum globulin type protein<br>• Level of syndecan-4 in blood<br>• Life span trait<br>• Linoleic acid measurement<br>• Low density lipoprotein cholesterol measurement<br>• Low tension glaucoma<br>• Lung adenocarcinoma<br>• Lung cancer<br>• Lung carcinoma<br>• Macula attribute<br>• Major depressive disorder<br>• Mean corpuscular hemoglobin concentration<br>• Mean fractional anisotropy measurement<br>• Melanoma |

| | | | | |
|---|---|---|---|---|
| | | | | • Memory performance<br>• Metabolic syndrome<br>• Migraine disorder<br>• Mortality<br>• Myocardial infarction<br>• Nasopharyngeal neoplasm<br>• Neuroimaging measurement<br>• Neuropathy<br>• Nevus<br>• Non-alcoholic fatty liver disease<br>• Non-high density lipoprotein cholesterol measurement<br>• Non-melanoma skin carcinoma<br>• Non-neoplastic nevus<br>• Occlusion precerebral artery<br>• Open-angle glaucoma<br>• Optic cup area<br>• Optic disc size trait<br>• Optic nerve disorder<br>• Optic nerve glioma<br>• Optic neuritis<br>• Oral cavity cancer<br>• Ovarian carcinoma |

| | | | | |
|---|---|---|---|---|
| | | | | • Ovarian endometrioid carcinoma<br>• Ovarian serous carcinoma<br>• Pain<br>• Parental longevity<br>• Peptic ulcer and gastro-oesophageal reflux disease (GORD) drug use measurement<br>• Peripheral arterial disease<br>• Peripheral vascular disease<br>• Personality trait<br>• Pilocytic astrocytoma<br>• Plasminogen activator inhibitor 1 measurement<br>• Platelet count<br>• Platelet crit<br>• Platelet quantity<br>• Polypeptide N-acetylgalactosaminyltransferase 3 measurement<br>• Post-traumatic stress disorder<br>• Potassium measurement<br>• Prostate carcinoma<br>• Protein measurement<br>• Psychosis predisposition measurement |

| | | | | |
|---|---|---|---|---|
| | | | | • Pulse pressure measurement<br>• Receptor-type tyrosine-protein phosphatase H measurement<br>• Red cell distribution width<br>• Response to trauma exposure<br>• Reticulocyte amount<br>• Reticulocyte count<br>• Retinopathy<br>• Schizophrenia<br>• Sex hormone-binding globulin measurement<br>• Sex interaction measurement<br>• Skin cancer<br>• Skin neoplasm<br>• Spleen volume<br>• Squamous cell carcinoma<br>• Squamous cell lung carcinoma<br>• Stanniocalcin-2 measurement<br>• Strand of hair color<br>• Stroke<br>• Subarachnoid hemorrhage<br>• Systolic blood pressure<br>• Systolic heart failure |

| | | | | |
|---|---|---|---|---|
| | | | | • Thoracic aortic aneurysm<br>• Tissue plasminogen activator amount<br>• Total cholesterol measurement<br>• Trait in response to thiazide<br>• Triglyceride measurement<br>• Type 2 diabetes mellitus<br>• Type 2 diabetes nephropathy<br>• Tyrosine measurement<br>• Upper aerodigestive tract neoplasm<br>• Vasodilators used in cardiac diseases use measurement<br>• Ventricular rate measurement<br>• Vestibular Schwannoma<br>• Visual pathway disorder<br>• Von Willebrand factor quality<br>• White matter integrity<br>• White matter microstructure measurement |

| | | | | |
|---|---|---|---|---|
| 3. *AS-IL1α / Gm14023 / KR095173* (mouse) | Regulates IL-1α transcription by facilitating the recruitment of RNAP-II to the *IL-1α* locus[4]. | 2:112757497-112887384<br>*ENSG00000299339* | • Acne<br>• Allergic disease<br>• Allergic rhinitis<br>• Asthma<br>• Behcet's syndrome<br>• C-reactive protein measurement<br>• Eczematoid dermatitis<br>• Interleukin 1 receptor antagonist measurement<br>• Interleukin-1 beta measurement<br>• Interleukin-1 receptor antagonist protein measurement<br>• Leukocyte quantity<br>• Neutrophil count<br>• Neutrophil percentage of leukocytes<br>• Respiratory system disease | • Birth measurement<br>• Birth weight<br>• Corneal endothelial cell attribute<br>• Dysmenorrhea<br>• Dysmenorrheic pain measurement<br>• Endometriosis<br>• Gestational age<br>• Glycerate measurement<br>• Lean body mass<br>• Oropharynx cancer<br>• Platelet volume<br>• Prostate carcinoma<br>• Quality of life<br>• Short-term memory<br>• Sleep apnea measurement |

| | | | | |
|---|---|---|---|---|
| 4. *AVAN / XLOC_040025* (human) | Promotes antiviral innate immunity by interacting with TRIM25 and enhancing the transcription of FOXO3a[5]. | 6:108558901-108559417 | • None | • None |
| 5. *BCALM / AC099524.1* (human) | Negatively regulates B cell receptor-mediated calcium signaling[6]. | 16:81739082-81767868 | • Allergen exposure measurement | • Alzheimer disease<br>• Gut microbiome measurement<br>• Ischemic stroke<br>• Platelet volume<br>• Response to antineoplastic agent<br>• Response to carboplatin |

| | | | | |
|---|---|---|---|---|
| 6. *DLEU1* (human) | Osteoarthritis associated[7]. | 13:50081725-50906856 | • Allergen exposure measurement<br>• Allergic rhinitis<br>• Autoimmune disease<br>• Basophil count<br>• Blood immunoglobulin amount<br>• Celiac disease<br>• COVID-19<br>• Eczematoid dermatitis<br>• Eosinophil count<br>• Eosinophil percentage of granulocytes<br>• Eosinophil percentage of leukocytes<br>• Fc receptor-like protein 2 measurement<br>• Interleukin 9 measurement<br>• Leukocyte quantity<br>• Low affinity immunoglobulin epsilon | • Abnormality of the skeletal system<br>• Acute myeloid leukemia<br>• Adiponectin measurement<br>• Adolescent idiopathic scoliosis<br>• Alzheimer disease<br>• Amount of iron in brain<br>• Anterior thigh muscle volume<br>• Anthropometric measurement<br>• APOE carrier status<br>• Appendicular lean mass<br>• Ascending aorta diameter<br>• Atrial fibrillation<br>• Attention deficit hyperactivity disorder<br>• Base metabolic rate measurement<br>• Benign prostatic hyperplasia<br>• Bipolar disorder<br>• Blood phosphate measurement<br>• Blood urea nitrogen amount<br>• BMI-adjusted adiponectin measurement |

| | | | | |
|---|---|---|---|---|
| | | | Fc receptor measurement<br>• Lymphocyte amount<br>• Lymphocyte count<br>• Lymphocyte percentage of leukocytes<br>• Monocyte count<br>• Monocyte percentage of leukocytes<br>• Multiple sclerosis<br>• Osteoarthritis<br>• Psoriasis<br>• TGF-beta amount<br>• TNF-related apoptosis-inducing ligand measurement<br>• Tumor necrosis factor alpha amount | • BMI-adjusted hip circumference<br>• BMI-adjusted waist circumference<br>• BMI-adjusted waist-hip ratio<br>• Body composition measurement<br>• Body fat percentage<br>• Body height<br>• Body mass index<br>• Body surface area<br>• Body weight<br>• Bone density<br>• Breast carcinoma<br>• Breast density<br>• Breastfeeding duration<br>• Bulb of aorta size<br>• Cardiovascular disease<br>• Caudal anterior cingulate cortex volume<br>• Chronic lymphocytic leukemia<br>• Clonal hematopoiesis<br>• Coffee consumption measurement<br>• Colorectal cancer<br>• Corticoliberin measurement |

| | | | | |
|---|---|---|---|---|
| | | | | • Dentin matrix acidic phosphoprotein 1 amount<br>• Diabetes mellitus<br>• Diabetic maculopathy<br>• Diastolic blood pressure<br>• Diffuse large b-cell lymphoma<br>• Drugs used in diabetes use measurement<br>• Educational attainment<br>• Electrocardiography<br>• Endometriosis<br>• Erythrocyte count<br>• Erythrocyte volume<br>• Fatty acid amount<br>• Fear of severe pain measurement<br>• FEV/FVC ratio<br>• Forced expiratory volume<br>• Gamma-aminoisobutyric acid measurement<br>• Gestational diabetes<br>• Glomerular filtration rate<br>• Glucose measurement<br>• Glycine measurement<br>• Grip strength measurement<br>• Gut microbiome measurement<br>• HbA1c measurement |

| | | | | |
|---|---|---|---|---|
| | | | | • Heel bone mineral density<br>• Height growth attribute<br>• Hematocrit<br>• Hemoglobin measurement<br>• Hemorrhoid<br>• High density lipoprotein cholesterol measurement<br>• Hip circumference<br>• Hypertension<br>• Hypothyroidism<br>• Inguinal hernia<br>• Intelligence<br>• Lean body mass<br>• Left ventricular structural measurement<br>• Level of ribonuclease H2 subunit A in blood<br>• Lipoprotein A measurement<br>• Low density lipoprotein cholesterol measurement<br>• Lower urinary tract symptom<br>• Magnesium measurement<br>• Magnetic Resonance Imaging of the Heart<br>• Major depressive disorder |

| | | | | |
|---|---|---|---|---|
| | | | | • Matrix extracellular phosphoglycoprotein amount<br>• Metabolic syndrome<br>• Neck of femur size<br>• Neonatal abstinence syndrome<br>• Non-high density lipoprotein cholesterol measurement<br>• Number of teeth<br>• Obstructive sleep apnea<br>• Odontogenesis<br>• Oligodendroglioma<br>• Open-angle glaucoma<br>• Osteosarcoma<br>• Otosclerosis<br>• Peak expiratory flow<br>• PHF-tau measurement<br>• Platelet component distribution width<br>• Platelet count<br>• Platelet-to-lymphocyte ratio<br>• Posterior thigh muscle fat infiltration measurement<br>• Posterior thigh muscle volume<br>• Primary biliary cirrhosis |

| | | | | |
|---|---|---|---|---|
| | | | | • Prostate carcinoma<br>• Prostate specific antigen amount<br>• Protein measurement<br>• Puberty<br>• QRS duration<br>• QRS-T angle<br>• Response to antipsychotic drug<br>• Response to lithium ion<br>• Reticulocyte amount<br>• Reticulocyte count<br>• S wave amplitude<br>• Sensory perception of smell<br>• Serum alanine aminotransferase amount<br>• Serum amyloid P-component amount<br>• Serum creatinine amount<br>• Serum gamma-glutamyl transferase measurement<br>• Serum urea amount<br>• Sex hormone-binding globulin measurement<br>• Sexual dimorphism measurement<br>• Sleep apnea<br>• Smoking initiation<br>• Snoring measurement |

| | | | | |
|---|---|---|---|---|
| | | | | • Sphingomyelin 22:0 measurement<br>• Systolic blood pressure<br>• Testosterone measurement<br>• Thyroid stimulating hormone amount<br>• Total blood protein measurement<br>• Total cholesterol measurement<br>• Total hip arthroplasty<br>• Total joint arthroplasty<br>• Transmembrane protease serine 5 measurement<br>• Triglyceride measurement<br>• Triglyceride:HDL cholesterol ratio<br>• Type 2 diabetes mellitus<br>• Upper arm circumference<br>• Urate measurement<br>• Urinary albumin to creatinine ratio<br>• Uterine fibroid<br>• Uterine leiomyoma<br>• Vital capacity<br>• Waist circumference<br>• Waist-hip ratio<br>• Whole body water mass |

| | | | | |
|---|---|---|---|---|
| 7. *Dnm3os* (mouse) | Involved in macrophage polarisation[8]. | 1:172138397-172144840<br><br>*DNM3OS* | • None | • Body height |

| | | | | |
|---|---|---|---|---|
| 8. *EPIC1* (human) | Promotes tumour immune evasion and resistance to checkpoint inhibitor therapy[9]. | 22:47630827-48023004 | • Allergen exposure measurement<br>• Human papilloma virus infection<br>• Interferon gamma measurement<br>• Level of chemokine-like protein TAFA-5 in blood<br>• Monocyte percentage of leukocytes<br>• Susceptibility to mononucleosis measurement<br>• Takayasu arteritis | • 1,7-dimethylurate measurement<br>• Acute myeloid leukemia<br>• Age at onset<br>• Alzheimer disease<br>• Attention deficit hyperactivity disorder<br>• Blood insulin amount<br>• BMI-adjusted hip circumference<br>• BMI-adjusted waist-hip ratio<br>• Body height<br>• Body mass index<br>• Calvaria morphology trait<br>• Cholesteryl ester 18:0 measurement<br>• Cysteine s-sulfate measurement<br>• Delivery measurement<br>• DNA methylation<br>• Dry eye syndrome<br>• Economic and social preference<br>• Educational attainment<br>• Essential tremor<br>• Eye morphology trait<br>• Generalised epilepsy |

| | | | | |
|---|---|---|---|---|
| | | | | • Glioma pathogenesis-related protein 1 measurement<br>• Gut microbiome measurement<br>• Hematological measurement<br>• Homocysteine measurement<br>• Intelligence<br>• Lipid measurement<br>• Mathematical ability<br>• Memory performance<br>• Mixed connective tissue disease<br>• Neurofibrillary tangles measurement<br>• Obsessive-compulsive disorder<br>• Obsessive-compulsive symptom measurement<br>• Oropharynx cancer<br>• Periodontal disorder<br>• PHF-tau measurement<br>• Protein measurement<br>• Response to stimulus<br>• Schizophrenia<br>• Schizophrenia symptom severity measurement |

| | | | | |
|---|---|---|---|---|
| | | | | • Self-reported educational attainment<br>• Skin pigmentation<br>• Sleep duration trait<br>• Smoking initiation<br>• Trait in response to paliperidone<br>• Triglyceride measurement<br>• Upper aerodigestive tract neoplasm<br>• Waist-hip ratio |
| 9. *FAS-AS1* (human) | Regulates FAS receptor (CD95; TNFRSF6) signaling in B cell lymphomas FAS-AS1 binds RBM5 to block the alternative splicing of FAS pre-mRNA, which is required for the production of soluble FAS[10]. | 10:88991420-88992975 | • None | • Chronic lymphocytic leukemia<br>• Erythrocyte volume<br>• Lymphoid leukemia |

| | | | | |
|---|---|---|---|---|
| 10. *Firre* (mouse) | Regulates expression of several inflammatory genes, demonstrates NFkB-dependent expression, and interacts with hnRNPU to regulate mRNA, increases VCAM1[11].<br><br>Promotes an increase in frequency of common lymphoid progenitors and peripheral blood CD4+ and CD8+ T cells[12,13]. | X:131688779-131830928<br><br>*FIRRE* | • None | • None |
| 11. *Flatr / 2610204G07Rik* (mouse) | Promotes Treg differentiation[14]. | 5:64946134-64956710<br><br>None annotated (EST *AL701018*) | • Animal allergen seropositivity | • Central corneal thickness<br>• Diverticular disease |
| 12. *Flicr / 4930524L23Rik* (mouse) | Modulates modulates Foxp3 expression and autoimmunity; reduces Treg activity and inhibits disease in a murine model of autoimmune diabetes[15]. | X:49262866-49270521<br><br>*FLICR* | • None | • None |

| | | | | |
|---|---|---|---|---|
| 13. *Gaplinc* (mouse) | Highly expressed following macrophage differentiation. Down-regulated upon inflammatory activation. Macrophages depleted of GAPLINC display enhanced expression of inflammatory genes, while overexpression of GAPLINC suppresses this response. *Gaplinc* knockout mice display enhanced basal levels of inflammatory genes and resistance to lipopolysaccharide (LPS)-induced endotoxic shock[16]. | 18:3466200-3487049<br>*GAPLINC* | • PR interval[a] | • Breast density<br>• Citrulline measurement<br>• Major depressive disorder |
| 14. *Gas5* (mouse) | Necessary and sufficient for normal growth arrest in T-cell lines as well as human peripheral blood T cells[17].<br>Diabetic retinopathy and respiratory disease associated and involved in macrophage polarisation[18]. | 1:173851424-173868940<br>*GAS5* | • None | • Body height<br>• Erythrocyte count<br>• Hematocrit<br>• Hemoglobin measurement<br>• Intelligence<br>• Level of tenascin-N in blood<br>• Red blood cell density |

[a] PR interval in Tripanosoma cruzi seropositivity

| | | | | |
|---|---|---|---|---|
| 15. *Gm34199* (mouse) | Regulates T cell autoimmunity; augments allogeneic T-cell proliferation and TCR signaling[19,20]. | 13:74231401-74555515<br>*LINC00402 / ReLoT* | • Chronic obstructive pulmonary disease<br>• C-reactive protein measurement<br>• Cutaneous mastocytosis<br>• Cytokine measurement<br>• Interstitial lung disease<br>• Lymphocyte count<br>• Osteonecrosis<br>• Response to vaccine<br>• Salmonella seropositivity | • 1,5 anhydroglucitol measurement<br>• Abdominal fat cell number<br>• Acid sphingomyelinase-like phosphodiesterase 3a measurement<br>• Alkaline phosphatase measurement<br>• Apolipoprotein B measurement<br>• Body height<br>• Cadherin-15 measurement<br>• DNA methylation<br>• Environmental exposure measurement<br>• FEV/FVC ratio<br>• Glomerular filtration rate<br>• Glucose homeostasis trait<br>• Gut microbiome measurement<br>• Handedness<br>• Income<br>• Insulin sensitivity measurement<br>• Level of trehalase in blood<br>• Lipid measurement<br>• Low density lipoprotein cholesterol measurement |

| | | | | |
|---|---|---|---|---|
| | | | | • Male reproductive system disease<br>• Mathematical ability<br>• Non-high density lipoprotein cholesterol measurement<br>• Omega-3 polyunsaturated fatty acid measurement<br>• Optic disc size trait<br>• Phospholipid amount<br>• Platelet count<br>• Platelet volume<br>• Prostate cancer<br>• Response to antidepressant<br>• Response to glucocorticoid<br>• Restless legs syndrome<br>• Self-reported educational attainment<br>• Sex hormone-binding globulin measurement<br>• Sudden cardiac arrest<br>• Total cholesterol measurement |

| | | | | |
|---|---|---|---|---|
| 16. *H19* (mouse) | Regulates self-renewal of long-term hematopoietic stem cells[21]. | 11:1995165-2004552<br>*H19* | • Celiac disease<br>• IgF-1 measurement<br>• Type 1 diabetes mellitus | • Age at menopause<br>• Alzheimer disease<br>• Base metabolic rate measurement<br>• Birth weight<br>• BMI-adjusted hip circumference<br>• Body height<br>• Body mass index<br>• Body surface area<br>• Breast cancer<br>• Breast carcinoma<br>• Cystatin c measurement<br>• Diastolic blood pressure<br>• DNA methylation<br>• Environmental exposure measurement<br>• Family history of Alzheimer's disease<br>• Femoral neck bone mineral density<br>• Glomerular filtration rate<br>• Gut microbiome measurement<br>• HbA1c measurement<br>• High density lipoprotein cholesterol measurement<br>• Hypertension<br>• Lung cancer |

| | | | | |
|---|---|---|---|---|
| | | | | • Menstrual cycle attribute<br>• Pulse pressure measurement<br>• Pursuit maintenance gain measurement<br>• Reticulocyte count<br>• Serum creatinine amount<br>• Sex hormone-binding globulin measurement<br>• Smoking status measurement<br>• Squamous cell lung carcinoma<br>• Systolic blood pressure<br>• Tinnitus<br>• Type 2 diabetes mellitus<br>• Urate measurement<br>• Vanillactate measurement<br>• White matter hyperintensity measurement<br>• Whole body water mass |
| 17. *HCG4* (human) | Regulates RIG-I-mediated IFN production to suppress H1N1 swine influenza virus replication[22]. | 6:29791031-29793073 | • Cutaneous mastocytosis<br>• Mastocytosis<br>• Rheumatoid arthritis<br>• Takayasu arteritis | • Hypothyroidism<br>• Total cholesterol measurement |

| | | | | |
|---|---|---|---|---|
| 18. *HCP5* (human) | Diabetic retinopathy associated modulating the miR-93-5p/HMGA2 axis[23]. | 6:31463144-31494470 | • Aids<br>• Animal allergen seropositivity<br>• Ankylosing spondylitis<br>• Anti-tetanus toxoid IgG measurement<br>• Asthma<br>• Atopic asthma<br>• Basophil count<br>• Blautia seropositivity<br>• Blood immunoglobulin amount<br>• Carpal tunnel syndrome<br>• CCL5/CXCL3 protein level ratio in blood<br>• Chronic interstitial cystitis<br>• Clostridiales seropositivity<br>• Clostridium difficile infection<br>• Complement C4 measurement | • 25-hydroxyvitamin D3 measurement<br>• Abnormality of the skeletal system<br>• Age at diagnosis<br>• Aging<br>• Alcohol use disorder measurement<br>• Amount of iron in brain<br>• Angina pectoris<br>• Anilide use measurement<br>• Apolipoprotein a 1 measurement<br>• Apolipoprotein B measurement<br>• APP/DKK1 protein level ratio in blood<br>• Appendicular lean mass<br>• Arterial stiffness measurement<br>• Autism spectrum disorder<br>• Beta blocking agent use measurement<br>• Blood protein amount<br>• Blood VLDL cholesterol amount<br>• BMI-adjusted hip circumference |

| | | | | |
|---|---|---|---|---|
| | | | • COVID-19<br>• C-X-C motif chemokine 9 measurement<br>• CXCL12 measurement<br>• Cytokine measurement<br>• Enterobacter phage virus seropositivity<br>• Eosinophil count<br>• Eosinophil percentage of leukocytes<br>• Granulocyte percentage of myeloid white cells<br>• Granulysin measurement<br>• Granzyme b measurement<br>• Herpes Zoster<br>• HIV-1 infection<br>• Human papilloma virus infection<br>• Inflammatory bowel disease | • BMI-adjusted waist circumference<br>• BMI-adjusted waist-hip ratio<br>• Body height<br>• Body weight<br>• CD46/TIMP1 protein level ratio in blood<br>• CDKN2D/MANF protein level ratio in blood<br>• Cholesterol:total lipids ratio<br>• Cholesteryl ester measurement<br>• Cholesteryl esters:total lipids ratio<br>• Choline measurement<br>• Chylomicron amount<br>• Clinical treatment<br>• Coagulation factor V amount<br>• Colorectal adenoma<br>• Colorectal cancer<br>• Corneodesmosin measurement<br>• Coronary artery disease<br>• Diastolic blood pressure<br>• Drug use measurement<br>• Drug-induced agranulocytosis |

| | | | | |
|---|---|---|---|---|
| | | | • Interleukin 7 receptor subunit alpha measurement<br>• Lactobacillus phage virus seropositivity<br>• Leukocyte quantity<br>• Level of interleukin-15 in blood serum<br>• Level of MHC class I polypeptide-related sequence A in blood<br>• Level of MHC class I polypeptide-related sequence B in blood<br>• Level of T-cell surface glycoprotein CD8 alpha chain in blood serum<br>• Lymphocyte count<br>• Lymphotoxin-alpha amount<br>• Macrophage metalloelastase level<br>• Malaria | • Drug-induced liver injury<br>• Dupuytren contracture<br>• EGF/MANF protein level ratio in blood<br>• Endometriosis<br>• Epithelial discoidin domain-containing receptor 1 measurement<br>• Fatty acid amount<br>• Flavin adenine dinucleotide (FAD) measurement<br>• Forced expiratory volume<br>• Free cholesterol measurement<br>• Gallstones<br>• Glioma pathogenesis-related protein 1 measurement<br>• Glucose measurement<br>• Glutamate carboxypeptidase 2 measurement<br>• Hematological measurement<br>• Hemoglobin measurement<br>• High density lipoprotein cholesterol measurement<br>• Hodgkins lymphoma<br>• Hypertension |

| | | | | |
|---|---|---|---|---|
| | | | • MHC class I polypeptide-related sequence A measurement<br>• MHC class I polypeptide-related sequence B measurement<br>• Monocyte count<br>• Mosquito bite reaction itch intensity measurement<br>• Mosquito bite reaction size measurement<br>• Multiple sclerosis<br>• Myositis<br>• Natural cytotoxicity triggering receptor 3 measurement<br>• Neonatal systemic lupus erythematosus<br>• Polymyositis<br>• Psoriasis<br>• Response to vaccine<br>• Rheumatoid arthritis<br>• Sacroiliac arthritis | • Hypothyroidism<br>• Inguinal hernia<br>• Intermediate density lipoprotein measurement<br>• Ischemic stroke<br>• ITGB1BP2/LAT2 protein level ratio in blood<br>• LCN2/PGLYRP1 protein level ratio in blood<br>• Level of chondroadherin in blood serum<br>• Level of disintegrin and metalloproteinase domain-containing protein 9 in blood<br>• Level of epidermal growth factor-like protein 7 in blood<br>• Level of laminin subunit beta-1 in blood<br>• Level of protogenin in blood<br>• Level of tetraspanin-1 in blood<br>• Level of thioredoxin domain-containing protein 15 in blood<br>• Level of U6 snRNA-associated Sm-like protein LSm8 in blood |

| | | | | |
|---|---|---|---|---|
| | | | • Sclerosing cholangitis<br>• Seasonal allergic rhinitis<br>• Staphylococcus seropositivity<br>• Streptococcus seropositivity<br>• Susceptibility to chickenpox measurement<br>• Susceptibility to childhood ear infection measurement<br>• Susceptibility to cold sores measurement<br>• Susceptibility to mumps measurement<br>• Susceptibility to plantar warts measurement<br>• Susceptibility to pneumonia measurement<br>• Susceptibility to shingles measurement<br>• Systemic lupus erythematosus | • Linoleic acid measurement<br>• Lipid measurement<br>• Lobe attachment<br>• Low density lipoprotein cholesterol measurement<br>• Lung cancer<br>• Lung carcinoma<br>• Mean corpuscular hemoglobin<br>• Metabolic syndrome<br>• Mitochondrial DNA measurement<br>• MMR-related febrile seizures<br>• Mosaic loss of chromosome X measurement<br>• NKG2-D type II integral membrane protein amount<br>• Omega-3 polyunsaturated fatty acid measurement<br>• Omega-6 polyunsaturated fatty acid measurement<br>• Oropharynx cancer<br>• Parental longevity<br>• PHF-tau measurement<br>• Phosphoglycerides measurement<br>• Phospholipid amount |

| | | | | |
|---|---|---|---|---|
| | | | • Tonsillectomy risk measurement<br>• Tumor necrosis factor ligand superfamily member 13B amount<br>• Type 1 diabetes mellitus<br>• Ulcerative colitis | • Phospholipids:total lipids ratio<br>• Physical activity measurement<br>• Platelet aggregation<br>• Platelet count<br>• Platelet crit<br>• Platelet volume<br>• Platelet-to-lymphocyte ratio<br>• Polyunsaturated fatty acid measurement<br>• Polyunsaturated fatty acids to monounsaturated fatty acids ratio<br>• Preeclampsia<br>• Prion disease<br>• Prolow-density lipoprotein receptor-related protein 1 measurement<br>• Prostate carcinoma<br>• Protein kinase C-binding protein NELL2 measurement<br>• Protein measurement<br>• Proteoglycan 3 measurement<br>• Psychotic symptoms |

| | | | | |
|---|---|---|---|---|
| | | | | • Pulmonary surfactant-associated protein d measurement<br>• Raynaud disease<br>• Red cell distribution width<br>• Response to bezlotoxumab<br>• Response to thioamide<br>• Reticulocyte count<br>• Retinal vasculature measurement<br>• Retinoic acid receptor responder protein 2 measurement<br>• Saturated fatty acids measurement<br>• Saturated fatty acids to total fatty acids percentage<br>• Schizophrenia<br>• Seizure 6-like protein 2 measurement<br>• Serum alanine aminotransferase amount<br>• Serum creatinine amount<br>• Sex hormone-binding globulin measurement<br>• Sex interaction measurement<br>• Sialoadhesin measurement |

| | | | | |
|---|---|---|---|---|
| | | | | • Squamous cell lung carcinoma<br>• Systolic blood pressure<br>• Taste liking measurement<br>• Tenascin-X measurement<br>• Testosterone measurement<br>• Thyrotoxic periodic paralysis<br>• TNFRSF13B/TNFRSF9 protein level ratio in blood<br>• Total cholesterol measurement<br>• Triacylglycerol 50:1 measurement<br>• Triglyceride measurement<br>• Upper aerodigestive tract neoplasm<br>• Urinary system trait<br>• Visceral:abdominal adipose tissue ratio measurement<br>• Visual perception quality |

| | | | | |
|---|---|---|---|---|
| 19. *HIF1A-AS2* (human) | Cardiovascular disease associated. Forms a complex with USF1, elevating ATF2 expression and promoting the development of atherosclerotic inflammation[24]. | 14:61747038-61749089 | • None | • None |
| 20. *Hotair* (mouse) | Regulates LPS-induced cytokine expression and inflammatory response in macrophages[25].<br>Promotes LPS-Induced inflammation in cardiomyocytes (a model of septic cardiomyopathy) by enhancing PDCD4 stability[26].<br>Regulates the miR-1277-5p/SGTB axis in osteoarthritis[27]. | 12:53962308-53975055<br>*HOTAIR* | • Hemorrhoid | • BMI-adjusted waist circumference<br>• BMI-adjusted waist-hip ratio<br>• Heel bone mineral density<br>• Level of phosphatidylinositol |
| 21. *Hotairm1* (mouse) | Specifically expressed in myeloid cells and is involved in myelopoiesis through modulation of the HOXA cluster[28]. | 7:27095647-27100265<br>*HOTAIRM1* | • Multiple sclerosis<br>• Tonsillectomy risk measurement | • BMI-adjusted waist circumference<br>• BMI-adjusted waist-hip ratio |

| | | | | |
|---|---|---|---|---|
| 22. *Ifngas1 / NeST / TMEVPG1* (mouse) | Promotes Th1 lineage-specific expression of IFN-γ and regulates susceptibility to viral and bacterial pathogens[29,30].<br><br>Binds to WDR5 to mediate H3K4me3 at the *Ifng* promoter and promotes the IFN-γ expression in cis as an enhancer lncRNA in CD8+ T cells[30].<br><br>*IFNG-AS1* overexpression leads to increased IFNγ secretion in natural killer cells[31].<br><br>Associated with an IBD susceptibility risk locus and expression is increased in patients with ulcerative colitis and acute graft-versus-host disease[32].<br><br>Enhances T helper type 1 cell response in patients with Sjögren syndrome[33]. | 12:67989446-68234686<br><br>*IFNG-AS1* | • Allergen exposure measurement<br>• Alopecia areata<br>• Ankylosing spondylitis<br>• Anti-centromere-antibody-positive systemic scleroderma<br>• Basophil measurement<br>• Crohn's disease<br>• C-X-C motif chemokine 9 measurement<br>• Eosinophilic esophagitis<br>• Inflammatory bowel disease<br>• Leukocyte quantity<br>• Lymphocyte count<br>• Mosquito bite reaction itch intensity measurement<br>• Mosquito bite reaction size measurement<br>• Myeloid leukocyte count<br>• Psoriasis | • Adolescent idiopathic scoliosis<br>• Antisaccade response measurement<br>• Arthralgia<br>• Bilirubin measurement<br>• Blood lead amount<br>• Bone density<br>• Diabetic retinopathy<br>• Forced expiratory volume<br>• Gut microbiome measurement<br>• Hypothyroidism<br>• Mental or behavioural disorder<br>• Metabolic disease<br>• Myopia<br>• Perceived unattractiveness to mosquitos measurement<br>• Platelet count<br>• Platelet crit<br>• Quality of life<br>• Response to bronchodilator<br>• Sex interaction measurement<br>• Systolic blood pressure change measurement<br>• Thyroid preparation use measurement |

| | | | | |
|---|---|---|---|---|
| | | | • Rheumatoid arthritis<br>• Sclerosing cholangitis<br>• Susceptibility to plantar warts measurement<br>• Systemic lupus erythematosus<br>• Ulcerative colitis | • Wellbeing measurement |
| 23. *IL1β-RBT46* and *IL1β-eRNA* (human) | Both nuclear-localized and both favour LPS-induced transcription and release of the pro-inflammatory mediators IL-1β and CXCL8[34]. | 2:112838838-112842855 | • Interleukin-1 beta measurement | • Glycerate measurement |

| | | | | |
|---|---|---|---|---|
| 24. *INCR1 / ENSG00000286162* (human) | Inhibits T cell-mediated tumor killing and promotes tumor cell growth and division[35]. | 9:5457477-5629748 | • Eosinophil percentage of leukocytes<br>• Interleukin-1 receptor-like 2 measurement<br>• Programmed cell death 1 ligand 1 amount<br>• Programmed cell death 1 ligand 2 amount | • Age at onset<br>• Alkaline phosphatase measurement<br>• Alzheimer disease<br>• Arterial stiffness measurement<br>• Blood protein amount<br>• Cathepsin F measurement<br>• Environmental exposure measurement<br>• Epididymis-specific alpha-mannosidase measurement<br>• Female fertility<br>• Fertility trait<br>• Gut microbiome measurement<br>• Hexadecadienoate (16:2n6) measurement<br>• ICOSLG/PDCD1LG2 protein level ratio in blood<br>• Level of prorelaxin H2 in blood<br>• Level of Sphingomyelin (d34:2) in blood serum<br>• Level of sphingomyelin phosphodiesterase in blood<br>• N-acylethanolamine-hydrolyzing acid amidase measurement |

| | | | | |
|---|---|---|---|---|
| | | | | • Parental longevity<br>• Protein measurement<br>• Tobacco smoke exposure measurement |
| 25. *LIMIT* (human) | Promotes antitumor immunity and enhances checkpoint inhibitor responses by increasing the expression of MHC-I[36]. | 1:89421079-89422551 | • None | • None |
| 26. *LINC00265* (human) | Expression is increased in osteoarthritis chondrocytes, knockdown in chondrocytes inhibits inflammation, and overexpression increases inflammation[37]. | 7:39733122-39794745 | • COVID-19 | • None |

| | | | | |
|---|---|---|---|---|
| 27. *LINC00305* (human) | An atherosclerosis-associated SNP is located in the intron of *LINC00305*. *LINC00305* expression is enriched in atherosclerotic plaques and monocytes. Overexpression of LINC00305 promotes the expression of inflammation-associated genes in THP-1 cells and activates nuclear factor-kappa beta (NF-κB), inhibition of which abolishes *LINC00305*-mediated activation of cytokine expression[38]. | 18:64079989-64153377 | • None | • Gastric cancer<br>• Level of serpin B8 in blood<br>• Memory performance<br>• Peptide measurement<br>• Sex interaction measurement<br>• Vaginal microbiome measurement<br>• Waist-hip ratio |

| | | | | |
|---|---|---|---|---|
| 28. *LINC02288* (human) | Increased in osteoarthritis patient cartilage RNA sequencing data. Knockdown reduces apoptosis and inflammation of osteoarthritis chondrocytes (induced by interleukin-1β) and protects against development of pathological changes and production of inflammatory molecules in an osteoarthritis mouse model[39]. | 14:77027711-77086720 | • C-reactive protein measurement<br>• Chronic obstructive pulmonary disease | • Alcohol consumption quality<br>• Aspartate aminotransferase measurement<br>• Attention deficit hyperactivity disorder<br>• Body fat percentage<br>• Body height<br>• Body mass index<br>• Body weight<br>• Diastolic blood pressure<br>• Educational attainment<br>• Feeling nervous measurement<br>• FEV/FVC ratio<br>• Glomerular filtration rate<br>• Glycoprotein measurement<br>• Lifestyle measurement<br>• Mathematical ability<br>• Maximum cigarettes per day measurement<br>• Response to anticonvulsant<br>• Serum alanine aminotransferase amount<br>• Serum albumin amount<br>• Serum gamma-glutamyl transferase measurement<br>• Sleep quality<br>• Smoking behavior trait |

| | | | | |
|---|---|---|---|---|
| | | | | • Smoking initiation<br>• Smoking status measurement<br>• Social inhibition quality<br>• Substance abuse<br>• Systolic blood pressure<br>• Triglyceride measurement |
| 29. *LINC02574* (human) | Inhibition of LINC02574 expression in A549 cells enhances influenza A virus (IAV) replication, while overexpression of LINC02574 inhibits viral production. Knockdown attenuates the expression of type I and type III IFNs and multiple ISGs, as well as the activation of STAT1 triggered by IAV infection. LINC02574 deficiency impairs the expression of RIG-I, TLR3, and MDA5[40]. | 1:27660327-27666279 | • Leukocyte quantity<br>• Monocyte count<br>• Neutrophil count | • Body mass index<br>• Body weight<br>• Level of scavenger receptor cysteine-rich domain-containing group B protein in blood<br>• Mean arterial pressure<br>• Pulse pressure measurement<br>• S-warfarin measurement<br>• Systolic blood pressure |

| | | | | |
|---|---|---|---|---|
| 30. *Linc-MAF-4 / MAFTRR* (human) | Acts as a scaffold to recruit and modulate the enzymatic activity of EZH2 on the MAF promoter, which regulates MAF transcription and promotes a CD4+ TH1 phenotype[41].<br><br>Elevated in PBMCs of patients with acute graft-versus-host disease[42] and is associated with multiple sclerosis[43]. | 16:79605850-79770651 | • Blood immunoglobulin amount<br>• COVID-19<br>• C-reactive protein measurement<br>• Eosinophil count<br>• Eosinophil percentage of leukocytes<br>• Gout<br>• Graves disease<br>• High affinity immunoglobulin alpha and immunoglobulin mu Fc receptor measurement<br>• IgF-1 measurement<br>• Joint disease<br>• Multiple sclerosis<br>• Systemic lupus erythematosus | • Afamin measurement<br>• Age at menopause<br>• Alkaline phosphatase measurement<br>• Alpha-2-HS-glycoprotein measurement<br>• Alpha-fetoprotein amount<br>• Aspartate aminotransferase measurement<br>• Axial length measurement<br>• Bilirubin measurement<br>• Blood insulin amount<br>• Blood phosphate measurement<br>• Blood protein amount<br>• Body fat percentage<br>• Body height<br>• Breast density<br>• Carboxypeptidase M measurement<br>• Cathepsin L2 measurement<br>• Cholesterol:total lipids ratio<br>• Cholesteryl esters:total lipids ratio<br>• Coagulation factor X amount<br>• Colorectal cancer<br>• Cystatin c measurement |

| | | | | |
|---|---|---|---|---|
| | | | | • Differentiated thyroid carcinoma<br>• Glomerular filtration rate<br>• Goiter<br>• High density lipoprotein cholesterol measurement<br>• Hormone measurement<br>• Hyperthyroidism<br>• Hypothyroidism<br>• Level of angiopoietin-related protein 1 in blood serum<br>• Level of cadherin-related family member 2 in blood<br>• Level of organic solute transporter subunit beta in blood<br>• Level of thyrotropin subunit beta in blood<br>• Level of transthyretin in blood<br>• Low density lipoprotein cholesterol measurement<br>• Methionine sulfone measurement<br>• Mortality<br>• Multinodular goiter<br>• Nontoxic goiter<br>• Obesity |

| | | | | |
|---|---|---|---|---|
| | | | | • Oropharynx cancer<br>• Platelet count<br>• Platelet crit<br>• Polycystic ovary syndrome<br>• Serum alanine aminotransferase amount<br>• Serum creatinine amount<br>• Sex hormone-binding globulin measurement<br>• Testosterone measurement<br>• Thyroglobulin measurement<br>• Thyroid cancer<br>• Thyroid function<br>• Thyroid gland volume<br>• Thyroid stimulating hormone amount<br>• Thyrotoxicosis<br>• Thyroxine amount<br>• Toxic nodular goiter<br>• Trait in response to thiazide<br>• Triglyceride measurement<br>• Tyrosine measurement<br>• Urate measurement<br>• Uric acid measurement<br>• Urinary system trait<br>• Vitamin B deficiency<br>• Vitamin C measurement<br>• Vitamin D amount |

| | | | | |
|---|---|---|---|---|
| | | | | • Vitamin D-binding protein measurement<br>• Xaa-Pro aminopeptidase 2 amount<br>• Xaa-Pro aminopeptidase 2 measurement |

| | | | | |
|---|---|---|---|---|
| 31. *LincRNA-Cox2 / Ptgs2os2* (mouse) | Positive and negative regulator of immune response genes[44].<br>Regulates innate PRR pathways[45].<br>Upregulated in TLR4-stimulated cells but not TLR3-stimulated cells[46].<br>Functions as an eRNA to regulate the activity of the *COX2* gene as well as the expression of NF-kB, IL-12, iNOS, and TNF-α and several immune genes *in vivo*[47,48].<br>Regulates inflammatory gene expression by modulating SWI/SNF-mediated chromatin remodeling[49] and interacting with hnRNP-A/B and hnRNP-A2/B1, and expression is induced by TLR ligands in a MyD88- and NF-κB-dependent manner[44]. | 1:186614990-186626662<br>*ENSG00000288562* | • None | • None |

| | | | | |
|---|---|---|---|---|
| 32. *LincRNA-EPS / Ttc39aos1* (mouse) | Negatively regulates inflammatory cytokine production by binding hnRNPL and is protective against endotoxemia and Listeria monocytogenes infections in mice[50].<br><br>Associated with IRG region chromatin where it maintains chromatin in an epigenetically repressed state[47,50]. | 1:51329616-51335410<br><br>*TTC39A-AS1* | • None | • Amygdala volume<br>• Breastfeeding duration<br>• FEV/FVC ratio<br>• Gut microbiome measurement<br>• Heart failure<br>• Waist-hip ratio |
| 33. *Lnc13* (mouse) | Decreased expression and hypofunctional polymorphisms seen in patients with celiac disease[51]. | 2:102452050-102454594<br><br>*LNC13* | • Celiac disease<br>• Inflammatory bowel disease | • None |

| | | | | |
|---|---|---|---|---|
| 34. *LncATV (human)* | RIG-I antiviral signaling is enhanced by LncATV knockdown and inhibited by overexpressed lncATV[52]. | 1:234957342-234970062 | • Gout<br>• Level of T-cell immunoglobulin and mucin domain-containing protein 4 in blood<br>• Monocyte count<br>• Monocyte percentage of leukocytes | • Apolipoprotein a 1 measurement<br>• Cognitive function measurement<br>• Low density lipoprotein cholesterol measurement<br>• Omega-6 polyunsaturated fatty acid measurement<br>• Platelet count<br>• Regenerating islet-derived protein 3-alpha measurement<br>• Serum gamma-glutamyl transferase measurement<br>• Sialic acid-binding Ig-like lectin 6 amount<br>• Total cholesterol measurement |
| 35. *Lnc-DC* (human) | Transcribed from the *Wdnm1-like* pseudogene[53]. Exclusively expressed in dendritic cells (DC). Required for optimal DC differentiation from monocytes and regulates DC activation of T cells. Interacts with the transcription factor STAT3[54]. | 17:60083560-60091985 | • None | • None |

| | | | | |
|---|---|---|---|---|
| 36. *LncBAZ2B / BAZ2B-AS1* (human) | Increased in children with asthma, promotes M2 macrophage polarisation, and overexpression exacerbates cockroach allergen extract-induced lung inflammation[55]. | 2:159615292-159618637 | • Granulocyte percentage of myeloid white cells<br>• Monocyte percentage of leukocytes<br>• Neutrophil count<br>• Neutrophil percentage of leukocytes | • Blood protein amount<br>• Color vision disorder<br>• Educational attainment<br>• FEV/FVC ratio<br>• Hypothyroidism<br>• Serum gamma-glutamyl transferase measurement |
| 37. *lncBST2 / BISPR* (human) | Induced by influenza and VSV mutants that are unable to block the IFN response, and their expression increases with HCV infection and in the liver of infected patients[56]. | 19:17405686-17419324 | • CD6 measurement | • Level of bone marrow stromal antigen 2 in blood<br>• Level of bone marrow stromal antigen 2 in blood serum<br>• Level of signaling threshold-regulating transmembrane adapter 1 in blood serum<br>• Level of Src kinase-associated phosphoprotein 1 in blood<br>• Low density lipoprotein cholesterol measurement |

| | | | | |
|---|---|---|---|---|
| 38. *LncCSR-IgA* (mouse) | Deletion of lncRNA-CSR impairs IgA class switching in B cells. Ectopic expression of lncRNA-CSR rescues class switching in *lncRNA-CSR*–deficient B cells[57].<br>Deletion results in intestinal dysbiosis and mucosal inflammation[57]. | 14:101928673-101964420<br>*ENSG00000289482, ENSG00000300533* | • None | • Body mass index<br>• Protein amnionless measurement |
| 39. *Lnc-EGFR / EGILA* (human) | Promotes Treg differentiation and inhibits cytotoxic T lymphocyte-mediated killing. Increased expression correlates with increased hepatocellular carcinoma tumor size[58]. | 14:20693480-20707120 | • None | • Angiogenin measurement<br>• Blood protein amount<br>• Level of ribonuclease 4 in blood serum<br>• Memory performance<br>• Opioid dependence<br>• Psychotic symptoms<br>• Ribonuclease 4 measurement<br>• Ribonuclease K6 measurement<br>• Ribonuclease pancreatic measurement<br>• Trait in response to paclitaxel |

| | | | | |
|---|---|---|---|---|
| 40. *lnc-EPAV* (mouse) | Enhances innate immune responses via derepressing RELA expression[59]. | No alignment | • N/A | • N/A |

| | | | | |
|---|---|---|---|---|
| 41. *LncHSC1* / *AK039852* / *Gm35551* (mouse) | Regulates myeloid differentiation[47,60]. | 2:42972255-43040662<br>*LINC01819* | • Cough<br>• Drug allergy<br>• Eosinophil count<br>• Tumor necrosis factor receptor superfamily member EDAR amount | • Agents acting on the renin-angiotensin system use measurement<br>• Aspartate aminotransferase measurement<br>• Balding measurement<br>• Birth weight<br>• Body height<br>• Bone density<br>• Cardiovascular disease<br>• Carotid artery thickness<br>• Corneal resistance factor<br>• Diastolic blood pressure<br>• Erythrocyte volume<br>• Glucose tolerance test<br>• Glucose-dependent insulinotropic peptide measurement<br>• Heel bone mineral density<br>• Hematological measurement<br>• High density lipoprotein cholesterol measurement<br>• Hypertension<br>• Insomnia<br>• Mean corpuscular hemoglobin<br>• Memory performance |

| | | | | |
|---|---|---|---|---|
| | | | | • N-acetyltaurine measurement<br>• Neuroticism measurement<br>• Platelet count<br>• Platelet volume<br>• Pregnancy disorder<br>• Prostate carcinoma<br>• Prostate specific antigen amount<br>• Pulse pressure measurement<br>• Red cell distribution width<br>• Response to angiotensin-converting enzyme inhibitor<br>• S-6-hydroxywarfarin measurement<br>• Sex interaction measurement<br>• Skin disease<br>• Smoking status measurement<br>• Solar lentigines measurement<br>• Systolic blood pressure<br>• Thyroid stimulating hormone amount<br>• Trait in response to platinum |

| | | | | |
|---|---|---|---|---|
| | | | | • Type 2 diabetes mellitus<br>• Waist-hip ratio<br>• White matter hyperintensity measurement |

| | | | | |
|---|---|---|---|---|
| 42. *LncHSC2 / 4930519L02Rik* (mouse) | Involved in HSC self-renewal and T cell differentiation, and recruits the hematopoietic transcription factor E2A to its binding sites[60]. | 1:87620803-88685450<br>*PKN2-AS1* | • C-C motif chemokine 2 level<br>• CCL3 measurement<br>• Clostridium difficile infection<br>• COVID-19<br>• CXCL9 measurement<br>• Cytokine measurement<br>• Irritable bowel syndrome<br>• Microglial activation attribute<br>• Periodontitis<br>• Prostate specific antigen amount<br>• Response to tnf antagonist<br>• Streptococcus seropositivity<br>• Susceptibility to childhood ear infection measurement | • Abnormality of refraction<br>• Adenosine deaminase measurement<br>• Adolescent idiopathic scoliosis<br>• Age at first birth measurement<br>• Age at first sexual intercourse measurement<br>• Age-related hearing impairment<br>• Alcohol consumption quality<br>• Alcoholic liver disease<br>• Anxiety disorder measurement<br>• Aspartate aminotransferase measurement<br>• Aspartate aminotransferase to alanine aminotransferase ratio<br>• Attention deficit hyperactivity disorder<br>• Autism spectrum disorder<br>• Autosomal dominant compelling helio-ophthalmic outburst syndrome |

| | | | | |
|---|---|---|---|---|
| | | | • Tumor necrosis factor receptor superfamily member 9 amount | • Benign prostatic hyperplasia<br>• Bladder calculus<br>• Blood sodium bicarbonate amount<br>• BMI-adjusted waist circumference<br>• Body height<br>• Body mass index<br>• Body surface area<br>• Brain attribute<br>• Brain connectivity attribute<br>• Brain volume<br>• Breast carcinoma<br>• Cannabis dependence measurement<br>• Cerebral cortex area attribute<br>• Cognition<br>• Color vision disorder<br>• Corneal resistance factor<br>• Coronary artery disease<br>• Cortical thickness<br>• Dementia<br>• Descending aorta diameter<br>• Diastolic blood pressure<br>• Diet measurement<br>• Diffuse plaque measurement |

| | | | | |
|---|---|---|---|---|
| | | | | • Educational attainment<br>• Erythrocyte count<br>• Fat pad mass<br>• Gait quality<br>• Glaucoma<br>• Glioma pathogenesis-related protein 1 measurement<br>• Glomerular filtration rate<br>• Glutamate measurement<br>• Gut microbiome measurement<br>• Hematocrit<br>• Hemoglobin measurement<br>• Hypertension<br>• Imidazole lactate measurement<br>• Insomnia<br>• Intraocular pressure measurement<br>• Level of guanylate-binding protein 1 in blood serum<br>• Level of guanylate-binding protein 4 in blood<br>• Lifestyle measurement<br>• Lipid measurement<br>• Mean arterial pressure<br>• Memory performance |

| | | | | |
|---|---|---|---|---|
| | | | | • Narcolepsy-cataplexy syndrome<br>• Neuroimaging measurement<br>• Neuropsychological test<br>• Nicotine dependence symptom count<br>• Obsessive-compulsive disorder<br>• Open-angle glaucoma<br>• Parental genotype effect measurement<br>• Peripheral arterial disease<br>• Platelet component distribution width<br>• Platelet count<br>• Platelet crit<br>• Post-traumatic stress disorder<br>• Prostate carcinoma<br>• Protein measurement<br>• Pulse pressure measurement<br>• Red blood cell density<br>• Response to angiotensin-converting enzyme inhibitor<br>• Risk-taking behaviour |

| | | | | |
|---|---|---|---|---|
| | | | | • Self-reported educational attainment<br>• Serum alanine aminotransferase amount<br>• Serum creatinine amount<br>• Serum gamma-glutamyl transferase measurement<br>• Serum metabolite level<br>• Serum ST2 amount<br>• Size<br>• Sleep apnea measurement<br>• Smoking cessation<br>• Smoking initiation<br>• Social inhibition quality<br>• Substance abuse<br>• Suicide behaviour<br>• Systolic blood pressure<br>• Tobacco smoke exposure measurement<br>• Traffic air pollution measurement<br>• Trait in response to paclitaxel<br>• Type 2 diabetes mellitus<br>• Vaginal microbiome measurement<br>• Vertigo<br>• Vitamin D amount |

| | | | | |
|---|---|---|---|---|
| 43. *Lnc-IL7R* (human) | Overlaps the 3'UTR of IL7 protein-coding gene. Regulates the expression of the inflammatory mediators IL-6, IL-8, E-selectin, and VCAM-1 by deposition of H3K27me3 at the promoters[61]. | 5:35878189-35879601 | • Allergic disease<br>• Allergic rhinitis<br>• Asthma<br>• CD48 antigen measurement<br>• Eczematoid dermatitis<br>• Interleukin 7 receptor subunit alpha measurement<br>• Lymphocyte count<br>• Multiple sclerosis | • None |
| 44. *lncISG15 / ENSG00000224969* (human) | Induced by influenza and VSV mutants that are unable to block the IFN response, and expression increases with HCV infection[56]. | 1:1011997-1013193 | • None | • None |
| 45. *LncITPRIP-1 / CFAP58-DT* (human) | Binds to the C-terminus of MDA5 (melanoma differentiation-associated protein 5) and promotes its oligomerization to enhance IFN signaling[62]. | 10:104351296-104353685 | • None | • Platelet volume<br>• Uterine fibroid |

| | | | | |
|---|---|---|---|---|
| 46. *LncKdm2b / A930024E05Rik* (mouse) | Promotes ILC3 maintenance and proliferation, enhances the effector functions of ILC3s, and protects against intestinal bacterial infection in a murine model[63]. | 12:121580360-121597226<br>*KDM2B-DT* | • C-reactive protein measurement | • Alzheimer disease<br>• Hematological measurement<br>• Left ventricular mass index<br>• Platelet volume<br>• Polygenic risk score |
| 47. *Lnc-Lsm3b* (mouse) | Inactivates RIG-1 innate activity and type I IFN production[64]. | 3:14181556-14182069<br>None annotated | • None | • None |
| 48. *LncNSPL / LOC105370355* (human) | Highly expressed in monocytes in patients infected with influenza A virus. LncNSPL overexpression in mice increases the susceptibility to IAV infection and impaired IFN-I production. *LncNSPL* binds to RIG-I and blocks the interaction between RIG-I and E3 ligase, reducing RIG-I ubiquitination and limiting the downstream production of antiviral mediators during the late stage of IAV infection[65]. | 13:108328029-108365281 | • B-cell antigen receptor complex-associated protein beta chain measurement<br>• Granulocyte percentage of myeloid white cells<br>• Leukocyte quantity<br>• Lymphocyte:monocyte ratio<br>• Monocyte count<br>• Monocyte percentage of leukocytes<br>• Neutrophil count | • Blood protein amount<br>• Diffuse plaque measurement<br>• Fas apoptotic inhibitory molecule 3 measurement<br>• Isolated dystonia<br>• Level of serum globulin type protein<br>• Platelet count<br>• Venous thromboembolism |

| | | | | |
|---|---|---|---|---|
| 49. *LncRNA-CCL2 / ENSG00000301139* (human) | Regulates CCL2 expression in endothelial cells[66]. | 17:34219125-34252520 | • Ankylosing spondylitis<br>• C-C motif chemokine 13 level<br>• C-C motif chemokine 7 level<br>• C-C motif chemokine 8 level<br>• CCL11 measurement<br>• Crohn's disease<br>• Inflammatory bowel disease<br>• Monocyte percentage of leukocytes<br>• Psoriasis<br>• Sclerosing cholangitis<br>• Ulcerative colitis | • Corneodesmosin measurement |

| | | | | |
|---|---|---|---|---|
| 50. *LincR-Ccr2-5'AS* (mouse) | Knockdown decreases the expression of chemokine receptor genes and compromises the migration of $T_H2$ cells to lung tissues[67]. | 3:46282974-46317963<br>*ENSG00000288724 / ENSG00000296106 / UQCRC2P1* | • C-C motif chemokine 13 level<br>• C-C motif chemokine 2 level<br>• Celiac disease<br>• COVID-19<br>• Granulocyte percentage of myeloid white cells<br>• Lymphocyte amount<br>• Lymphocyte:monocyte ratio<br>• Monocyte count<br>• Monocyte percentage of leukocytes<br>• Neutrophil percentage of leukocytes<br>• Oral ulcer | • Brain attribute<br>• Response to antidepressant |
| 51. *lncRNA-CMPK2 / NRIR* (human) | Has a negative regulatory role in the modulation of the IFN response.<br>Regulates the expression and protein release of CXCL10 and CCL8 and demonstrates type I IFN-dependent expression[68,69]. | 2:6819463-6840464 | • Bronchial disease | • Cognitive function measurement<br>• Depressive symptom measurement<br>• Pancreatic carcinoma<br>• Smoking status measurement |

| | | | | |
|---|---|---|---|---|
| 52. *LncRNA-FA2H-2* (human) | Silencing activates inflammation and inhibited autophagy flux in endothelial and smooth muscle cells[70]. | 16:74869397-74873372 | • None | • None |
| 53. *LncRNA-GM / Mxd4 3' UTR* (human) | Promotes the antiviral innate immune response[71]. | 4:2247432-2249343 | • None | • High density lipoprotein cholesterol measurement |
| 54. *LncRNA-ISIR / Gm31208 / Gm57827 / A330040F15Rik* (mouse) | Protects against viral infection by enhancing interferon production in viral infection and autoinflammation[72].<br><br>Expression is increased in peripheral blood mononuclear cells of patients with systemic lupus erythematosus and correlates with disease severity[72]. | 11:59268876-59284033 *ENSG00000214797 / AK131315* | • None | • Breastfeeding duration<br>• Conotruncal heart malformations<br>• Gut microbiome measurement |

| | | | | |
|---|---|---|---|---|
| 55. *LncRNA-MAP3K4 / 4732491K20Rik* (mouse) | Regulates vascular inflammation via the p38 MAPK signaling pathway[73]. | 6:160990318-160992342<br><br>*MAP3K4-AS1* | • C-X-C motif chemokine 9 measurement<br>• Cytokine measurement<br>• Periodontitis<br>• Susceptibility to childhood ear infection measurement | • Aging<br>• Angiostatin measurement<br>• Blood protein amount<br>• Blood VLDL cholesterol amount<br>• Body height<br>• Cation-independent mannose-6-phosphate receptor measurement<br>• Cholesterol:total lipids ratio<br>• Cholesteryl ester measurement<br>• Cholesteryl esters:total lipids ratio<br>• Chylomicron amount<br>• Clinical treatment<br>• Cognitive decline measurement<br>• Coronary artery disease<br>• Degree of unsaturation measurement<br>• Diabetic nephropathy<br>• Docosahexaenoic acid to total fatty acids percentage<br>• Fatty acid amount<br>• Free cholesterol measurement<br>• Free cholesterol:total lipids ratio |

| | | | | |
|---|---|---|---|---|
| | | | | • Glioblastoma multiforme<br>• High density lipoprotein cholesterol measurement<br>• Level of fibroblast growth factor-binding protein 3 in blood<br>• Level of hypoxia up-regulated protein 1 in blood<br>• Level of protein-arginine deiminase type-4 in blood<br>• Linoleic acid measurement<br>• Lipid measurement<br>• Lipoprotein A measurement<br>• Low density lipoprotein cholesterol measurement<br>• Myocardial infarction<br>• Oligodendroglioma<br>• Omega-3 polyunsaturated fatty acid measurement<br>• Omega-6 polyunsaturated fatty acid measurement<br>• Parental longevity<br>• Phospholipid amount<br>• Phospholipids:total lipids ratio<br>• Plasma plasminogen measurement<br>• Plasminogen activator inhibitor 1 measurement |

| | | | | |
|---|---|---|---|---|
| | | | | • Polyunsaturated fatty acid measurement<br>• Polyunsaturated fatty acids to monounsaturated fatty acids ratio<br>• Polyunsaturated fatty acids to total fatty acids percentage<br>• Protein measurement<br>• S-(5-Adenosy)-L-homocysteine measurement<br>• S-6-hydroxywarfarin measurement<br>• Saturated fatty acids measurement<br>• Saturated fatty acids to total fatty acids percentage<br>• Smoking initiation<br>• Splenic disease<br>• Total cholesterol measurement<br>• Triglyceride measurement<br>• Triglycerides to phosphoglycerides ratio<br>• Triglycerides:total lipids ratio<br>• Uterine leiomyoma<br>• VLDL particle size |

| | | | | |
|---|---|---|---|---|
| 56. *lincRNA-Tnfaip3 / ENSG00000299400* (mouse) | Acts as a coactivator of NF-κB for the transcription of inflammatory genes in innate immune cells through modulation of epigenetic chromatin remodeling[74]. | 6:137817503-137861008<br>*lincRNA-TNFAIP3 / ENSG00000299400* | • Allergen exposure measurement<br>• Allergic rhinitis<br>• Asthma<br>• Eczematoid dermatitis<br>• Eosinophil count<br>• Multiple sclerosis<br>• Psoriasis<br>• Systemic lupus erythematosus | • Hypothyroidism<br>• Gut microbiome measurement<br>• Neuropathic pain<br>• Total blood protein measurement |

| | | | | |
|---|---|---|---|---|
| 57. *Lnc-Smad3* / *Gm16759* (mouse) | Negatively regulates Treg differentiation[75]. | 15:67290636-67521898<br><br>*IQCH-AS1* | • Chronic obstructive pulmonary disease<br>• Crohn's disease<br>• Diverticular disease<br>• Inflammatory bowel disease<br>• Response to peginterferon alfa-2a | • 25-hydroxyvitamin D3 measurement<br>• Abdominal aortic aneurysm<br>• Age at menarche<br>• Bipolar disorder<br>• BMI-adjusted hip circumference<br>• BMI-adjusted waist circumference<br>• Body fat percentage<br>• Body height<br>• Body mass index<br>• Bone density<br>• Corneal resistance factor<br>• Erythrocyte count<br>• Forced expiratory volume<br>• Hypertension<br>• Insomnia<br>• Insomnia measurement<br>• Left ventricular diastolic function measurement<br>• Metabolic syndrome<br>• Neuroticism measurement<br>• Risk-taking behaviour<br>• Serum gamma-glutamyl transferase measurement<br>• Taste liking measurement<br>• Vital capacity |

| | | | | |
|---|---|---|---|---|
| | | | | • Waist-hip ratio |
| 58. *Lnc-UC* / *Lncucc1* (mouse) | Decreases inflammatory signaling and prevents colitis in mouse models[76]. | 5:169052253-169052777<br>*EST R07572* | • None | • None |
| 59. *Lnczc3h7a* (mouse) | Promotes TRIM25-mediated RIG-I antiviral innate immune response[77]. | 16:11760963-11761401<br>None annotated | • None | • None |

| | | | | |
|---|---|---|---|---|
| 60. *Loup* (mouse) | Regulates *SPI1* influencing myeloid differentiation and knockdown upregulates NFkB-targeted genes[78]. | 11:47381509-47409369<br><br>*SLC39A13-AS1 / LOUP* | • Level of interleukin-12 subunit beta in blood<br>• Neutrophil count | • Alcohol consumption quality<br>• Alcohol use disorder measurement<br>• Apolipoprotein a 1 measurement<br>• Blood urea nitrogen amount<br>• Body height<br>• Body mass index<br>• Chronic kidney disease<br>• Collectin-12 measurement<br>• C-type lectin domain family 5 member A measurement<br>• Diastolic blood pressure<br>• Electrocardiography<br>• Erythrocyte count<br>• Glomerular filtration rate<br>• Glucose measurement<br>• Hematocrit<br>• Hypertension<br>• Insomnia<br>• Level of C-type lectin domain family 4 member C in blood<br>• Level of sarcoplasmic reticulum histidine-rich calcium-binding protein in blood |

| | | | | |
|---|---|---|---|---|
| | | | | • Level of seizure 6-like protein in blood<br>• Level of V-set and immunoglobulin domain-containing protein 4 in blood<br>• Level of Xaa-Pro dipeptidase in blood<br>• Longitudinal alcohol consumption measurement<br>• Mean fractional anisotropy measurement<br>• Neuroticism measurement<br>• Non-lobar intracerebral hemorrhage<br>• Omega-6 polyunsaturated fatty acid measurement<br>• PHF-tau measurement<br>• Platelet count<br>• Platelet crit<br>• Prostate carcinoma<br>• Serum albumin amount<br>• Serum creatinine amount<br>• Systolic blood pressure<br>• Urate measurement |

| | | | | |
|---|---|---|---|---|
| 61. *LOXL1-AS1* (human) | Upregulated in osteoarthritis patient cartilage and silencing in chondrocytes reduced inflammation[79]. | 15:73898471-73928296 | • Amount of CD276 antigen (human) in blood<br>• Carpal tunnel syndrome | • Anthropometric measurement<br>• Aortic measurement<br>• Atrial fibrillation<br>• BMI-adjusted waist circumference<br>• Body height<br>• Body mass index<br>• Descending aorta diameter<br>• Educational attainment<br>• Exfoliation syndrome<br>• Feeling "fed-up" measurement<br>• Insomnia<br>• Metabolic syndrome<br>• Open-angle glaucoma<br>• Sexual dimorphism measurement |
| 62. *LRIR / ENST00000514933 / GCLC* (human) | Knockdown promotes IAV replication; overexpression inhibits viral replication[80]. | 6:53514335-53523928 | • C-reactive protein measurement | • Diet measurement<br>• Lung adenocarcinoma |

| | | | | |
|---|---|---|---|---|
| 63. *MaIL1 / MAILR* (human) | Highly expressed in LPS-activated macrophages. Stabilizes the TLR4 signaling-protein OPTN and is required for TBK1-kinase–dependent IRF3 transcription factor phosphorylation and immune gene activation[81]. | 8:102864029-103002424 | • Leukocyte quantity<br>• Lymphocyte percentage of leukocytes<br>• Myeloid leukocyte count<br>• Neutrophil count<br>• Neutrophil percentage of leukocytes | • 10-nonadecenoate 19:1n9 measurement<br>• Agents acting on the renin-angiotensin system use measurement<br>• Alzheimer disease<br>• Amygdala volume<br>• Apolipoprotein a 1 measurement<br>• Ascorbic acid 2-sulfate measurement<br>• Bone density<br>• Breast density<br>• Cerebrospinal fluid composition attribute<br>• Diastolic blood pressure<br>• Environmental exposure measurement<br>• Erythrocyte count<br>• Erythrocyte volume<br>• Female infertility<br>• Gut microbiome measurement<br>• Hematocrit<br>• High density lipoprotein cholesterol measurement<br>• JT interval<br>• Mean corpuscular hemoglobin concentration |

| | | | | |
|---|---|---|---|---|
| | | | | • Mean reticulocyte volume<br>• Metabolic syndrome<br>• N-acetylputrescine measurement<br>• PHF-tau measurement<br>• Phospholipids:total lipids ratio<br>• Platelet volume<br>• PR interval<br>• QT interval<br>• Reticulocyte amount<br>• Schizophrenia<br>• Schizophrenia symptom severity measurement<br>• Serum selenium amount<br>• Sex interaction measurement<br>• TPE interval measurement<br>• Trait in response to paliperidone<br>• Type 2 diabetes mellitus |

| | | | | |
|---|---|---|---|---|
| 64. *Malat1* (mouse) | Negatively regulates innate antiviral responses in myeloid and CD4+ T cells and promotes immune tolerance[82,83].<br><br>Promotes immune tolerogenic dendritic cells that augment Treg expansion and anti-inflammatory cytokine production[84].<br><br>Regulates LPS-mediated M1 macrophage activation and IL-4-mediated M2 differentiation [47,85]. Interacts p50/p65 subunit to inhibit NF-kB DNA binding activity[86]. | 11:65497606-65508073<br><br>*MALAT1* | • Lymphocyte count<br>• Osteoarthritis<br>• Periodontitis | • BMI-adjusted hip circumference<br>• Body mass index<br>• Bone density<br>• Fat pad mass<br>• Femoral neck bone mineral density<br>• Glaucoma<br>• Heart rate<br>• High density lipoprotein cholesterol measurement<br>• Level of latent-transforming growth factor beta-binding protein 3 in blood<br>• Metabolic syndrome<br>• Schizophrenia symptom severity measurement<br>• Sex hormone-binding globulin measurement<br>• Total joint arthroplasty<br>• Trait in response to paliperidone<br>• Triglyceride measurement<br>• Uric acid measurement<br>• Visceral adipose tissue quantity<br>• Waist-hip ratio |

| | | | | |
|---|---|---|---|---|
| 65. *MEG3* (human) | Promotes proliferation and inhibits apoptosis in osteoarthritis chondrocytes[87].<br>Promotes fibrosis and inflammatory response in diabetic nephropathy[88].<br>Regulates IL-1β abundance in lung infection[89]. | 14:100779206-100861031 | • IgF-1 measurement<br>• Type 1 diabetes mellitus | • 2,6-dihydroxybenzoic acid measurement<br>• Birth weight<br>• Blood cobalt amount<br>• Blood protein amount<br>• Body height<br>• Breast carcinoma<br>• Breastfeeding duration<br>• Chymotrypsinogen B measurement<br>• Delta-like protein 1 measurement<br>• Diastolic blood pressure change measurement<br>• Emphysema imaging measurement<br>• Environmental exposure measurement<br>• Forced expiratory volume<br>• Glucose measurement<br>• Gut microbiome measurement<br>• HbA1c measurement<br>• Health trait<br>• HETE measurement<br>• Inactive pancreatic lipase-related protein 1 measurement<br>• Lean body mass |

| | | | | |
|---|---|---|---|---|
| | | | | • Level of phospholipase A2 in blood<br>• Optic disc size trait<br>• Peripheral arterial disease<br>• Protein delta homolog 1 measurement<br>• Response to diuretic<br>• Size<br>• Traffic air pollution measurement<br>• Trypsin-2 measurement<br>• Type 2 diabetes mellitus<br>• Vascular cell adhesion protein 1 amount |

| | | | | |
|---|---|---|---|---|
| 66. *MIR3142HG* (human) | Regulates CCL2 and IL-8 and demonstrates NF-kB-dependent expression[90]. | 5:160438594-160600965 | • Allergic disease<br>• Allergic rhinitis<br>• Asthma<br>• Blood immunoglobulin amount<br>• COVID-19<br>• Dermatomyositis<br>• Eczematoid dermatitis<br>• Eosinophil count<br>• Eosinophil percentage of leukocytes<br>• Juvenile dermatomyositis<br>• Myositis<br>• Respiratory system disease<br>• Response to tnf antagonist<br>• Rheumatoid arthritis<br>• Sjogren syndrome<br>• Systemic lupus erythematosus | • Acute myeloid leukemia<br>• Adolescent idiopathic scoliosis<br>• Body mass index<br>• Color vision disorder<br>• Inhalant adrenergic use measurement<br>• Level of tryptase alpha/beta-1 in blood<br>• Skin pigmentation<br>• Wellbeing measurement |

| | | | | |
|---|---|---|---|---|
| | | | • Systemic scleroderma | |
| 67. *Mirt2* (mouse) | Regulates macrophage polarization and aberrant inflammatory activity.<br>Inhibits the K63-ubiquitination of TRAF6[91]. | 8:144010951-144013413<br>None annotated | • Tonsillectomy risk measurement | • Circulating fibrinogen levels<br>• Venous thromboembolism |

| | | | | |
|---|---|---|---|---|
| 68. *mNAIL / Gm16685 / IL7-AS* (mouse) | Expression is increased in patients with ulcerative colitis and exacerbates murine model of colitis[92].<br><br>IL7-AS negatively regulates IL-6 release[90,93].<br><br>IL-7–AS promotes the expression of several inflammatory genes, including CCL2, CCL5, CCL7, and IL-6, in cells in response to LPS. It interacts with p300 to regulate histone acetylation levels around the promoter regions of these gene loci. IL-7–AS and p300 complex modulate the assembly of SWI/SNF complex to the promoters[94]. | 8:78805179-78956082<br><br>*MITA1 / hNAIL / IL7-AS / LOC105375914* | • COVID-19<br>• Eosinophil count<br>• Hpv seropositivity<br>• Interleukin 7 measurement<br>• Lymphocyte count<br>• Mosquito bite reaction size measurement<br>• Staphylococcus seropositivity | • Adolescent idiopathic scoliosis<br>• Age at onset<br>• Attempted suicide<br>• Blood protein amount<br>• Body height<br>• Bone density<br>• Breastfeeding duration<br>• Cardiomyopathy<br>• Color vision disorder<br>• Creutzfeldt jacob disease<br>• Diabetic ketoacidosis<br>• Environmental exposure measurement<br>• Exostosis<br>• Frontotemporal dementia<br>• Glioblastoma multiforme<br>• Gut microbiome measurement<br>• Inferior temporal gyrus volume<br>• Intelligence<br>• Jaw disease<br>• Neurofibrillary tangles measurement<br>• PHF-tau measurement<br>• Platelet count<br>• Platelet-to-lymphocyte ratio |

| | | | | |
|---|---|---|---|---|
| | | | | • Protein measurement<br>• Retinal drusen<br>• Schizophrenia<br>• Self-reported educational attainment<br>• Total cholesterol measurement<br>• X-12063 measurement |

| | | | | |
|---|---|---|---|---|
| 69. *Morrbid / Gm14005* (mouse) | Promotes myeloid cell survival, promotes apoptotic death of infected lymphoid cells, and regulates CD8+ T cell survival and function[83,95,96].<br><br>Expression is elevated in patients with hypereosinophilic syndromes[95]. | 2:111006015-111523376<br><br>*MORRBID / MIR4435-2HG* | • ACPA-positive rheumatoid arthritis<br>• Adult onset asthma<br>• Allergic disease<br>• Allergic rhinitis<br>• Alopecia areata<br>• Amount of CD160 antigen (human) in blood<br>• Amount of natural killer cells antigen CD94 (human) in blood<br>• Anti-hepatitis E virus antibody measurement<br>• Asthma<br>• Atopic asthma<br>• Atopic eczema<br>• Basophil count<br>• Basophil measurement<br>• Basophil percentage of granulocytes<br>• Basophil percentage of leukocytes | • 4-androsten-3beta,17beta-diol disulfate 1 measurement<br>• Age at menopause<br>• Alkaline phosphatase measurement<br>• Appendicular lean mass<br>• Aspartate aminotransferase measurement<br>• Beta-1,3-galactosyl-O-glycosyl-glycoprotein beta-1,6-N-acetylglucosaminyltransferase measurement<br>• Blood protein amount<br>• BMI-adjusted hip circumference<br>• BMI-adjusted waist circumference<br>• BMI-adjusted waist-hip ratio<br>• Body height<br>• Body mass index<br>• Body weight<br>• Bone density<br>• Breast carcinoma<br>• Calcium measurement<br>• Cancer<br>• Central corneal thickness |

| | | | | |
|---|---|---|---|---|
| | | | • B-cell receptor CD22 level<br>• CD160/CXCL16 protein level ratio in blood<br>• CD160/IL15 protein level ratio in blood<br>• CD5 antigen-like measurement<br>• Childhood onset asthma<br>• Chronic obstructive pulmonary disease<br>• Churg-Strauss syndrome<br>• COVID-19<br>• C-X-C motif chemokine 16 measurement<br>• Eczematoid dermatitis<br>• Eosinophil count<br>• Eosinophil percentage of granulocytes<br>• Eosinophil percentage of leukocytes<br>• Fc receptor-like protein 1 measurement | • Cholesterol:total lipids ratio<br>• Cholesteryl ester measurement<br>• Cholesteryl esters:total lipids ratio<br>• Chronic lymphocytic leukemia<br>• Cleft lip<br>• Cognitive function measurement<br>• Corneal endothelial cell attribute<br>• Corneal resistance factor<br>• Cystatin c measurement<br>• Dehydroisoandrosterone sulfate DHEA-S measurement<br>• Diacylglycerol 36:2 measurement<br>• Diastolic blood pressure<br>• Disorder of pharynx<br>• Drugs used in diabetes use measurement<br>• Endothelial cell-specific molecule 1 measurement<br>• Erythrocyte attribute<br>• Erythrocyte count<br>• Erythrocyte volume<br>• Fatty acid amount |

| | | | | |
|---|---|---|---|---|
| | | | • Fc receptor-like protein 3 measurement<br>• Fc receptor-like protein 5 measurement<br>• Granulocyte percentage of myeloid white cells<br>• Granulocyte-macrophage colony-stimulating factor measurement<br>• IgF-1 measurement<br>• IL15/KLRD1 protein level ratio in blood<br>• IL15/NCR1 protein level ratio in blood<br>• Immunoglobulin isotype switching attribute<br>• Immunoglobulin J chain measurement<br>• Inflammatory bowel disease<br>• Interleukin 12 measurement | • FEV/FVC ratio<br>• Free androgen index<br>• Free cholesterol measurement<br>• Free cholesterol:total lipids ratio<br>• Glomerular filtration rate<br>• Gut microbiome measurement<br>• HbA1c measurement<br>• Heart rate<br>• Heel bone mineral density<br>• Hematocrit<br>• Hematological measurement<br>• Hemoglobin A1 measurement<br>• Hemoglobin measurement<br>• High density lipoprotein cholesterol measurement<br>• Hip circumference<br>• Hodgkins lymphoma<br>• Hormone measurement<br>• Immature reticulocyte measurement<br>• Lean body mass<br>• Level of coiled-coil domain-containing glutamate-rich protein 2 (human) in blood |

| | | | | |
|---|---|---|---|---|
| | | | • Interleukin-2 receptor subunit beta measurement<br>• Interleukin-5 receptor subunit alpha measurement<br>• Killer cell lectin-like receptor subfamily F member 1 level<br>• Leukocyte immunoglobulin-like receptor subfamily A member 4 measurement<br>• Leukocyte quantity<br>• Level of interleukin-12 subunit beta in blood<br>• Level of interleukin-15 in blood serum<br>• Level of killer cell lectin-like receptor subfamily B member 1 in blood serum<br>• Level of lymphocyte antigen 96 in blood | • Level of creatine kinase U-type mitochondrial in blood<br>• Level of C-type lectin domain family 4 member C in blood<br>• Level of serum globulin type protein<br>• Lipid measurement<br>• L-Selectin measurement<br>• Lymphoid leukemia<br>• Lymphotactin measurement<br>• Lymphotoxin-alpha amount<br>• Mean corpuscular hemoglobin<br>• Mean corpuscular hemoglobin concentration<br>• Mean reticulocyte volume<br>• Moderate albuminuria<br>• Multiple myeloma<br>• NKG2-D type II integral membrane protein amount<br>• Non-Hodgkins lymphoma<br>• Omega-6 polyunsaturated fatty acid measurement<br>• Ovarian carcinoma<br>• Ovarian serous adenocarcinoma<br>• Ovarian serous carcinoma |

| | | | | |
|---|---|---|---|---|
| | | | • Level of MHC class I polypeptide-related sequence A in blood<br>• Level of MHC class I polypeptide-related sequence B in blood<br>• Lymphocyte amount<br>• Lymphocyte count<br>• Lymphocyte percentage of leukocytes<br>• Lymphocyte:monocyte ratio<br>• Monocyte count<br>• Monocyte percentage of leukocytes<br>• Multiple sclerosis<br>• Natural cytotoxicity triggering receptor 1 measurement<br>• Natural killer cell receptor 2B4 measurement<br>• Neutrophil count | • Phospholipids:total lipids ratio<br>• Platelet component distribution width<br>• Platelet count<br>• Platelet crit<br>• Platelet-to-lymphocyte ratio<br>• Polyunsaturated fatty acids to monounsaturated fatty acids ratio<br>• Polyunsaturated fatty acids to total fatty acids percentage<br>• Potassium measurement<br>• Prostate cancer<br>• Prostate carcinoma<br>• Prostate specific antigen amount<br>• Protein measurement<br>• Proteinuria<br>• Red blood cell density<br>• Red cell distribution width<br>• Reticulocyte amount<br>• Reticulocyte count<br>• Retinal vasculature measurement<br>• Retinopathy |

| | | | | |
|---|---|---|---|---|
| | | | • Neutrophil measurement<br>• Neutrophil percentage of granulocytes<br>• Neutrophil percentage of leukocytes<br>• Peritonsillar abscess<br>• Respiratory system disease<br>• Rheumatoid arthritis<br>• Rheumatoid factor seropositivity measurement<br>• Sclerosing cholangitis<br>• Severe acute respiratory syndrome<br>• Systemic lupus erythematosus<br>• T-lymphocyte surface antigen Ly-9 level<br>• Tonsillitis<br>• Ulcerative colitis<br>• Vasculitis | • Semaphorin-7A measurement<br>• Serum alanine aminotransferase amount<br>• Serum albumin amount<br>• Serum gamma-glutamyl transferase measurement<br>• Sex hormone-binding globulin measurement<br>• Size<br>• SLAM family member 7 measurement<br>• SLAM family member 8 measurement<br>• Smoking status measurement<br>• Systolic blood pressure<br>• Tenascin measurement<br>• Testosterone measurement<br>• Total blood protein measurement<br>• Total cholesterol measurement<br>• Trait in response to warfarin<br>• Triacylglycerol 50:4 measurement<br>• Triglyceride measurement |

| | | | | |
|---|---|---|---|---|
| | | | | • Triglyceride:HDL cholesterol ratio<br>• Triglycerides in IDL measurement<br>• Type 2 diabetes mellitus<br>• Upper respiratory tract disorder<br>• Urate measurement<br>• Uric acid measurement<br>• Urinary albumin to creatinine ratio<br>• Vascular cell adhesion protein 1 amount<br>• Visual perception quality<br>• Vitiligo<br>• Waist-hip ratio |

| | | | | |
|---|---|---|---|---|
| 70. *Neat1* (mouse) | Binds to SFPQ to translocate it from the IL-8 promoter region to paraspeckles, which results in activation of IL-8 in the TLR3-p38 signaling pathway[97].<br><br>Inflammation is reduced in Neat1$^{-/-}$ mouse models of peritonitis and pneumonia[98].<br><br>Involved in inhibiting HIV-1 replication and is upregulated during HIV-1 infection[99]. | 11:65422774-65445540<br><br>*NEAT1 / VINC* | • None | • Abnormality of the skeletal system<br>• Acute myeloid leukemia<br>• BMI-adjusted hip circumference<br>• Diastolic blood pressure<br>• Family history<br>• Hemoglobin A1 measurement<br>• Level of sorting nexin-15 in blood<br>• N6,N6,N6-trimethyllysine measurement<br>• N6,N6-dimethyllysine measurement<br>• Schizophrenia<br>• Serum alanine aminotransferase amount<br>• Sex hormone-binding globulin measurement<br>• Systolic blood pressure |

| | | | | |
|---|---|---|---|---|
| 71. *NKILA* (human) | Induced by LPS, TNF-α, and IL-1β; binds to the NF-κB/IkB complex and represses NF-κB signalling and inflammation; overall survival is decreased in breast cancer patients with >30% of tumour infiltrating lymphocytes expressing high levels of *NKILA*[100]. | 20:57710156-57712780 | • Allergen exposure measurement<br>• Arthritis<br>• Asparaginase hypersensitivity<br>• Asthma exacerbation measurement<br>• Hand, foot and mouth disease | • Adolescent idiopathic scoliosis<br>• Alkaline phosphatase measurement<br>• Attention deficit hyperactivity disorder<br>• Cancer<br>• Cortical thickness<br>• Educational attainment<br>• Energy intake<br>• Facial width measurement<br>• Growth rate attribute<br>• Gut microbiome measurement<br>• Heel bone mineral density<br>• Illness severity status<br>• Lipid measurement<br>• Lysophosphatidylethanolamine 17:0 measurement<br>• Major depressive disorder<br>• Memory performance<br>• Neurofibrillary tangles measurement<br>• Overall survival<br>• Pain measurement<br>• PHF-tau measurement<br>• Protein measurement<br>• Reaction time measurement |

| | | | | |
|---|---|---|---|---|
| | | | | • Response to bevacizumab<br>• Response to fenofibrate<br>• Serum metabolite level<br>• Sex interaction measurement<br>• Smoking initiation<br>• Smoking status measurement<br>• Sphingomyelin measurement<br>• Vital capacity |
| 72. *NRAV* (human) | NRAV is downregulated during infection with influenza virus and negatively regulates the initial transcription of IFN-stimulated genes[101]. | 12:120488079-120495954 | • None | • None |
| 73. *Nron / Gm56436* (mouse) | Prevents the activation of T cells and promotes HIV latency[83,102,103].<br>Expression of *NRON* is reduced with HIV-1 infection and then enhances HIV-1 replication through increased NFAT and viral LTR activity[102,103]. | 9:126408041-126408410<br>*NRON* | • None | • None |

| | | | | |
|---|---|---|---|---|
| 74. *PACER / PACERR* (human) | Activates COX-2 expression by occluding NF-κB complexes in primary human mammary epithelial cells and PMA-stimulated human monocyte–macrophage cells[104]. | 1:186680108-186683165 | • Mosquito bite reaction itch intensity measurement<br>• Mosquito bite reaction size measurement<br>• Osteoarthritis | • Amino acid measurement<br>• Blood urea nitrogen amount<br>• Breast carcinoma<br>• Breastfeeding duration<br>• Glomerular filtration rate<br>• Gut microbiome measurement<br>• Handedness<br>• Hodgkins lymphoma<br>• Serum creatinine amount<br>• Serum metabolite level<br>• Taurolithocholate 3-sulfate measurement<br>• Urate measurement |
| 75. *PTPRE-AS1 / ENSG00000232259* (human) | PTPRE-AS1 deficiency enhances IL-4-mediated M2 macrophage activation by regulating PTPRE-dependent signalling, and accelerated pulmonary allergic inflammation while reducing chemical-induced colitis[105]. | 10:127929376-127934517 | • None | • None |

| | | | | |
|---|---|---|---|---|
| 76. *Pvt1* (mouse) | *PVT1* knockdown inhibits the immunosuppressive function of myeloid-derived suppressor cells[106].<br><br>*PVT1* maintains glycolytic levels, glycolytic capacity under stress and ECAR/OCR ratios during T cell activation. *PVT1* depletion decreases $CD4^{+}$ T cell infiltration and disease progression in Sjögren's syndrome-like NOD/Ltj mice[107].<br><br>Enhances Treg function, prolongs allograft survival, and alleviates graft rejection[108]. | 8:127794513-128188202<br><br>*PVT1* | • Age of onset of childhood onset asthma<br>• Allergic disease<br>• Allergic rhinitis<br>• Allergic sensitization measurement<br>• Ankylosing spondylitis<br>• Asthma<br>• Autoimmune thyroid disease<br>• Basophil count<br>• Bell's palsy<br>• Childhood onset asthma<br>• C-reactive protein measurement<br>• Crohn's disease<br>• Eczematoid dermatitis<br>• Eosinophil count<br>• Eosinophil percentage of granulocytes<br>• Eosinophil percentage of leukocytes | • APOL1 risk genotype carrier status<br>• Appendicular lean mass<br>• Basal cell carcinoma<br>• Base metabolic rate measurement<br>• Birth weight<br>• Blood protein amount<br>• BMI-adjusted waist circumference<br>• Body height<br>• Body mass index<br>• Body surface area<br>• Bone density<br>• Breast cancer<br>• Breast carcinoma<br>• Calcium measurement<br>• Central nervous system non-hodgkin lymphoma<br>• Chronic kidney disease<br>• Chronic lymphocytic leukemia<br>• Clear cell renal carcinoma<br>• Cognitive decline measurement<br>• Colorectal cancer<br>• Diastolic blood pressure<br>• Diffuse large b-cell lymphoma |

| | | | | |
|---|---|---|---|---|
| | | | • Gastritis<br>• Leukocyte quantity<br>• Lymphocyte amount<br>• Lymphocyte count<br>• Monocyte count<br>• Multiple sclerosis<br>• Myeloid leukocyte count<br>• Neutrophil count<br>• Neutrophil measurement<br>• Neutrophil percentage of granulocytes<br>• Neutrophil percentage of leukocytes<br>• Psoriasis<br>• Respiratory system disease<br>• Sclerosing cholangitis<br>• Selective IgA deficiency disease<br>• Sporadic amyotrophic lateral sclerosis | • Donor genotype effect measurement<br>• Energy intake<br>• Erythrocyte attribute<br>• Erythrocyte count<br>• Erythrocyte volume<br>• Facial morphology trait<br>• Facial pigmentation<br>• Gestational age<br>• Glioma pathogenesis-related protein 1 measurement<br>• Gut microbiome measurement<br>• HbA1c measurement<br>• Hematocrit<br>• Hematological measurement<br>• Hemoglobin measurement<br>• High density lipoprotein cholesterol measurement<br>• High grade ovarian serous adenocarcinoma<br>• Hodgkins lymphoma<br>• Hyperthyroidism<br>• Hypothyroidism<br>• Inhalant adrenergic use measurement<br>• Lean body mass |

| | | | | |
|---|---|---|---|---|
| | | | • Susceptibility to scarlet fever measurement<br>• Systemic lupus erythematosus<br>• Tonsillectomy risk measurement<br>• Type 1 diabetes mellitus<br>• Ulcerative colitis | • Left ventricular systolic function measurement<br>• Level of serum globulin type protein<br>• Lung adenocarcinoma<br>• Lung carcinoma<br>• Lymphatic system cancer<br>• Lymphoma<br>• Major depressive disorder<br>• Major depressive episode<br>• Mean corpuscular hemoglobin<br>• Mean corpuscular hemoglobin concentration<br>• Multiple myeloma<br>• Neoplasm of mature b-cells<br>• Neuroblastoma<br>• Non-Hodgkins lymphoma<br>• Non-melanoma skin carcinoma<br>• Ovarian carcinoma<br>• Ovarian serous carcinoma<br>• PHF-tau measurement<br>• Phosphatidylcholine 40:6 measurement<br>• Platelet count<br>• Platelet crit<br>• Platelet-to-lymphocyte ratio |

| | | | | |
|---|---|---|---|---|
| | | | | • Primary biliary cirrhosis<br>• Prostate carcinoma<br>• Prostate specific antigen amount<br>• Psychosis<br>• Pulse pressure measurement<br>• Renal carcinoma<br>• Renal cell carcinoma<br>• Renal transplant outcome measurement<br>• Reticulocyte amount<br>• Reticulocyte count<br>• RS-10-hydroxywarfarin measurement<br>• RS-10-hydroxywarfarin to RS-warfarin ratio measurement<br>• Size<br>• Skin water amount<br>• Suicide<br>• Systolic blood pressure<br>• Thyroid preparation use measurement<br>• Total blood protein measurement<br>• Vitamin D deficiency<br>• Waist-hip ratio<br>• Whole body water mass |

| | | | | |
|---|---|---|---|---|
| 77. *RP11-86H7.1 / SLC44A3-AS1* (human) | Upregulated in human bronchial epithelial cells after exposure to traffic-related air pollution particulate matter 2.5 (TRAPM2.5) and induces inflammation, likely by activating the NF-κB signaling pathway[109]. | 1:94585556-94855426 | • Allergen exposure measurement<br>• Hemorrhoid<br>• Lymphocyte count<br>• Neutrophil count | • Blood coagulation trait<br>• Body height<br>• Chemotherapy-induced alopecia<br>• Cognitive function measurement<br>• D dimer measurement<br>• Dementia<br>• Glioblastoma multiforme<br>• Gut microbiome measurement<br>• Intelligence<br>• Myocardial infarction<br>• Prothrombin time measurement<br>• QRS duration<br>• Response to 4'-epidoxorubicin<br>• Response to 5-fluorouracil<br>• Response to cyclophosphamide<br>• Smoking status measurement<br>• Spontaneous coronary artery dissection<br>• Tissue factor measurement<br>• Type 2 diabetes mellitus<br>• Vision disorder |

| | | | | |
|---|---|---|---|---|
| 78. *Rroid* / *Ak083360* / *Gm36723* (mouse) | Controls lineage identity, homeostasis, and function of group 1 ILCs[110]. | 2:7922425-8595477<br>*LINC00299* | • Age of onset of asthma<br>• Age of onset of childhood onset asthma<br>• Allergen exposure measurement<br>• Allergic disease<br>• Allergic rhinitis<br>• Allergic sensitization measurement<br>• Asthma<br>• Atopic asthma<br>• Atopic eczema<br>• Atopic march<br>• Basophil count<br>• Basophil percentage of granulocytes<br>• Basophil percentage of leukocytes<br>• Childhood onset asthma<br>• Chronic obstructive pulmonary disease<br>• Eczematoid dermatitis | • 1,3-dimethylurate measurement<br>• 5-acetylamino-6-amino-3-methyluracil measurement<br>• Adolescent idiopathic scoliosis<br>• Age at onset<br>• Amyloid-beta measurement<br>• Antiphospholipid syndrome<br>• Antipsychotic drug related weight gain<br>• Appendicitis<br>• Attention deficit hyperactivity disorder<br>• Body height<br>• Bone density<br>• Breastfeeding duration<br>• Cognitive decline measurement<br>• Depressive symptom measurement<br>• Educational attainment<br>• Electrocardiography<br>• Erythrocyte volume<br>• Executive function measurement<br>• FEV/FVC ratio<br>• Forced expiratory volume |

| | | | | |
|---|---|---|---|---|
| | | | • Eosinophil count<br>• Eosinophil percentage of leukocytes<br>• Granulocyte percentage of myeloid white cells<br>• Leukocyte quantity<br>• Lymphocyte amount<br>• Lymphocyte count<br>• Monocyte count<br>• Monocyte percentage of leukocytes<br>• Mosquito bite reaction itch intensity measurement<br>• Mosquito bite reaction size measurement<br>• Multiple sclerosis<br>• Myeloid leukocyte count<br>• Neutrophil measurement<br>• Neutrophil percentage of granulocytes | • Freckles<br>• Gastroesophageal reflux disease<br>• Glioblastoma multiforme<br>• Glioma pathogenesis-related protein 1 measurement<br>• Gut microbiome measurement<br>• HbA1c measurement<br>• Hematological measurement<br>• Hemoglobin A1 measurement<br>• Hypothyroidism<br>• Inhalant adrenergic use measurement<br>• Level of serum globulin type protein<br>• Life span trait<br>• Low density lipoprotein cholesterol measurement<br>• Mean corpuscular hemoglobin concentration<br>• Memory performance<br>• Neuritic plaque measurement<br>• Neurofibrillary tangles measurement |

| | | | | |
|---|---|---|---|---|
| | | | • Respiratory system disease<br>• Response to tnf antagonist<br>• Seasonal allergic rhinitis<br>• Selective IgA deficiency disease<br>• Serum IgE amount<br>• Soluble triggering receptor expressed on myeloid cells 2 measurement<br>• Susceptibility to infectious disease measurement<br>• Virologic response measurement | • Oligodendroglioma<br>• Opioid dependence<br>• Paraxanthine measurement<br>• Physical activity<br>• Primary biliary cirrhosis<br>• Prostate carcinoma<br>• Protein measurement<br>• QRS duration<br>• Receptor-type tyrosine-protein phosphatase H measurement<br>• Reticulocyte amount<br>• Reticulocyte count<br>• Retinal drusen<br>• S-6-hydroxywarfarin measurement<br>• Schizophrenia<br>• Sialic acid-binding Ig-like lectin 6 amount<br>• Sleep quality<br>• Smoking initiation<br>• Social inhibition quality<br>• Social risk factor<br>• Substance abuse<br>• Thyroid preparation use measurement<br>• Total cholesterol measurement |

| | | | | |
|---|---|---|---|---|
| | | | | • Trait in response to abacavir<br>• Trait in response to efavirenz<br>• Type 2 diabetes mellitus<br>• Vaginal microbiome measurement<br>• Venous thromboembolism<br>• Visual perception quality<br>• X-11315 measurement<br>• X-11470 measurement |
| 79. *Snhg1* (mouse) | Promotes CD8+ memory T cell differentiation and inhibits CD8+ effector differentiation in response to viral infection[111]. | 11:62851833-62856444<br>*SNHG1* | • None[b] | • None |

[b] Interacts with Vps13D which is associated with immune GWAS traits response to cytokine, interleukin-6 measurement and atrophic gastritis.

| | | | | |
|---|---|---|---|---|
| 80. *SNHG14* (human) | Expression promotes inflammatory response induced by cerebral ischemia/reperfusion injury[112].<br><br>Down-regulation protects against LPS invoked acute lung injury[113].<br><br>Inhibition alleviates LPS invoked inflammation in chondrocytes[114]. | 15:24978583-25420336 | • Asthma<br>• Inflammatory biomarker measurement | • Alzheimer disease<br>• Atrophic macular degeneration<br>• Blood osmolality<br>• Bone density<br>• Family history of Alzheimer's disease<br>• Generalized anxiety disorder<br>• Glioblastoma multiforme<br>• Myopia<br>• Neuroticism measurement<br>• Panic disorder<br>• PHF-tau measurement<br>• Prostate carcinoma<br>• RS-10-hydroxywarfarin measurement<br>• S-methylcysteine sulfoxide measurement<br>• X-24812 measurement<br>• YKL40 measurement |

| | | | | |
|---|---|---|---|---|
| 81. *SNHG16* (human) | Implicated in the autoimmune disease Myasthenia gravis[115].<br>Inhibition reduced apoptosis and inflammation in the sepsis-induced acute lung injury model[116] and correlates with acute respiratory distress syndrome occurrence and prognosis in sepsis patients[117].<br>Promotes pulmonary fibrosis[118]. | 17:76557191-76714384 | • COVID-19<br>• Leukocyte quantity<br>• Level of paired immunoglobulin-like type 2 receptor beta in blood<br>• Level of T-cell immunoglobulin and mucin domain-containing protein 4 in blood<br>• Neutrophil count<br>• Paired immunoglobulin-like type 2 receptor alpha measurement<br>• Severe acute respiratory syndrome<br>• TNFRSF1A/TNFRSF1B protein level ratio in blood | • Adolescent idiopathic scoliosis<br>• Amyloid-beta measurement<br>• Blood protein amount<br>• Bone density<br>• Cell growth regulator with EF hand domain protein 1 measurement<br>• Central corneal thickness<br>• Colorectal cancer<br>• Corneal resistance factor<br>• Diastolic blood pressure<br>• Heart rate<br>• Hepatocyte growth factor activator amount<br>• Level of extracellular glycoprotein lacritin in blood<br>• Level of matrix-remodeling-associated protein 7 in blood serum<br>• Level of meprin A subunit beta in blood<br>• Level of protein FAM3C in blood<br>• Macular telangiectasia type 2 |

| | | | | |
|---|---|---|---|---|
| | | | | • Matrix-remodeling-associated protein 7 measurement<br>• Memory performance<br>• N-acetylneuraminate measurement<br>• Plasma serine protease inhibitor measurement<br>• Protein measurement<br>• Pulse pressure measurement<br>• QT interval<br>• Reaction time measurement<br>• Reticulocyte count<br>• Retinal vasculature measurement<br>• Sex interaction measurement |
| 82. *SNHG5* (human) | Decreased expression in chronic obstructive pulmonary patients. Decreased in cells treated with cigarette smoke extract with increased inflammatory markers, which was mitigated by SNHG5 overexpression[119]. | 6:85644369-85678943 | • C-X-C motif chemokine 10 measurement<br>• Myeloid leukocyte count<br>• Neutrophil count | • 5'-nucleotidase measurement<br>• Body height<br>• Heel bone mineral density<br>• Level of carbonic anhydrase 14 in blood |

| | | | | |
|---|---|---|---|---|
| 83. *Spehd* (mouse) | Promotes multilineage differentiation and homeostasis[120]. | 3:128488130-128539314<br><br>*GATA2-AS1* | • Basophil measurement<br>• Basophil percentage of leukocytes<br>• Eosinophil count<br>• Lymphocyte count<br>• Lymphocyte:monocyte ratio<br>• Monocyte count<br>• Monocyte percentage of leukocytes<br>• Neutrophil count<br>• Neutrophil-to-lymphocyte ratio | • Aging<br>• Chromosome telomeric region length<br>• Diastolic blood pressure<br>• Epigenetic status<br>• Exploratory eye movement measurement<br>• Hematological measurement<br>• Neurofibrillary tangles measurement<br>• Pit and fissure surface dental caries<br>• Platelet count<br>• Platelet crit<br>• Prostate carcinoma<br>• Systolic blood pressure<br>• Thrombocytopenia<br>• Varicose veins |
| 84. *SμGLT* (mouse) | Regulates class switch recombination, tuned by $m^6A$ modifications[121]. | 14:105860527-105862032<br><br>None annotated | • None | • None |

| | | | | |
|---|---|---|---|---|
| 85. *TH2-LCR* (mouse) | Regulates the expression of Th2 cytokines[122]. | 5:132629735-132664272<br><br>*TH2LCRR* | • Adult onset asthma<br>• Allergic disease<br>• Allergic rhinitis<br>• Allergic sensitization measurement<br>• Alopecia areata<br>• Asthma<br>• Asthma exacerbation measurement<br>• Atopic asthma<br>• Atopic eczema<br>• Childhood onset asthma<br>• Crohn's disease<br>• Cutaneous psoriasis measurement<br>• Eczematoid dermatitis<br>• Eosinophil count<br>• Eosinophil percentage of leukocytes<br>• Erythematosquamous dermatosis<br>• Lymphocyte count | • Cardiovascular disease<br>• Diabetic ketoacidosis<br>• Glucocorticoid use measurement<br>• Hodgkins lymphoma<br>• Inhalant adrenergic use measurement<br>• Mortality<br>• Myoclonus<br>• Rosacea severity measurement<br>• Serum IgE amount<br>• Sex interaction measurement<br>• Type 2 diabetes mellitus |

| | | | | |
|---|---|---|---|---|
| | | | • Mosquito bite reaction itch intensity measurement<br>• Mosquito bite reaction size measurement<br>• Neutrophil percentage of granulocytes<br>• Neutrophil percentage of leukocytes<br>• Psoriasis<br>• Psoriasis vulgaris<br>• Psoriatic arthritis<br>• Respiratory system disease | |
| 86. *THRIL* (human) | Forms a functional transcriptional activating lncRNA-hnRNPL complex to regulate TNF-α[123]. | 12:125025434-125027410 | • None | • None |

| | | | | |
|---|---|---|---|---|
| 87. *UCA1* (human) | Overexpression contributed to the gastric cancer cell immune escape[124].<br><br>Upregulated in psoriatic skin samples. Silencing decreased inflammatory cytokine secretion and innate immunity gene expression in keratinocyte cell line HaCaT[125]. | 19:15828206-15836328 | • Ecosanoids measurement | • Alpha-CMBHC glucuronide measurement<br>• Aspartate aminotransferase measurement<br>• Autonomic nervous system disease<br>• Blood protein amount<br>• Cleft lip<br>• Coronary artery disease<br>• Gamma-CEHC glucuronide measurement<br>• Gamma-CEHC measurement<br>• Heel bone mineral density<br>• Level of N-acetylmuramoyl-L-alanine amidase in blood<br>• Level of oxylipin in blood plasma<br>• Level of sterol ester in blood serum<br>• Metabolite measurement<br>• Octadecadienedioate measurement<br>• Peripheral neuropathy<br>• Protein measurement<br>• Urinary metabolite measurement<br>• Waist circumference<br>• X-11905 measurement |

| | | | | |
|---|---|---|---|---|
| 88. *UMLILO* (human) | Involved in trained immunity. Promotes the epigenetic priming of CXCL8, CXCL1, CXCL2 and CXCL3[126]. | 4:73707725-73734688 | • C-X-C motif chemokine 6 level<br>• Interleukin-8 measurement | • Growth-regulated alpha protein measurement<br>• Serum albumin amount |

| | | | | |
|---|---|---|---|---|
| 89. *VIN / LINC01191* (human) | Induced by H1N1, H3N2, H7N7, and VSV but not by IBV or IFN-β. Knockdown by RNA interference restricts influenza A virus replication and viral protein synthesis[127]. | 2:113970621-114028083 | • Acute graft vs. host disease<br>• Allergen exposure measurement | • Aspartate aminotransferase measurement<br>• Bilirubin measurement<br>• BMI-adjusted waist circumference<br>• Chronic kidney disease<br>• Coagulation factor V amount<br>• Colorectal cancer<br>• Corneal endothelial cell attribute<br>• Diabetic nephropathy<br>• Diet measurement<br>• Donor genotype effect measurement<br>• Gut microbiome measurement Environmental exposure measurement<br>• Gestational age<br>• Hormone replacement therapy<br>• Memory performance<br>• Retinal drusen<br>• Serum alanine aminotransferase amount<br>• Serum gamma-glutamyl transferase measurement |

| | | | | |
|---|---|---|---|---|
| | | | | • Sex hormone-binding globulin measurement<br>• Systolic blood pressure<br>• Treatment-resistant hypertension |

**59/89 = 66.3% hit a GWAS region associated with an immune function**

Immune = 554

Green neural = 260

**Immune / neural = 2.13**
