## Supplementary material for "Frequent functional orthology of long noncoding RNAs and genomic loci associated with complex traits and disorders": Fig. S

### Supplementary Figures

#### Figure S1

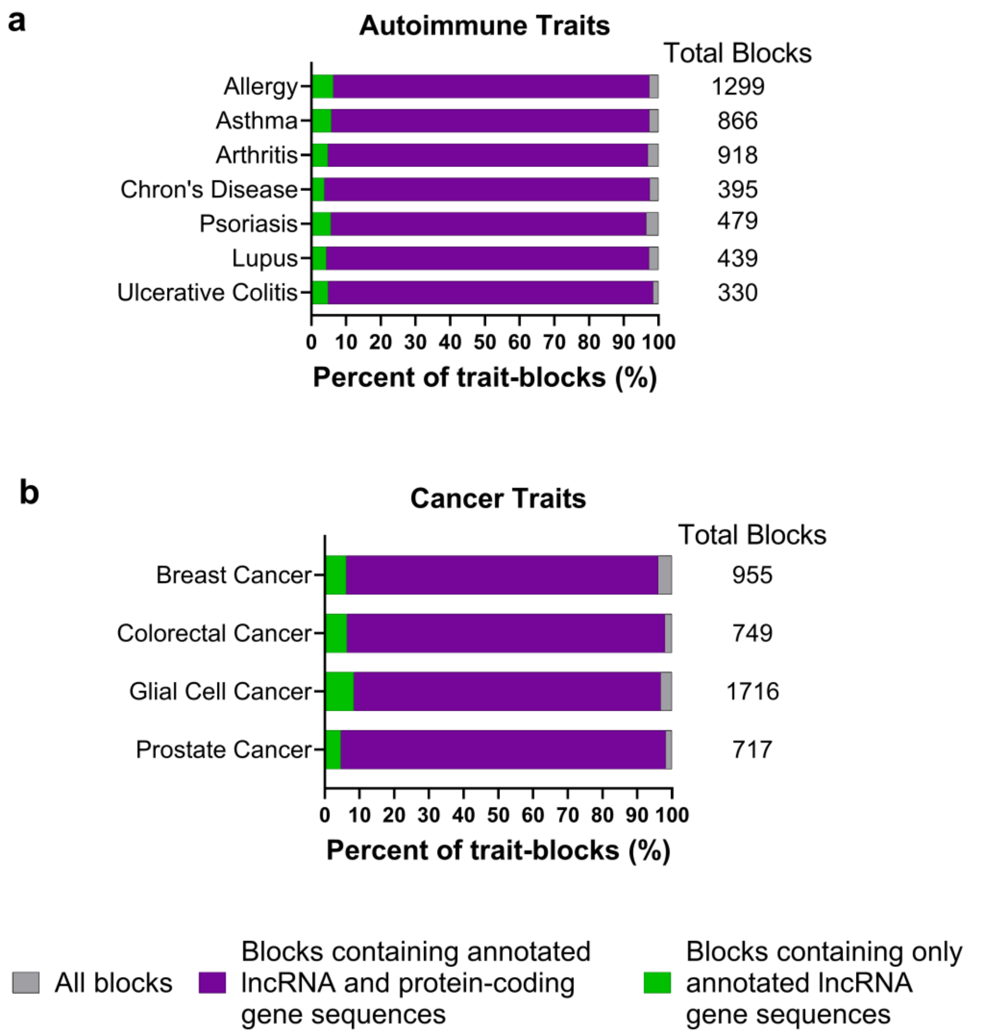


**Figure S1. Summary of haplotype blocks containing lncRNAs in autoimmune disorders and cancer.** Percentages of haplotype blocks containing lncRNAs for (**a**) autoimmune disorders and (**b**) cancers. Bars of block types are superimposed over one another.

#### Figure S2

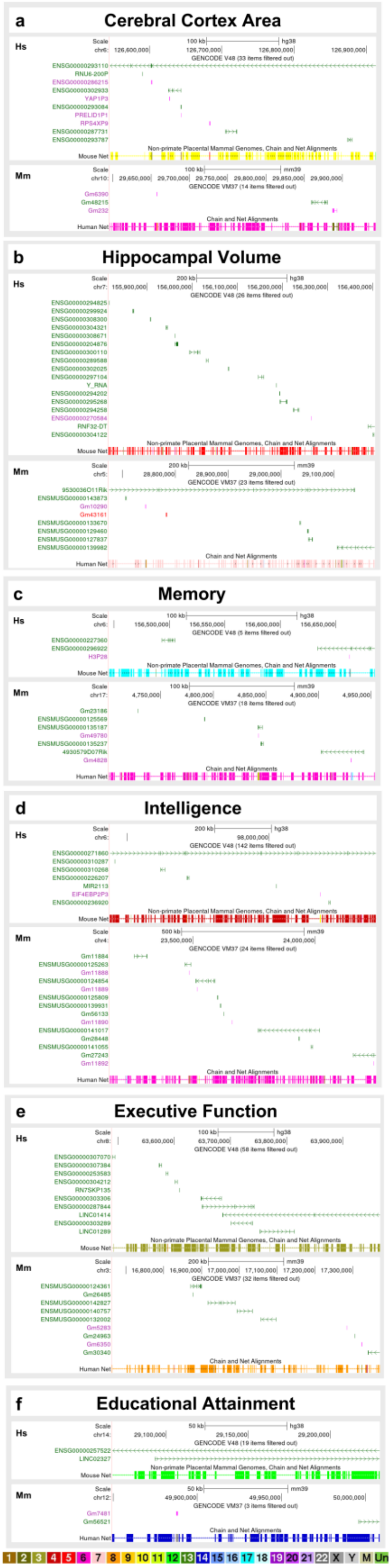


**Figure S2. UCSC genome browser view of haplotype blocks associated with brain structure and cognitive traits by GWAS.** (**a**) A haplotype block containing 55 GWAS associations with cerebral cortex area and the mouse syntenic locus. (**b**) A haplotype block containing 6 GWAS associations with hippocampal volume and the mouse syntenic locus. (**c**) A haplotype block containing 41 GWAS associations with memory and the mouse syntenic locus. (**d**) A haplotype block containing 94 GWAS associations with intelligence and the mouse syntenic locus. (**e**) A haplotype block containing 5 GWAS associations with executive function and the mouse syntenic locus. (**f**) A haplotype block containing 16 GWAS associations with educational attainment and the mouse syntenic locus. Chronograph of chromosome number applies to all browser images. Gene names: Green, annotated non-coding; pink, pseudogene; red, problem transcripts (retained introns, to be experimentally confirmed, or disrupted domains); blue, coding. Hs, *Homo sapiens*. Mm, *Mus musculus*.

Figure S3

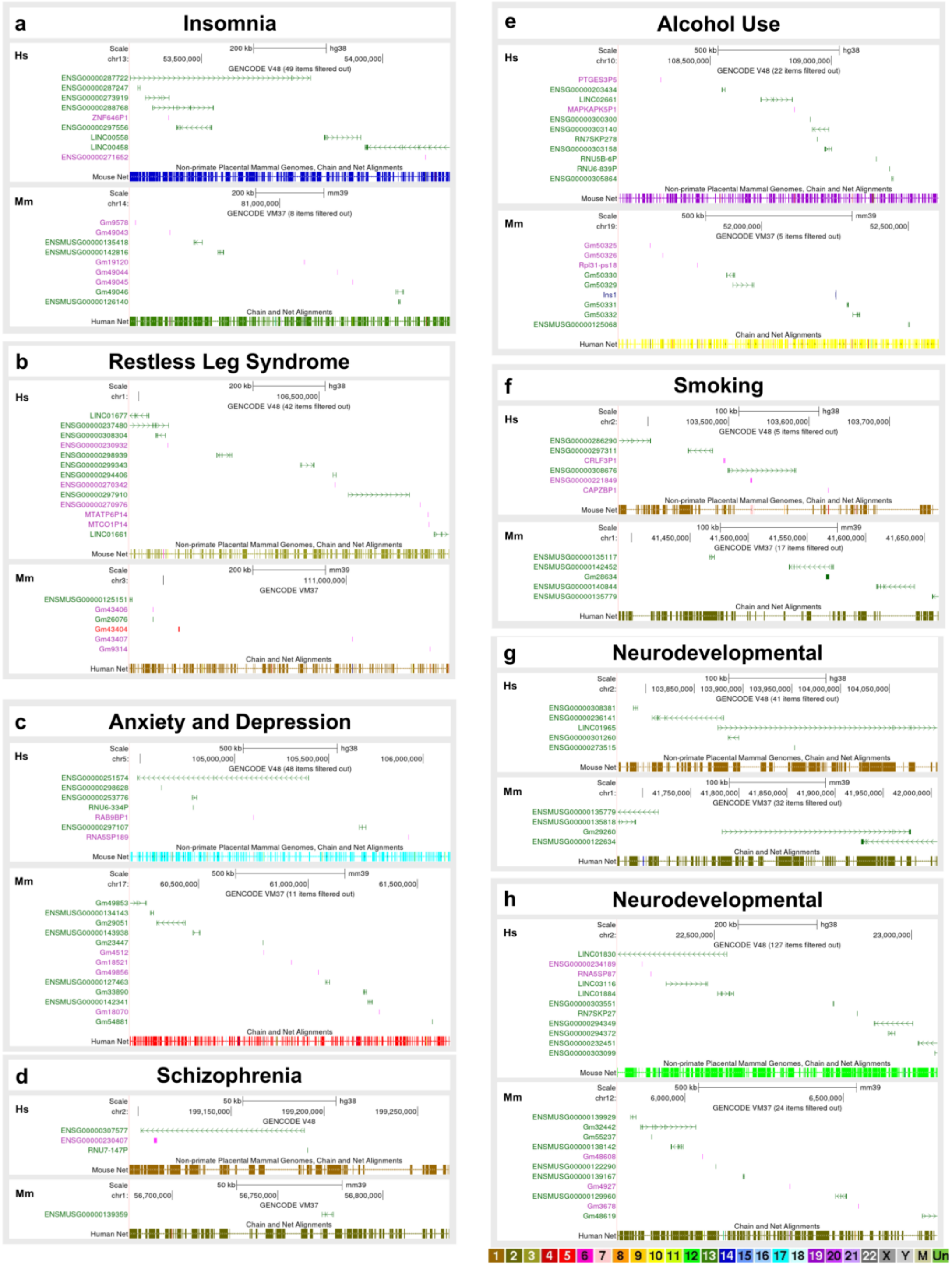

a
Insomnia
b
Restless Leg Syndrome
c
Anxiety and Depression
d
Schizophrenia
e
Alcohol Use
f
Smoking
g
Neurodevelopmental
h
Neurodevelopmental

**Figure S3. UCSC genome browser view of haplotype blocks associated with sleep, neuropsychiatric, and neurodevelopmental disorders by GWAS.** (**a**) A haplotype block containing 57 GWAS associations with insomnia and the mouse syntenic locus. (**b**) A haplotype block containing 7 GWAS associations with restless leg syndrome and the mouse syntenic locus. (**c**) A haplotype block containing 153 GWAS associations with anxiety and/or depression and the mouse syntenic locus. (**d**) A haplotype block containing 12 GWAS associations with schizophrenia and the mouse syntenic locus. (**e**) A haplotype block containing 28 GWAS associations related to alcohol consumption and the mouse syntenic locus. (**f**) A haplotype block containing 32 GWAS associations related to smoking and the mouse syntenic locus. (**g**) A haplotype block containing 7 GWAS associations related to neurodevelopmental disorders (attention deficit/hyperactivity disorder) and the mouse syntenic locus. (**h**) A haplotype block containing 8 GWAS associations related to neurodevelopmental disorders (autism and attention deficit/hyperactivity disorder) and the mouse syntenic locus. Chronograph of chromosome number applies to all browser images. Gene names: Green, annotated non-coding; pink, pseudogene; red, problem transcripts (retained introns, to be experimentally confirmed, or disrupted domains); blue, coding. Hs, *Homo sapiens*. Mm, *Mus musculus*.

#### Figure S4

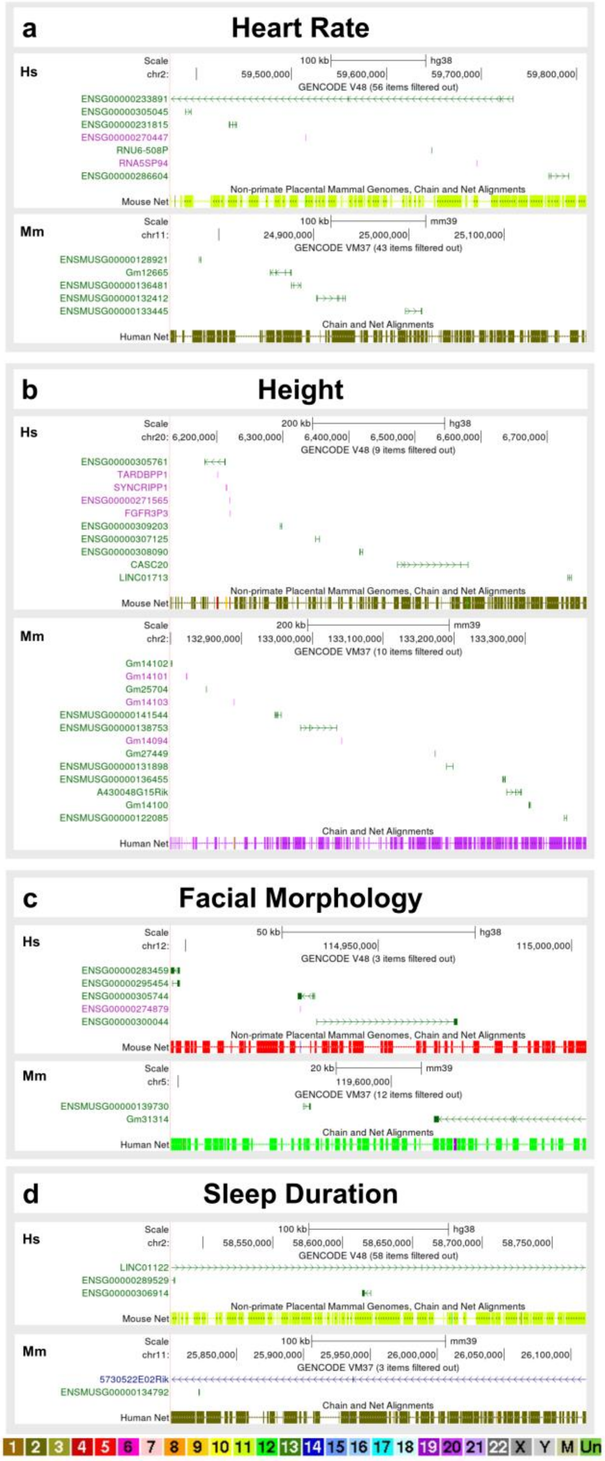


**Figure S7. UCSC genome browser view of haplotype blocks associated with body morphology and physiological traits by GWAS.** (**a**) A haplotype block containing 12 GWAS associations related to heart rate and the mouse syntenic locus. (**b**) A haplotype block containing 93 GWAS associations related to body height and the mouse syntenic locus. (**c**) A haplotype block containing 28 GWAS associations with facial morphology traits and the mouse syntenic locus. (**d**) A haplotype block containing 6 GWAS associations with sleep duration and the mouse syntenic locus. Chronograph of chromosome number applies to all browser images. Gene names: Green, annotated non-coding; pink, pseudogene; red, problem transcripts (retained introns, to be experimentally confirmed, or disrupted domains); blue, coding. Hs, *Homo sapiens*. Mm, *Mus musculus*.

#### Figure S5

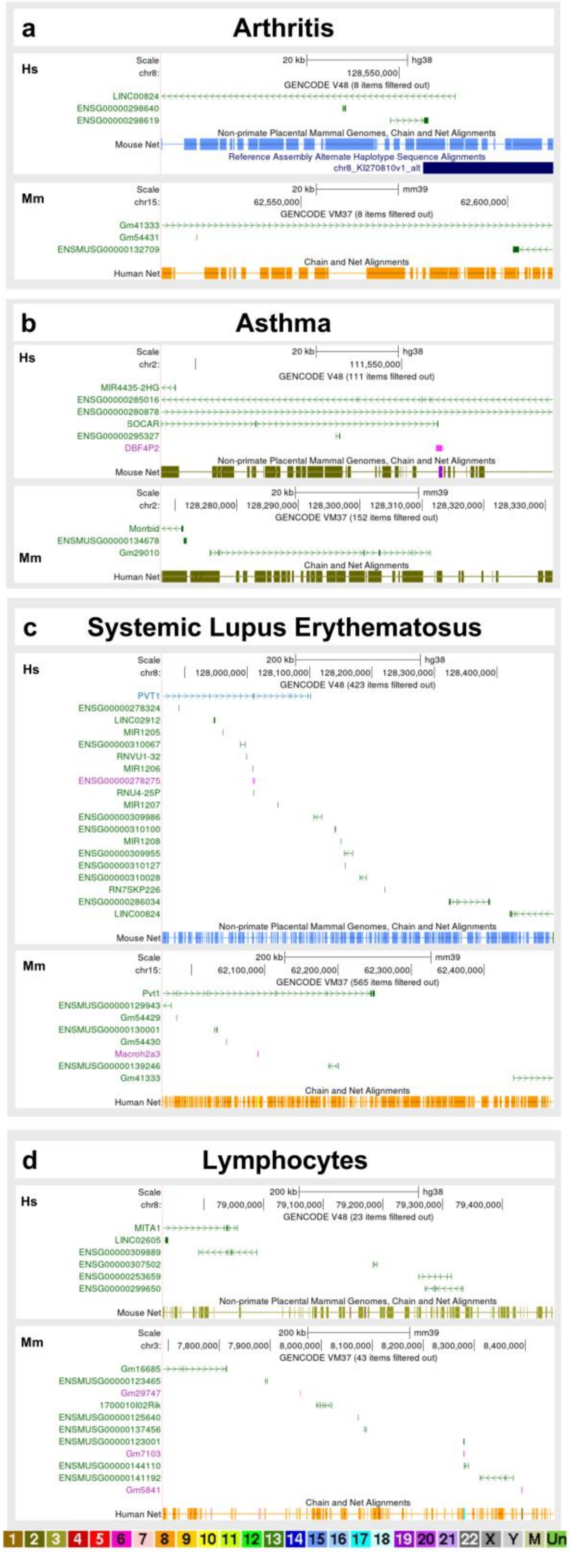


**Figure S5. UCSC genome browser view of haplotype blocks associated with autoimmune disorders and immune system traits by GWAS.** (**a**) A haplotype block containing 23 GWAS associations with arthritis and the mouse syntenic locus. (**b**) A haplotype block containing 6 GWAS associations related to asthma (11 autoimmune) and the mouse syntenic locus. (**c**) A haplotype block containing 7 GWAS associations with systemic lupus erythematosus and the mouse syntenic locus. (**d**) A haplotype block containing 9 GWAS associations related to lymphocyte count (14 immune cell count) and the mouse syntenic locus. Chronograph of chromosome number applies to all browser images. Gene names: Green, annotated non-coding; pink, pseudogene; red, problem transcripts (retained introns, to be experimentally confirmed, or disrupted domains); blue, coding. Hs, *Homo sapiens*. Mm, *Mus musculus*.

#### Figure S6

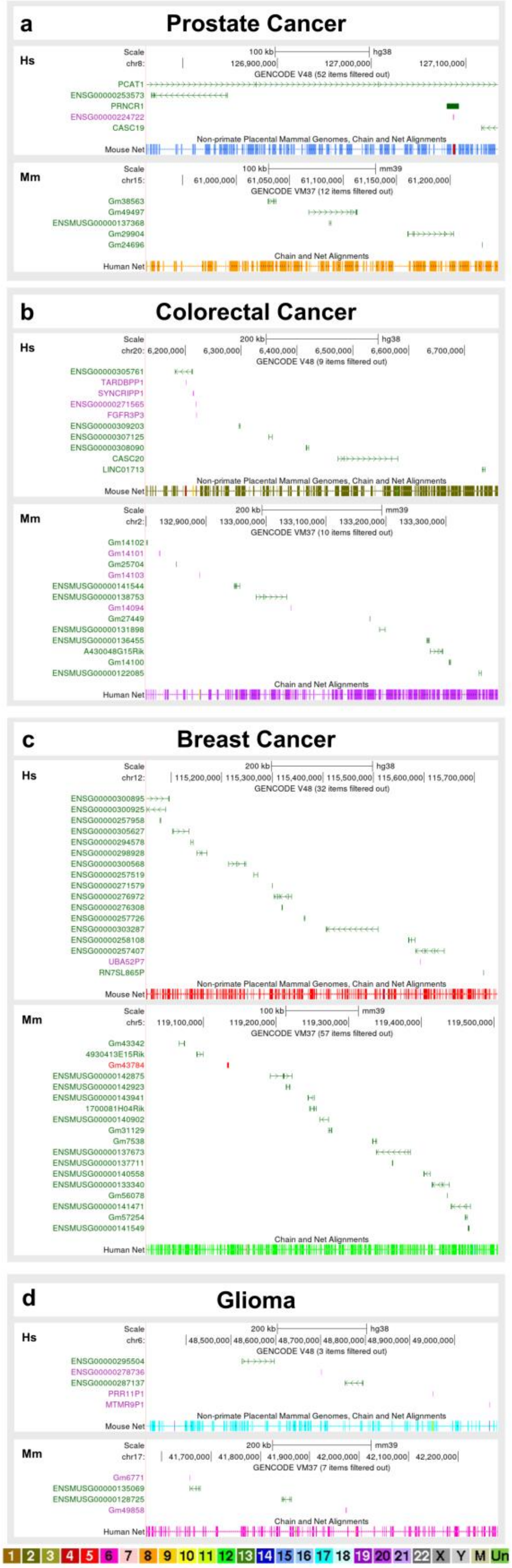


**Figure S6. UCSC genome browser view of haplotype blocks associated with cancer by GWAS.** (**a**) A haplotype block containing 185 GWAS associations with prostate cancer (202 with cancer generally) and the mouse syntenic locus. (**b**) A haplotype block containing 37 GWAS associations with colorectal cancer (40 with cancer generally) and the mouse syntenic locus. (**c**) A haplotype block containing 14 GWAS associations related to breast cancer (23 cancer generally) and the mouse syntenic locus. (**d**) A haplotype block containing 12 GWAS associations related to glioma/glioblastoma (13 cancer generally) and the mouse syntenic locus. Chronograph of chromosome number applies to all browser images. Gene names: Green, annotated non-coding; pink, pseudogene; red, problem transcripts (retained introns, to be experimentally confirmed, or disrupted domains); blue, coding. Hs, *Homo sapiens*. Mm, *Mus musculus*.

### Figure S7

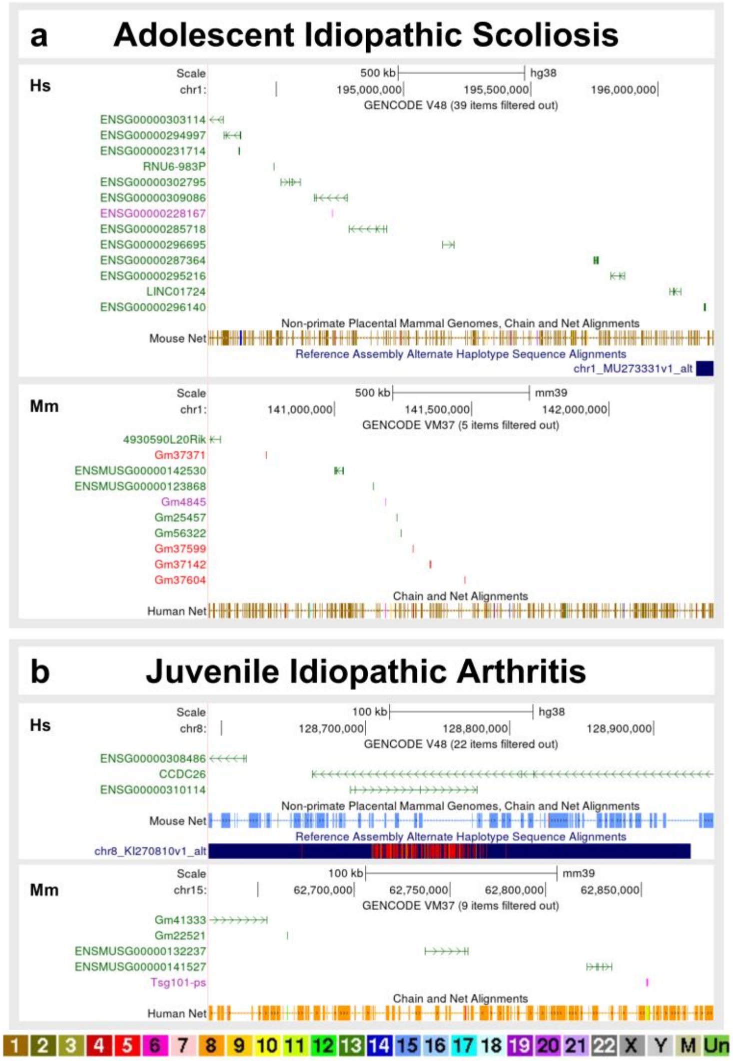


**Figure S7. UCSC genome browser view of haplotype blocks associated with idiopathic diseases by GWAS.** (a) A haplotype block containing 5 GWAS associations with adolescent idiopathic scoliosis and the mouse syntenic locus. (**b**) A haplotype block containing 2 GWAS associations with juvenile idiopathic arthritis and the mouse syntenic locus. Chronograph of chromosome number applies to all browser images. Gene names: Green, annotated non-coding; pink, pseudogene; red, problem transcripts (retained introns, to be experimentally confirmed, or disrupted domains); blue, coding. Hs, *Homo sapiens*. Mm, *Mus musculus*.

### Figure S8

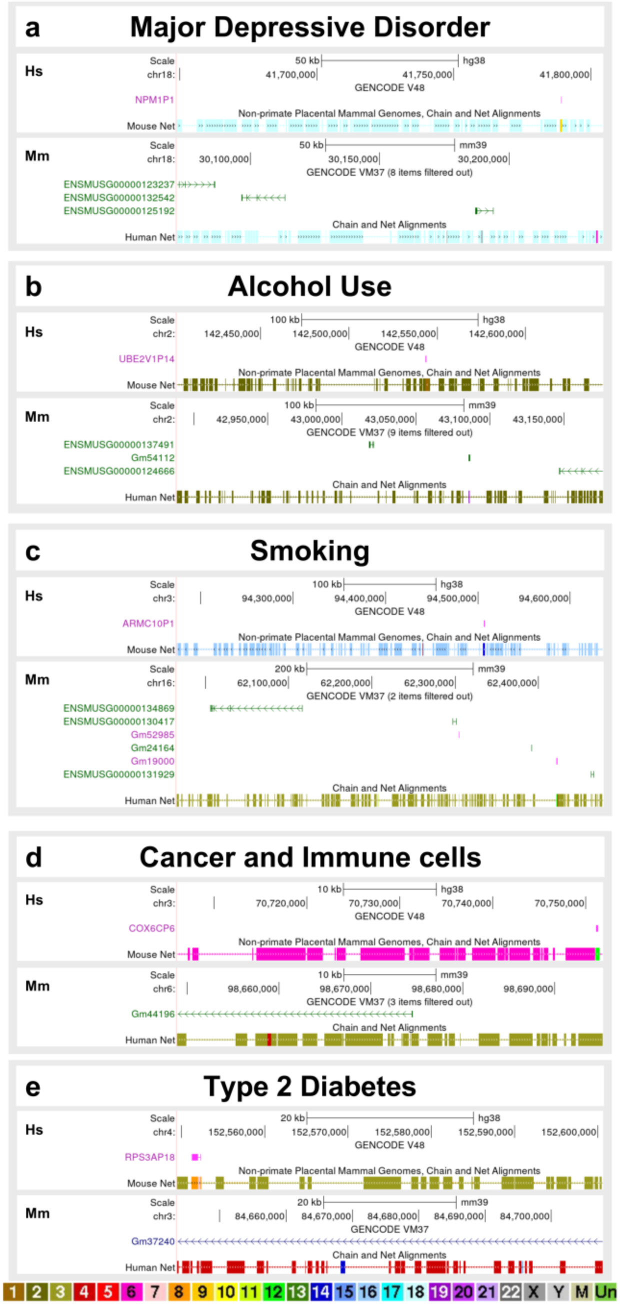


**Figure S8. UCSC genome browser view of haplotype blocks associated with GWAS traits but lacking a human gene.** (**a**) A haplotype block containing 21 GWAS associations related to neuropsychiatric disorders (27 neurological associations) and the mouse syntenic locus. (**b**) A haplotype block containing 6 GWAS associations related to alcohol consumption (10 neurological associations) and the mouse syntenic locus. (**c**) A haplotype block containing 7 GWAS associations related to smoking (24 neurological associations) and the mouse syntenic locus. (**d**) A haplotype block containing 3 GWAS associations related to prostate cancer and 3 related to immune cell numbers, and the mouse syntenic locus. (**e**) A haplotype block containing 28 GWAS associations related to diabetes mellitus and the mouse syntenic locus. Chronograph of chromosome number applies to all browser images. Gene names: Green, annotated non-coding; pink, pseudogene; red, problem transcripts (retained introns, to be experimentally confirmed, or disrupted domains); blue, coding. Hs, *Homo sapiens*. Mm, *Mus musculus*.
